# Leveraging mathematical models of disease dynamics and machine learning to improve development of novel malaria interventions

**DOI:** 10.1101/2021.01.05.21249283

**Authors:** Monica Golumbeanu, Guojing Yang, Flavia Camponovo, Erin M. Stuckey, Nicholas Hamon, Mathias Mondy, Sarah Rees, Nakul Chitnis, Ewan Cameron, Melissa A. Penny

## Abstract

**Background:** Substantial research is underway to develop next-generation interventions that address current malaria control challenges. As there is limited testing in their early development, it is difficult to predefine intervention properties such as efficacy that achieve target health goals, and therefore challenging to prioritize selection of novel candidate interventions. Here, we present a quantitative approach to guide intervention development using mathematical models of malaria dynamics coupled with machine learning. Our analysis identifies requirements of efficacy, coverage, and duration of effect for five novel malaria interventions to achieve targeted reductions in malaria prevalence. This study highlights the role of mathematical models to support intervention development.

**Methods:** A mathematical model of malaria transmission dynamics is used to simulate deployment and predict potential impact of new malaria interventions by considering operational, health-system, population, and disease characteristics. Our method relies on consultation with product development stakeholders to define the putative space of novel intervention specifications. We couple the disease model with machine learning to search this multi-dimensional space and efficiently identify optimal intervention properties that achieve specified health goals. We demonstrate the power of our approach by application to five malaria interventions in development.

**Results:** Aiming for malaria prevalence reduction, we identify and quantify key determinants of intervention impact along with their minimal properties required to achieve the desired health goals. While coverage is generally identified as the largest driver of impact, higher efficacy, longer protection duration or multiple deployments per year are needed to increase prevalence reduction. We show that the efficacy and duration needs depend on the biological action of the interventions. Interventions on multiple parasite or vector targets, as well as combinations the new interventions with drug treatment, lead to significant burden reductions and lower efficacy or duration requirements.

**Conclusions:** Our approach uses disease dynamic models and machine learning to support decision-making and resource investment, facilitating development of new malaria interventions. By evaluating the intervention capabilities in relation to the targeted health goal, our analysis allows prioritization of interventions and of their specifications from an early stage in development, and subsequent investments to be channeled cost-effectively towards impact maximization. Although we focus on five malaria interventions, the analysis is generalizable to other new malaria interventions.

## Background

Significant efforts to deploy malaria interventions worldwide have led to considerable progress and have reduced global malaria prevalence in Africa by half over the 2000 to 2015 period (1). Reductions were achieved through a diverse range of interventions including mass distribution of insecticide-treated mosquito nets, indoor residual spraying, rapid diagnosis, as well as artemisinin-based combination therapies. However, since 2015 progress has stalled, and several countries have seen an increase in malaria incidence of over 40% (2). Current interventions and malaria control programs are facing major challenges due to lack of funding, increases in drug and insecticide resistance and diagnostic-resistant parasites, as well as supply chain and deployment difficulties (3, 4). Strategic investment and timely development of novel interventions is crucial to maintain the progress made and to advance towards malaria elimination (2, 5). Two recent reports issued by the World Health Organization (WHO) (6) and the Lancet Commission (7) emphasize the need for novel malaria products, calling for a sustained investment in research and development (R&D).

Consequently, the malaria product development space has steadily expanded over the last 15 years. Novel products are diverse, ranging from therapeutic and immunological interventions, such as drugs and vaccines, to new vector control tools. Here we provide an overview of these novel interventions and explore the properties for a subset of key interventions.

With over 13 new drug compounds in early clinical development (2, 8), new antimalarial therapies will hopefully be available in the next five years (9). Nevertheless, emergence of drug resistance remains a threat for novel drugs, advocating for products that ensure sustained protection. Several malaria vaccine candidates are under development (10, 11), and after 30 years including phase 3 clinical trial and pilot implementation (12–14) the RTS,S/AS01 vaccine has potential to avert mortality in children in combination with other interventions (15–17), including as seasonal prevention (18). Several other vaccines are in phase 2 clinical studies, such as R21 which demonstrated a clinical efficacy of up to 77% (19). More recently, biological alternatives to vaccines include passive immunization with injectable small molecules (20) or monoclonal antibodies (21–23). Conferring protection against malaria during several months and being safe to administer during pregnancy, monoclonal antibodies are seen as potential interventions for seasonal malaria chemoprevention and protection for certain risk groups, with first human trials of monoclonal antibodies ongoing (7, 23).

Vector control has seen active development over the past years, with over 10 different categories of novel products aiming to reduce both indoor and outdoor mosquito biting (24–26). These include new insecticides (27), vector traps (28, 29), as well as genetically-altered mosquitoes that will eradicate mosquito populations (called ‘gene drives’) (30, 31). Furthermore, improved housing has been shown to significantly reduce indoor mosquito biting (32, 33), and effective house traps or lures are being developed to supplement traditional indoor interventions (34).

Malaria products under development are defined through Target Product Profiles (TPPs), which constitutes a vital reference for dialogue between various stakeholders (listed in the Methods section) to guide R&D investments. TPPs are dynamic documents used during the development of a cutting-edge medical product, defining its required characteristics to fulfill an unmet health need (35). Given the large amount of malaria interventions currently in the development pipeline, a systematic approach is essential to inform development decisions and prioritization of novel interventions to ensure a sustainable investment of resources and a fast pace of innovation. Currently, there is no approach systematically incorporating quantitative evidence and the aforementioned operational aspects in malaria product development (including intervention deployment, efficacy, duration, decay, and public health impact) from early development stages.

Mathematical models of malaria transmission dynamics can be used to bridge this gap, as they quantitatively estimate the impact of interventions while including considerable evidence of disease progression and transmission, host immunity, as well as environmental or health system dynamics and their interaction with interventions (36) (Fig. 1). These models have been used extensively to estimate the impact of malaria interventions and to optimize intervention packages for specific geographies (37, 38). Here an established individual-based malaria-transmission model, OpenMalaria (37, 39–43), has been used to simulate epidemiological disease and intervention dynamics to project the impact on public health, as has been employed to conduct several consensus modeling and validation studies (37, 41–43).

**Fig. 1.**
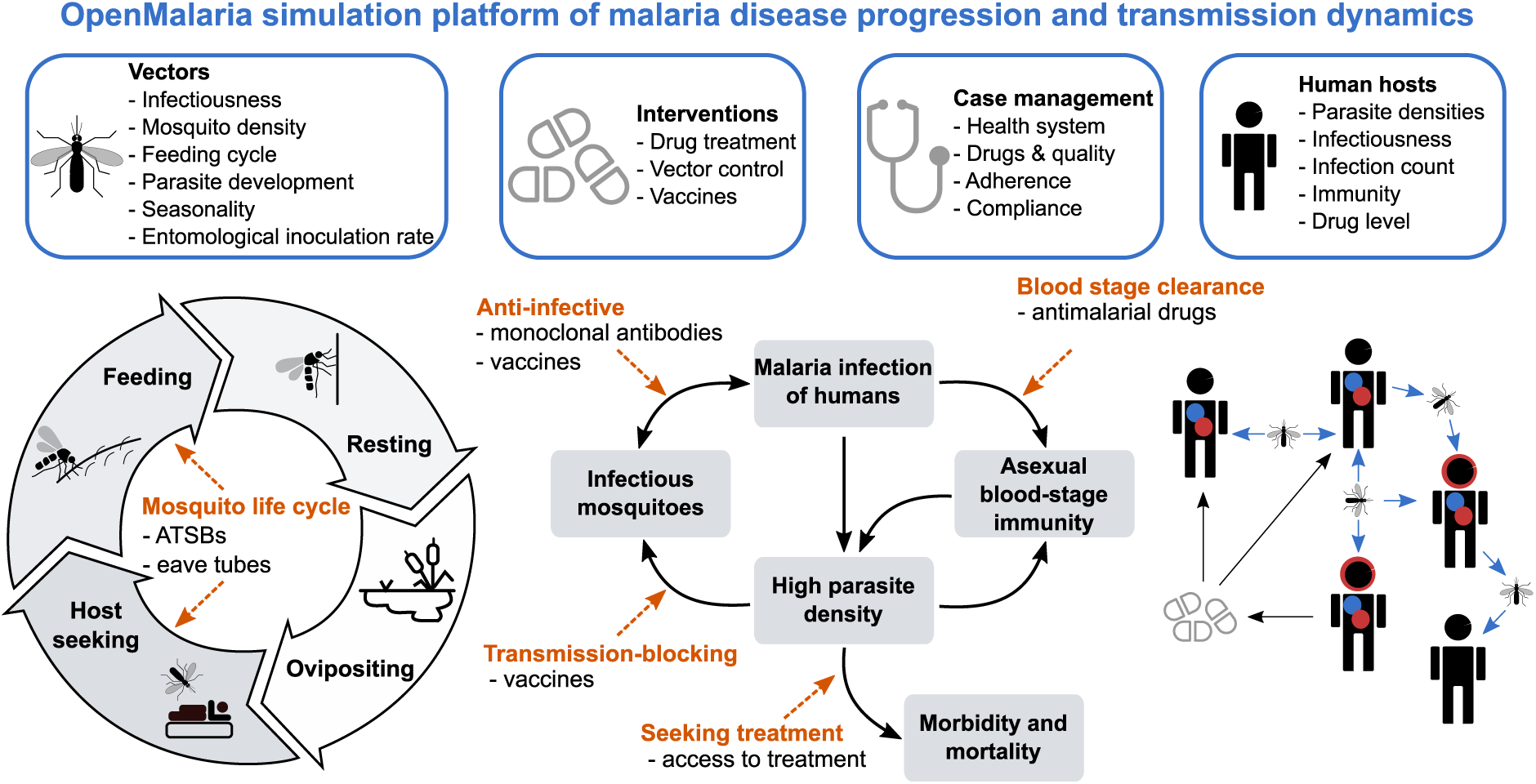
Simulation of malaria transmission dynamics with OpenMalaria: model schematic. Key components specified in the model are displayed on the top row including vector, intervention, case management, and human host-specific factors. Below, a detailed representation of the intrinsic modelled mosquito feeding and host transmission cycles is provided. The dashed, orange arrows mark the action and targets of the modelled interventions in this study, as indicated below the arrows. ATSBs stands for attractive targeted sugar baits. A detailed overview of model assumptions and parameters is provided in the Methods section and Additional file 1: section 1 and Tables S1.1-S1.3.

Due to absence of data at early intervention development stages and computational limitations in exploring a highly combinatorial parameter space of presumed intervention characteristics, models have mainly been used at late stages of intervention development. To date, models have only been minimally applied in directing the design of new interventions, or in understanding how intervention-specific, epidemiological, and systems factors jointly contribute to impact. Model investigations are usually informed by scenario analysis accounting for delivery and target age groups, as well as with properties of the new intervention pre-defined or informed by late clinical trials (41, 44, 45). In these constrained scenarios, high model and parameter complexity tend to obscure the complex relationships between intervention parameters, operational factors, health outcomes, and public health impact (46). Exhaustive scenario analyses are computationally expensive, rendering the full exploration of all possible interventions, in conjunction with all possible delivery scenarios, combinatorically infeasible. Previous approaches using disease models to inform TPPs have tackled the combinatorically complex parameter space by only exploring a discrete, constrained set of parameters (47–49). These approaches have provided insightful knowledge and have emphasized the importance of using disease models for defining TPPs. Nevertheless, they have provided a constrained view of intervention specifications.

Here, we propose a different approach (Fig. 2), where epidemiological models guide development of novel disease interventions designed to achieve quantified health goals from the early stages, placing the end goal of public health impact at the center of decision-making. To do this efficiently, we undertook an iteratively engaged exchange with malaria product development stakeholders to define desired outcomes and likely delivery use-cases of new malaria interventions. We then used mathematical models combined with machine learning to perform a directed search of the entire space of intervention profiles, to define properties of new interventions towards achieving the desired health goals. We used Gaussian processes (GPs) (50) to generate computationally light emulators that work in combination with the established OpenMalaria model of malaria dynamics. These emulators accurately captured the non-linear relationships between properties of deployed interventions, operational factors, and resulting health outcomes. They allowed efficient sensitivity analyses of intervention and health system parameters on predicted public health impacts at low computational cost. Furthermore, by coupling emulators with nonlinear optimization techniques, we constructed a predictive framework that identified key determinants of intervention impact as well as the minimal intervention profiles required for achieving a given health goal (Fig. 2).

**Fig. 2.**
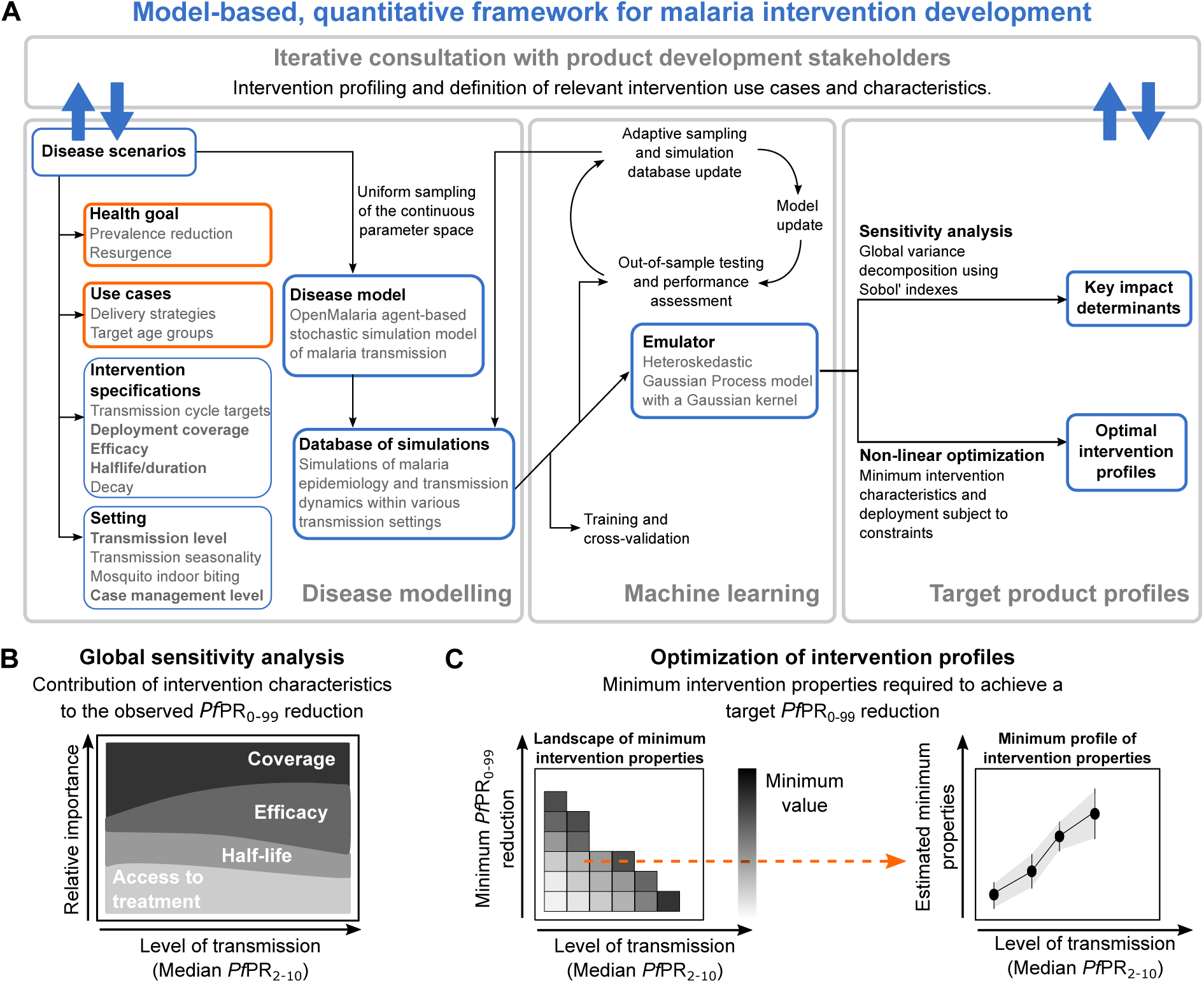
Overview of the approach to quantitatively define TPPs for novel malaria interventions. **(A)** Schematic description of the proposed model-based, quantitative framework to guide malaria product development. Results for applying this framework to guide development of five novel malaria interventions are provided for a range of simulated, true median *Pf*PR_2-10_ (before intervention deployment), and schematically described in subsequent figure panels. **(B)** Global sensitivity analysis for identifying the determinants of intervention impact: colors define intervention specifications, deployment coverage, and health system access levels varied in the analysis; the magnitude of the colored area at different levels of transmission (x-axis) represent the relative importance (y-axis) attributable to factors driving the observed *Pf*PR_0-99_ reductions following intervention deployment. **(C)** Optimization of intervention properties to achieve desired health goals: the heatmap (left panel) displays, for a given intervention property (coverage, efficacy, or half-life), the landscape of minimum required values to achieve various target *Pf*PR_0-99_ reductions. Each row of the heatmap corresponds to a target of *Pf*PR_0-99_ reduction and constitutes a minimum profile of the considered intervention characteristic (right panel).

We covered a diverse spectrum of interventions in the malaria development space, pertaining to 1) anti-infective monoclonal antibodies, 2) anti-infective vaccines, 3) transmission-blocking vaccines, 4) outdoor attractive targeted sugar baits, and 5) eave tubes. We used our approach to understand the link between intervention characteristics and resulting impact, and to define the requirements of these interventions in terms of coverage, efficacy, and impact duration to reach desired prevalence reduction goals, contingent on operational constraints (Fig. 2). We show how modelling can support the development process, and introduce a framework that quantitatively defines malaria product characteristics within TPPs. Our approach illustrates how modelling enables translation of R&D efforts into potential impact.

## Methods

The approach introduced here combines infectious disease modeling with machine learning to understand determinants and define quantitative properties of target product profiles of new malaria interventions. The methodological approach is schematically outlined in Fig. 2 with a full description in Additional file 1.

### Stakeholder engagement

There was active engagement and regular exchanges with different expert groups during development of the methodological framework for guiding TPPs of novel malaria interventions as described in full details in Additional file 1: section 1.1. Stakeholders involved in these discussions were the Bill and Melinda Gates Foundation (BMGF), the Innovative Vector Control Consortium (IVCC), the Program for Appropriate Technology in Health - Malaria Vaccine Initiative (PATH-MVI), and the World Health Organization (WHO). Guided by the BMGF, the stakeholder engagement process included initial meetings, interim meetings to define analyses, and presentations of results.

### The model

We used OpenMalaria v38.0 (39, 40), an established open-source stochastic, individual-based model to simulate malaria epidemiology and transmission dynamics across humans and mosquitoes in various settings with an overview in Fig. 1 and fully described in Additional file 1: section 1.2 and Tables S1.1-S1.3, with source code available from https://github.com/SwissTPH/openmalaria/. The OpenMalaria model was calibrated and validated in previous studies using historical epidemiological data (39, 40, 51), and this calibration was used for this study (as fully described in Additional file 1: section 1.2.1-1.2.2, including model components, core parameters and mosquito cycle dynamics in Tables S1.1-S1.3).

### Description of simulation experiments

The simulated human population size in this analysis was 10,000 individuals, with its age structure informed by health and demographic surveillance data for Ifakara, Tanzania (52). It is assumed that no infections were imported over the entire study period. Health system characteristics, mosquito entomological parameters driving infection patters, and seasonal exposure patterns are described in Additional file 1: sections 1.2.1-1.2.4, Figs. S2.1-S2.3, and Tables S1.1-S1.3. Parasite infections in simulated hosts are simulated individually and disease effects such as immunity, infectiousness to mosquitoes, morbidity, or mortality are tracked. Setting-specific characteristics include demographics, mosquito species entomological profiles are explicitly modelled and a wide range of human and vector interventions can be applied. Various health outcomes are monitored over time, including *Plasmodium falciparum* prevalence of infections (*Pf*PR), uncomplicated clinical or severe disease, hospitalization, and malaria mortality. Model assumptions have been described and validated with field data in previous studies (53, 54).

### Definition of intervention profiles their impact and health goals

We built a standardized representation for each malaria intervention. Accordingly, a malaria intervention was characterized according to the targets of the transmission life cycle it affects, along with the efficacy, half-life, and decay (Fig. 1, Additional file 1: Fig. S2.2 and Table 1). Descriptions of quantification of the efficacy of a given therapeutic or immunologic intervention, as well as definitions of intervention targets (as shown in Fig 1) are provided in Additional file 1: section 1.2.3. Each intervention or combination of interventions was applied as mass intervention targeting to all ages equally, along with continuous case management. Additional file 1 also provides descriptions of deployment of mass intervention packages (section 1.2.3), how the input the EIR was translated to *Pf*PR_2-10_ (section 1.2.4), as well as definitions of intervention impact and health goals (section 1.2.5).

**Table 1.**
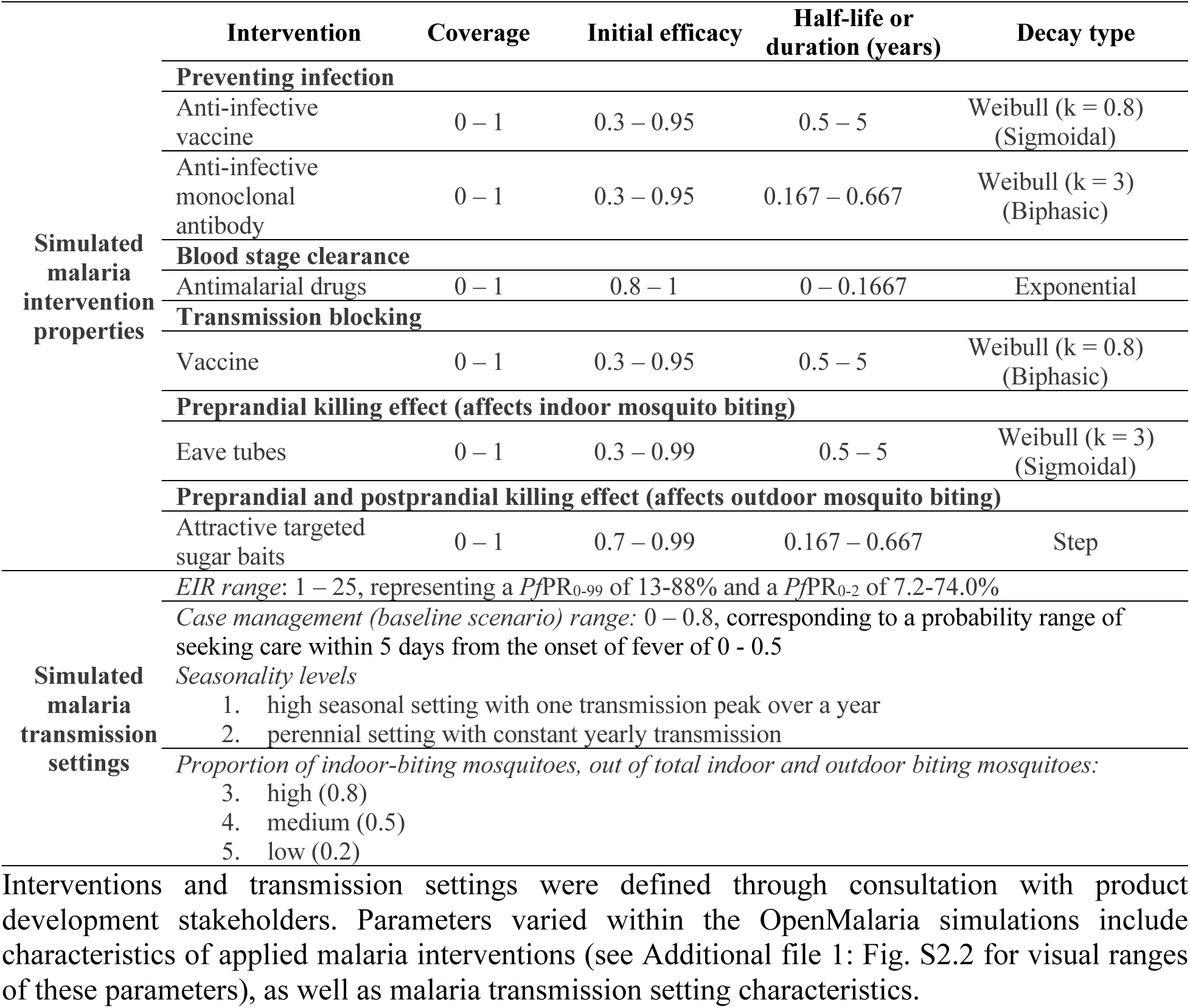
Description and ranges of OpenMalaria simulation interventions and transmission settings.

### Disease model emulator with Gaussian processes

As it was computationally intensive to simulate an exhaustive number of simulations to explore the entire parameter space for diverse combinations of interventions, settings, and deployments, machine learning techniques and kernel methods were applied. A full description of how the model emulator was built with Gaussian processes is provided in Additional file 1: section 2.1, along with a description of how the training dataset was built for each intervention and setting (section 2.2) and how emulators were trained on this dataset (section 2.3).

### Identifying impact determinants through sensitivity analysis

To estimate the contribution of each model input and its interactions with the other inputs to the variance of the model outcome, a global sensitivity analysis based on variance decomposition (55) was conducted. A complete description of the sensitivity analysis process can be found in Additional file 1: section 4.1.

### Finding minimal intervention properties

The trained GP models for each transmission setting and intervention were used within a general-purpose optimization scheme to identify minimum intervention properties that reach a defined *Pf*PR_0-99_ reduction goal given operational and intervention constraints. The calculations for finding the minimal intervention properties are provided in Additional file 1: section 5.1.

## Results

We present a disease model and machine learning approach to quantitatively define malaria interventions. Our approach (Fig. 2) started with an iterative consultation with product development experts to build sensibly informed TPP simulation scenarios, i.e., to define the breadth, range, and intervention profiles to simulate with OpenMalaria for the five malaria interventions considered. Accordingly, we collaborated with the BMGF, and the PATH-MVI and IVCC product development partnerships. For each new intervention in the portfolios for PATH-MVI, IVCC, and others, we conducted several expert group discussion sessions to catalogue the ranges of potential intervention effectiveness; potential delivery strategies; parasite or vector targets; and likely properties (age target, mass intervention, yearly deployment) in terms of mode of action (target), efficacy, and duration; and use-cases/delivery (see Methods and Additional file 1: section 1 for a description of the iterative stakeholder engagement process).

The public health goal in this study is to reduce the prevalence of *Plasmodium falciparum* malaria (denoted as *Pf*PR_0-99_ when evaluated for all ages and *Pf*PR_2-10_ when evaluated for 2-10-year-olds) between years one and three following deployment (Fig. 3A). We modelled each intervention by identifying their action on parasite or vector targets during the malaria transmission cycle (orange arrows in Fig. 1). Accordingly, each intervention was defined by its target, the ranges of the deployment coverage, initial efficacy, half-life, or duration of effect as well as the type of efficacy decay (see section 2 of the Methods and Additional file 1: section 1, Fig. S2.2 and Table 1 for detailed intervention specifications). For simplification, the words ‘half-life and ‘duration’ are used interchangeably to describe the longevity of an intervention effect. The simulated malaria transmission settings were defined by the yearly EIR, seasonality level, access to treatment, as well as proportion of indoor mosquitoes. Table 1 summarizes the results of the stakeholder discussions to set-up the OpenMalaria simulation scenarios and presents a comprehensive description of all subsequent intervention characteristics explored in this study, as well as the simulated malaria transmission settings.

**Fig. 3.**
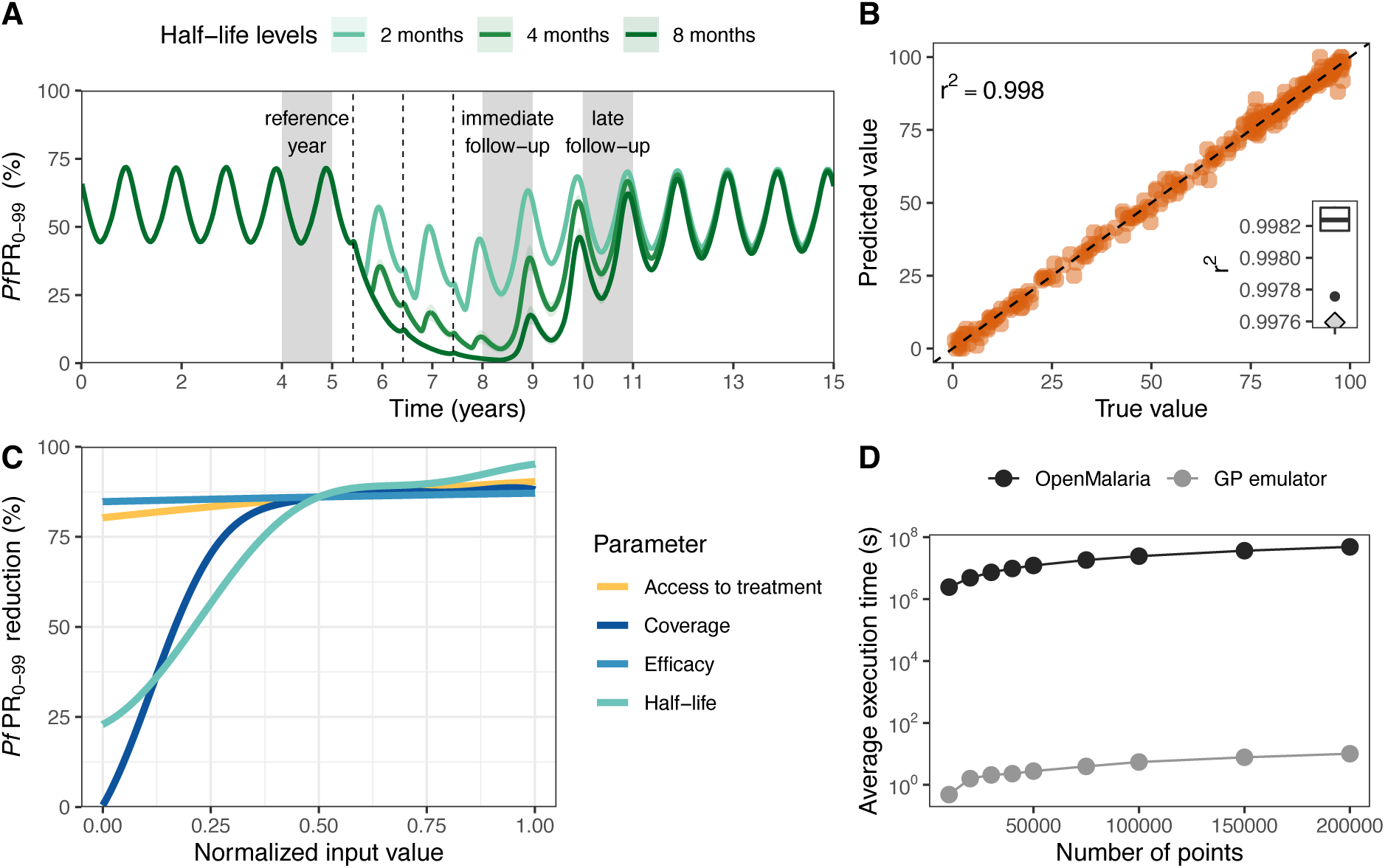
Training predictive GP emulators of simulated intervention impact with OpenMalaria. Examples are shown for attractive targeted sugar baits (ATSBs); results for other interventions are shown in Additional file 1: Figs. S3.1 and S4.2-S4.7 and Table S4.1. **(A)** Simulated malaria *Pf*PR_0-99_ time series at EIR=10 where ATSBs were deployed at a coverage of 70% and had an efficacy of 70%. Results are shown for three intervention half-life levels. The dotted lines indicate when interventions were applied (beginning of June). The effect of the interventions was assessed by evaluating the yearly average *Pf*PR_0-99_ reduction in all ages relative to the year prior to deployment (first grey block). Two outcomes were assessed, depending on whether the average prevalence was calculated over the year following deployment (immediate follow-up), or over the third year following deployment (late follow-up). **(B)** Correlation between simulated true (horizontal axis) and predicted (vertical axis) *Pf*PR_0-99_ reduction with a GP emulator trained to predict the immediate impact of ATSBs. The GP emulator was trained in a cross-validation scheme (distribution of the Pearson correlation coefficient *r^2^* shown in the boxplot) and validated on an out-of-sample test set (*r^2^* left upper corner and grey diamond lower right corner of the boxplot). **(C)** Relationship between each normalized input parameter and the resulting *Pf*PR_0-99_ reduction predicted with the trained GP emulator. Each parameter was in turn varied within its defined ranges (Table 1) while other parameters were set to their average values. **(D)** Estimated CPU execution time for varying sizes of input parameter sets evaluated with OpenMalaria (black) and with the trained GP emulator (grey).

Next, within the *“Disease model”* component of our approach (Fig. 2A), malaria transmission was modeled by the means of the established, stochastic, individual-based model OpenMalaria (39) (Figs. 1 and 3A, and Additional file 1: section 1 and Tables S1.1-S1.3). Here we used a previously published calibration of OpenMalaria, which reflects demographics, epidemiology, entomology, health system access, and seasonality (Additional file 1: Fig. S2.1) for a catchment area in Tanzania (56). Intervention impact was assessed through predicted reduction in *Pf*PR_0-99_, corresponding to true infection prevalence and not patent (detected with a diagnostic such as rapid diagnostic test (RDT) or polymerase chain reaction (PCR), Fig. 3A and Additional file 1: Figs. S2.3 and S3.1-S3.4). We simulated malaria epidemiology and transmission dynamics within various transmission settings based on the Tanzanian calibration. These settings cover a broad spectrum of transmission and mosquito biting behavior archetypes relevant for attaining general guiding principles in the early development phase of new malaria interventions. A comprehensive set of simulated scenarios was built by uniformly sampling the parameter space defined by intervention and transmission setting characteristics (defined in Fig. 2A and detailed in Table 1). These scenarios were simulated with OpenMalaria, yielding an extensive database of disease outcomes for the defined scenarios.

In the machine learning part of the approach (illustrated in the *“Machine learning”* panel of Fig. 2A), the database of simulated scenarios and corresponding outcomes (*Pf*PR_0-99_ reductions following intervention deployment) was used to train predictive models for the OpenMalaria simulation results (for an example of simulated *Pf*PR_0-99_ time series with OpenMalaria see Fig. 3A and Additional file 1: Fig. S3.1). A Heteroskedastic Gaussian process (GP) model was trained for each intervention and transmission setting (see detailed training procedure in section 3 of the Methods). Trained GP models accurately captured the dependencies between the disease model input parameters and the output intervention impact (Fig. 3B and C) and were able to reliably predict the reduction in *Pf*PR_0-99_ attributable to any input intervention characteristics in a given malaria transmission setting. Precisely, the correlation between true and predicted *Pf*PR_0-99_ reduction on out-of-sample test sets exceeded 95% while the absolute mean error was below 3% for all trained GP models (Fig. 3B and Additional file 1: Figs. S4.1-S4.3 and Table S4.1). As a result, the trained predictive GP models acted as emulators of the complex modelled parameter dynamics and non-linear relationships within the individual-based mathematical model of malaria transmission (Fig. 3C and Additional file 1: Figs. S4.5-S4.6) and could predict the disease outcome for the given health goal and for any set of input parameters.

Due to the significantly less intensive computational requirements of our emulator-based approach compared with OpenMalaria, we could reduce the analysis execution time by several orders of magnitude. This allowed us to conduct global sensitivity and optimization analyses, which required a large number of parameter set evaluations and would otherwise not have been possible (Fig. 3D and Additional file 1: Fig. S4.7). Thus, the trained GP emulators could be efficiently and promptly used in downstream analyses to explore the multi-dimensional space of intervention properties to design TPPs of new malaria interventions (panel *“Target product profiles”* in Fig. 2A), i.e., to identify the drivers of their impact and their quantitative properties in meeting the health goals previously defined (Fig. 2B and C). Specifically, through global sensitivity analysis, we identified the key determinants of intervention impact (Fig. 2B). In addition, we performed a constrained search for intervention and delivery profiles (TPPs) that maximized impact under a particular health goal, given concrete, expert-informed, operational constraints such as possible deployment coverage, or feasible intervention properties such as efficacy or duration of protection (Fig. 2C). Results of these analyses are detailed in the following sections and illustrated for seasonal transmission settings with high indoor mosquito biting. Results for the other simulated transmission settings (perennial settings and for other mosquito biting patterns) are provided in the supplement (additional sensitivity analysis results presented in Additional file 1: Figs. S5.1-S5.2, additional optimization results presented in Additional file 1: Figs. S6.1-S6.6 and S7.1-S7.5) and summarized in Table 2.

**Table 2.**
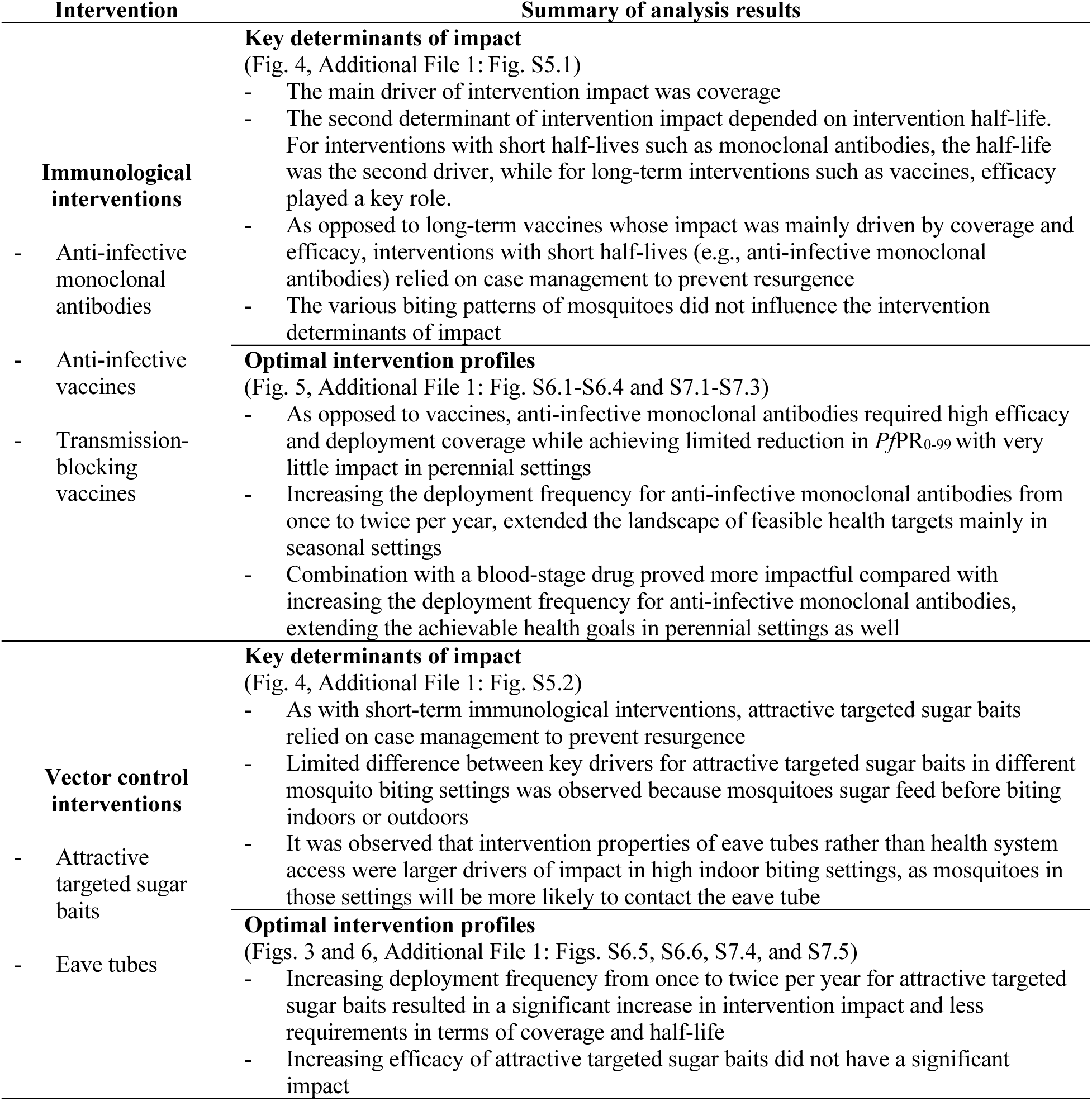
Key findings of our quantitative approach guiding target product profiles of new malaria interventions.

### Impact of malaria interventions and the importance of their characteristics

Following simulation with OpenMalaria of deployment of each of the studied interventions through mass administration campaigns over several years (see Methods), we first analyzed the predicted distributions of reduction in true *Pf*PR_0-99_. We found that, in general, when aiming for substantial, prompt reductions in prevalence for this particular health target, vector control was by far the most impactful intervention across all settings. Conversely, monoclonal antibodies, anti-infective and transmission-blocking vaccines had a more pronounced impact in low-transmission settings compared to endemic settings (Fig. 4A and Additional file 1: Figs. S3.1-S3.4 and Table 2).

**Fig. 4.**
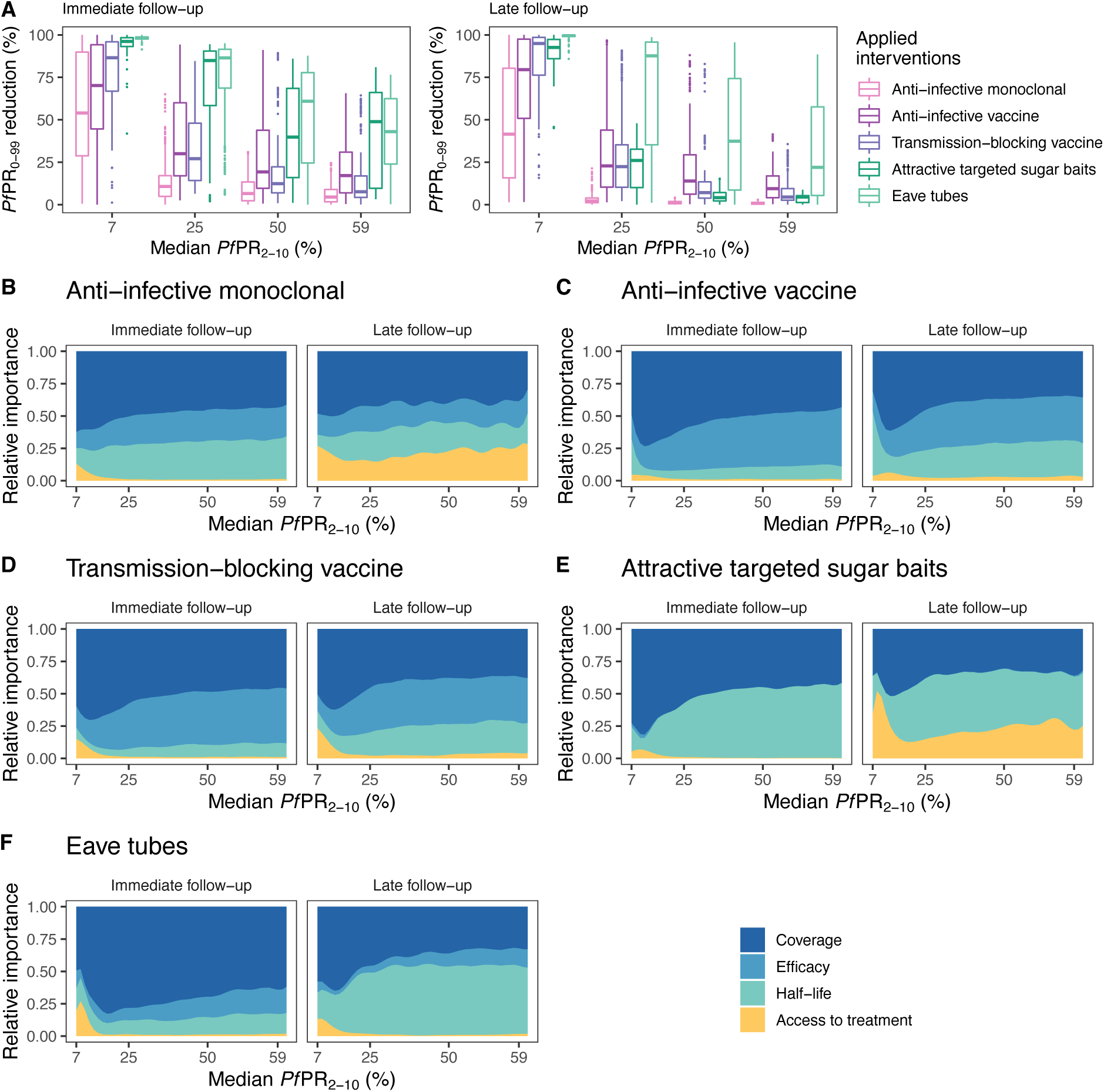
Effects of novel malaria interventions on *Pf*PR_0-99_ and their key drivers of impact. **(A)** Distribution of obtained reduction in *Pf*PR_0-99_ across the simulated scenarios with OpenMalaria following deployment of various malaria interventions under development (shown with different colors) for a range of simulated transmission settings (specified by median true *Pf*PR_2-10_ rounded values, x-axis). Each boxplot displays the interquartile range (box), the median value (horizontal line), the largest and smallest values within 1.5 times the interquartile range (whiskers), and the remaining outside values (points) of the *Pf*PR_0-99_ reduction values obtained across all the simulations for each given setting. The remaining panels present the results of global sensitivity analysis showing, across the same simulated *Pf*PR_2-10_ settings, the contribution of intervention characteristics to the resulting *Pf*PR_0-99_ reduction for anti-infective monoclonal antibodies **(B)**, anti-infective vaccines **(C)**, transmission-blocking vaccines **(D)**, attractive targeted sugar baits **(E),** and eave tubes **(F)**.

Sensitivity analysis indicated that the impact of these interventions on malaria prevalence was driven by different characteristics of their efficacy profiles, deployment strategies, or access to care for treatment of clinical cases, for short- and long-impact follow-up. Across a large proportion of the simulated scenarios, for all parasite and vector targets and interventions, deployed intervention coverage was overwhelmingly the primary driver of impact, especially in low-transmission settings (Fig. 4B-F and Additional file 1: Figs. S5.1-S5.2). For immunological interventions, the impact of short-term passive immunizations such as monoclonal antibodies relied on their deployment coverage and the health system (Fig. 4B and Additional file 1: Fig. S5.1). In contrast, for long-acting interventions such as vaccines, impact was driven by deployment coverage and efficacy (Fig. 4C and D and Additional file 1: Fig. S5.1). Highly efficient vector control interventions such as attractive targeted sugar baits had a strong effect on prevalence (Fig. 4A), and their duration of effect was the most important determinant (Fig. 4E and Additional file 1: Fig. S5.2). The immediate impact of long-term vector control interventions such as eave tubes was driven by deployment coverage, while their half-life was a key determinant for preventing resurgence (Fig. 4F and Additional file 1: Fig. S5.2). Determinants of impact were identified for both immediate and late follow-up when interventions were applied once per year for three years (full intervention specifications are provided in Methods and results for other settings and interventions are shown in Table 2 and Additional file 1: Figs. S5.1-S5.2).

### Minimal requirements of novel malaria interventions to achieve a defined health goal

For the five aforementioned malaria interventions, we explored their optimal properties for a broad set of *Pf*PR_0-99_ reduction targets, creating landscapes of intervention profiles according to their minimal characteristics across various transmission settings (Fig. 5 and 6 and Additional File 1: Figs. S6.1-S6.6 and S7.1-S7.5). These landscapes provide a comprehensive overview of the intervention potential capabilities and limitations in achieving a desired health goal. For example, as opposed to an anti-infective monoclonal antibody which required high efficacy and duration to achieve large *Pf*PR_0-99_ reductions in only a limited number of settings (Fig. 5A and B and Additional File 1: Figs. S6.1 and S6.2), attractive targeted sugar baits that kill mosquitoes also achieved a wider range of target *Pf*PR_0-99_ reductions in high-transmission settings (Fig. 6A and B and Additional File 1: Fig. S6.5). Similarly, while anti-infective and transmission-blocking vaccines had comparable requirements in achieving similar *Pf*PR_0-99_ reduction targets in settings with lower transmission (*Pf*PR_2-10_ < 30%), anti-infective vaccines showed a higher potential and reached additional targets in high-transmission, endemic settings (Fig. 5C-F).

**Fig. 5.**
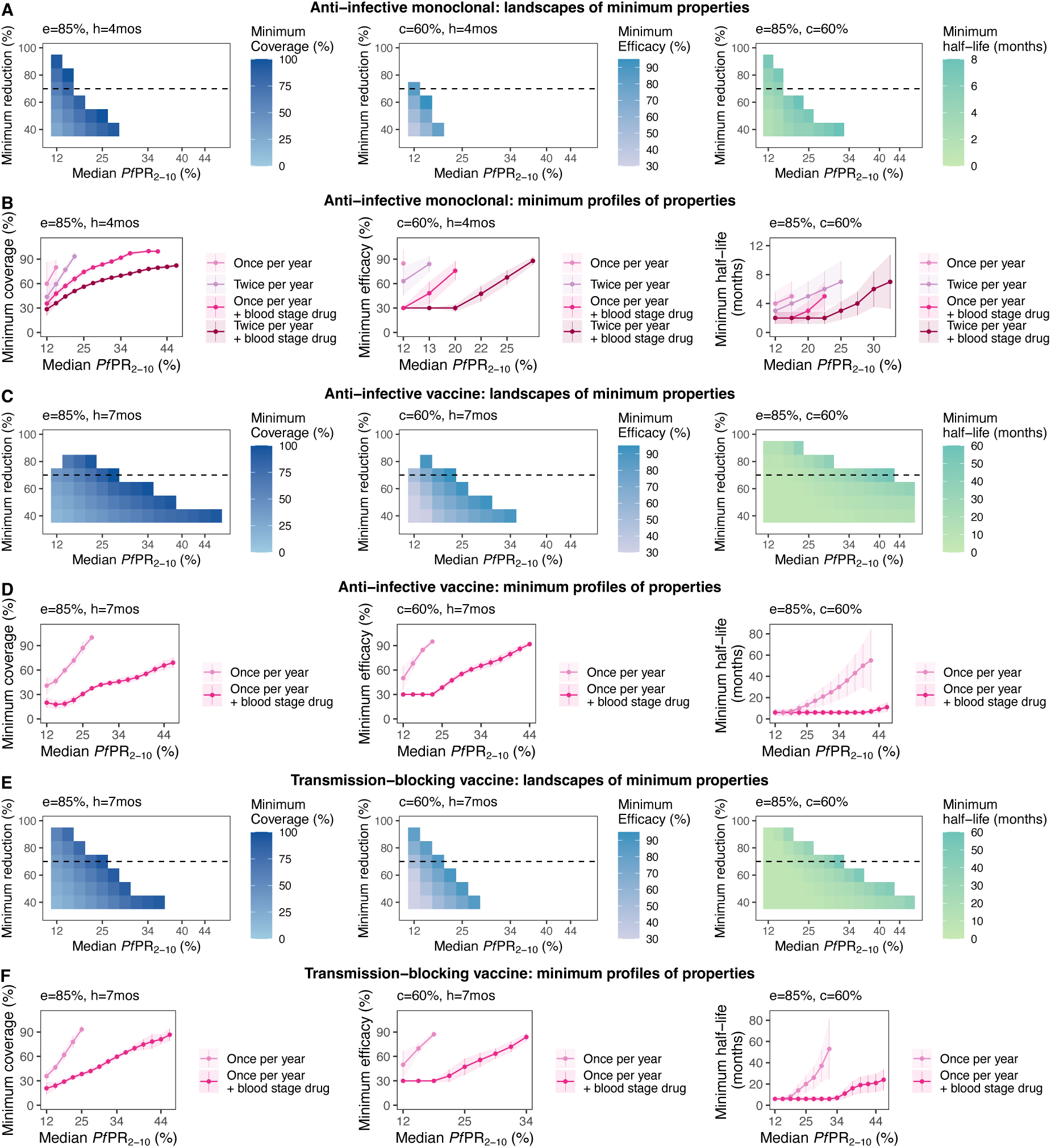
Estimated optimal intervention TPPs for immunological interventions. The heatmaps in panels **(A)**, **(C)** and **(E)** display, for each intervention property (coverage, efficacy, or half-life), the landscape of minimum required values to achieve various target PfPR_0-99_ reductions (y-axis) across different simulated transmission settings (true PfPR_2-10_ rounded values, x-axis). Each row of the heatmap corresponds to a target of PfPR_0-99_ reduction and constitutes the minimum required profile of the considered intervention. For a health goal of 70% PfPR_0-99_ reduction (dotted line on each heatmap), panels **(B)**, **(D),** and **(F)** present in detail how the minimum profile changes with transmission intensity. Each intervention characteristic was minimized in turn, while keeping other characteristics fixed (values marked on each panel where c = coverage, e = efficacy, and h = half-life). The simulated access to treatment, corresponding to a probability of seeking care within 5 days, was 25%. TPP=Target Product Profiles.

**Fig. 6.**
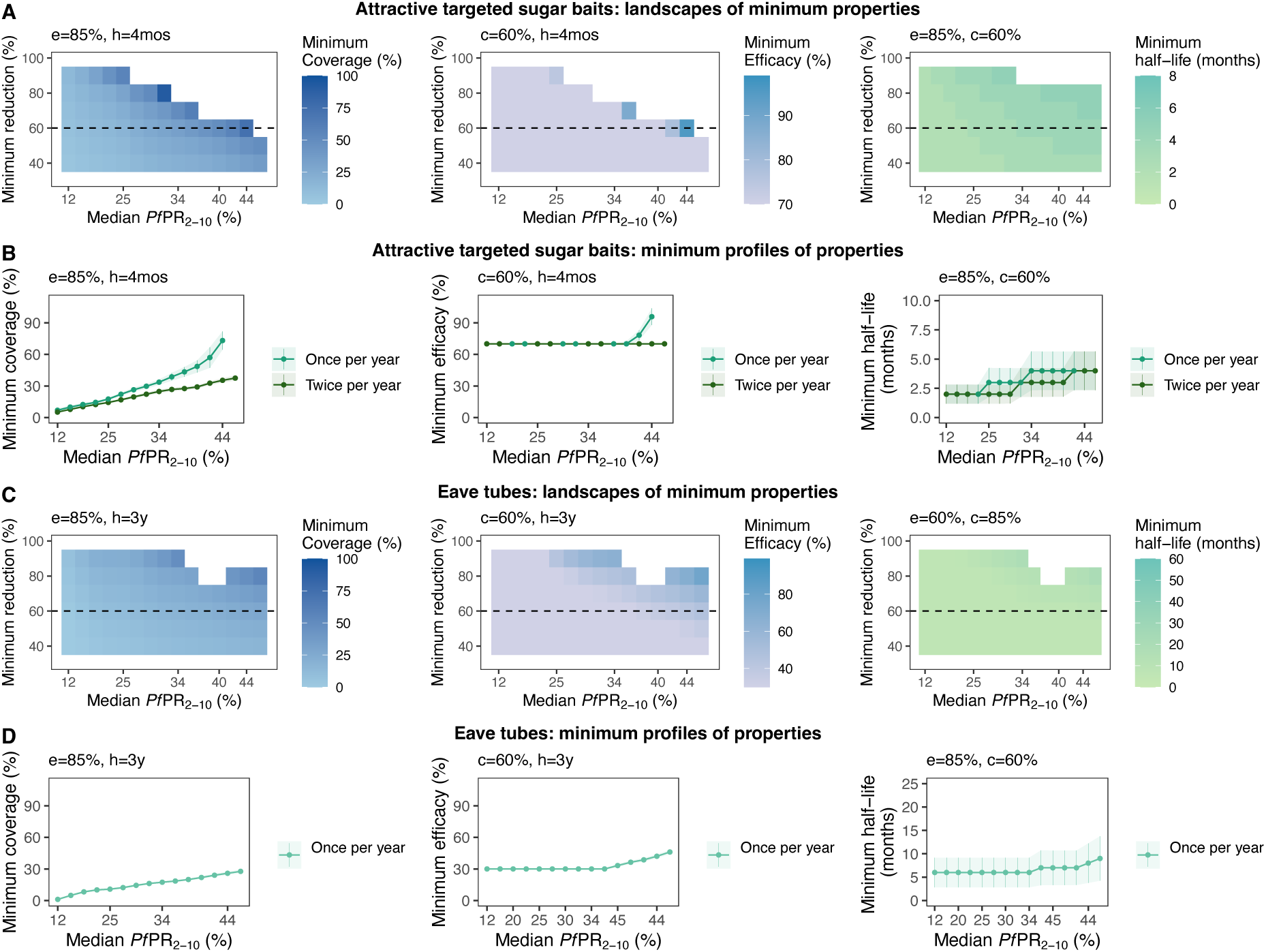
Estimated optimal intervention TPPs for vector control interventions. The heatmaps in panels **(A)** and **(C)** display, for each intervention property (coverage, efficacy, or half-life), the landscape of minimum required values to achieve various target *Pf*PR_0-99_ reductions (y-axis) across different simulated transmission settings (true *Pf*PR_2-10_ rounded values, x-axis). Each row of the heatmap corresponds to a target of *Pf*PR_0-99_ reduction and constitutes the minimum required profile of the considered intervention. For a selected health goal of 60% *Pf*PR_0-99_ reduction (dotted line on each heatmap), panels **(B)** and **(D)** present in detail how the minimum profile changes with transmission intensity. Each intervention characteristic was minimized in turn, while keeping the other characteristics fixed (values marked on each panel where c = coverage, e = efficacy, and h = half-life). The simulated access to treatment, corresponding to a probability of seeking care within 5 days, was 25%. TPP=Target Product Profiles.

For a detailed overview of landscapes of intervention profiles for all simulated settings and interventions see Additional File 1: Figs. S6.1-S6.6. These landscapes together with results of the sensitivity analysis offer an evidence-based prioritization of resources during product development. For example, we found that while both efficacy and half-life were important for immediate prevalence reductions with monoclonal antibodies, their effect was limited in preventing resurgence and was only supported by high case-management levels (Figs. 4 and 5 and Additional File 1: Figs. S5.1 and S6.1-S6.2). Conversely, the efficacy of anti-infective vaccines determined their immediate impact, whereas half-life of effect had greater importance for achieving and maintaining *Pf*PR_0-99_ reductions (Figs. 4 and 5 and Additional File 1: Figs. S5.1, S6.3, and S6.4). These results suggest that if monoclonal antibodies were to support preventing resurgence, then R&D efforts should focus on increasing and establishing antibody longevity.

Our analysis showed that coverage was a primary driver of impact (Fig. 4B-F and Additional File 1: Figs. S5.1 and S5.2). This has important implications for interventions requiring multiple applications to achieve high efficacy, indicating that it is of crucial importance to target both vulnerable populations and the proportion of the population missed by the intervention. While, for some interventions, high coverage deployment might be difficult or impossible to achieve, our analysis showed that this can be alleviated by increasing the deployment frequency or through deploying combinations of interventions, which may also have cost implications (Figs. 5B, 5D, 5F, and 6B, and Additional File 1: Figs. S6.1-S6.5 and S7.1-S7.4).

We found that combining several interventions targeting different stages in the transmission cycle can strongly affect the minimum requirements of a putative new intervention, potentially increasing the impact of an otherwise weaker intervention. For example, for an anti-infective monoclonal antibody with an initial half-life of 4 months that is deployed at a coverage of 60% reflecting completion of multiple doses, achieving 80% prevalence reduction was impossible when deployed once yearly for three years (Fig. 5A, and Additional File 1: Fig. S6.1). Furthermore, achieving the aforementioned health goal required an efficacy of over 80% when the intervention was deployed twice per year for three years (Additional File 1: Fig. S6.2). However, when monoclonal antibody deployment was coupled with a short half-life blood-stage parasite treatment such as dihydroartemisinin-piperaquine or artemether-lumefantrine, its minimum required efficacy was considerably reduced for both delivery frequencies (Fig. 5B and Additional File 1: Figs. S6.1, S6.2, and S7.1). Conversely, if an initial efficacy of 85% for the monoclonal antibody was assumed, its minimal required half-life could be reduced if this intervention was deployed in combination with a blood-stage parasite-clearing drug (Fig. 5B and Additional File 1: Figs. S6.1, S6.2, and S7.1). These results partly motivated the current development of anti-infective monoclonal antibodies; use-cases will likely include deployment with existing or new antimalarial treatment.

When coupled with a short half-life blood-stage parasite treatment, requirements of coverage, efficacy and half-life were also reduced for anti-infective and transmission blocking vaccines to achieve targeted reductions of *Pf*PR_0-99_ (Fig. 5C-F and Additional File 1: Figs. S6.3, S6.4, S7.2, and S7.3). In particular, for high-transmission settings (*Pf*PR_2-10_ > 30%), given an RTS,S-like half-life of seven months, both anti-infective and transmission-blocking vaccines could not achieve a defined prevalence reduction goal of 70% if deployed singly (Figs. 5D and 5F). This was the case for any deployment coverage given an initial efficacy of 85%, as well as for any efficacy given a 60% deployment coverage. Combining vaccine deployment with a blood-stage drug not only significantly expanded the achievable health targets in high-transmission settings, but also reduced vaccine properties requirements. Our analysis revealed that anti-infective vaccines had a higher potential than transmission-blocking vaccines, requiring less performance and achieving higher prevalence reductions targets in higher transmission settings. When combined with blood-stage parasite treatment, the coverage, efficacy, and half-life requirements of anti-infective vaccines were lower compared with those of transmission-blocking vaccines for the same prevalence reduction targets (Fig. 5 and Additional File 1: Figs. S6.3, S6.4, S7.2, and S7.3).

We also showed that a modified deployment schedule could reduce requirements for properties of some interventions. For example, for highly efficacious attractive targeted sugar baits, higher coverage and half-life were required when implemented once per year for three years compared with accelerated delivery of twice per year for three years (Fig. 6A and B). Except for high-transmission settings (*Pf*PR_2-10_ > 41%), a required efficacy of 70% was sufficient to attain the desired health goal for the majority of settings, for both delivery schedules (Fig. 6A and B, and Additional File 1: Figs. S6.5 and S7.4). This result was also reflected in the sensitivity analysis (Fig. 4E). Accordingly, the variation in intervention efficacy, across its investigated ranges, had little importance in driving the intervention impact. This suggests that, once a vector control intervention, such as attractive targeted sugar baits, has achieved a high killing efficacy (here ≥70%), a next step of optimizing other intervention characteristics, such as deployment coverage or duration, would lead to higher impact. These results demonstrate the strength of our analysis in identifying intervention characteristics to be prioritized for R&D.

Our comprehensive analysis was applied to explore determinants of impact and required profiles of interventions across two seasonal settings (seasonal and perennial) and three types of indoor mosquito biting patterns (low, medium, and high). A detailed overview of impact determinants and optimal intervention profiles is presented in Additional File 1: Figs. S6.1-S6.6 and S7.1-S7.5, with additional key results summarized in Table 2.

## Discussion

In this study, we introduced a quantitative framework, using detailed simulation models of malaria transmission dynamics that enables for the first time a quantitative differentiation between operational, transmission setting, and intervention parameters to better understand the potential impact of novel interventions against malaria. The framework consists of (i) a comprehensive disease progression and transmission simulation model applied on a discrete, uniformly sampled set of input parameters; (ii) training of a GP emulator on the sampled set of parameters and corresponding impact outcomes; (iii) using sensitivity analysis to understand drivers of intervention impact; and (iv) applying a non-linear constrained optimization algorithm to explore intervention operational and effectiveness characteristics meeting various targets and deployment use-cases specified following iterative consultation with product development experts. Our work thus builds on recent applications of GPs in disease modelling and burden prediction for malaria (57).

The value of our approach is realized through iterative collaboration with product development experts, by providing model-based guidance throughout the development process, and by refining feedback on model predictions as interventions progress through development. For malaria, where multiple interventions are in development, it also offers an approach for product developers from diverse fields (such as therapeutics and insecticide development) to collaborate and incorporate knowledge of other interventions into their TPP development. Coordinated by BMGF, the exchanges with stakeholders ensured a crucial discussion environment, guiding and supporting the methodology at various levels, from intervention profiling and defining relevant intervention use-cases to shaping research questions and subsequent analyses. This framework has been presented and validated in the presence of stakeholders in successive meetings. Discussions contributed to refinement of the investigation of various intervention profiles and led to an exploration of intervention combinations. Consequently, iterative exchanges with stakeholders have not only shaped the study approach, but had proven the value of this methodological framework in its versatility to adapt and address key questions along the product development pathway.

Using a detailed individual-based malaria transmission model like OpenMalaria brings significant advantages compared to a simpler and computationally less intensive Ross-MacDonald model. Individual-based models capture interactions between hosts, vectors, and parasites at the individual level, which provides a more realistic representation of the nonlinear transmission and epidemiological processes in the population as well as of the stochasticity of the modelled system (58). Second, it allows a realistic implementation of interventions, allowing for explicitly modeling intervention characteristics relevant for defining TPPs such as deployment regimes, efficacy, half-life, or shape of decay as well as explicit action at the different stages of the transmission cycle. Conversely, with Ross-MacDonald models, the modelled interventions have a very simplified representation through their effect only on transmission rates and this representation does not scale to the required detailed specifications of interventions in TPP documents.

The machine learning layer of our approach builds a simplified approximation of the relationships between intervention characteristics and resulting intervention impact. This approximation is very specific to the varied components across the model simulations (intervention characteristics, access to treatment and EIR) and to the resulting *Pf*PR_0-99_ reduction and does not explicitly provide a low-dimensional representation of all the processes captured by OpenMalaria. The complex disease dynamics processes captured by OpenMalaria (for a detailed description see Additional File 1: Table S1.1) are intrinsically captured when training the GP emulator and cannot be disentangled from the observed effects of the interventions per se. Furthermore, these relationships are non-linear and change with the transmission intensity (EIR) as shown in Additional File 1: Figs. S4.5 and S4.6. Therefore, to allow translation with product development processes, our approach for guiding novel interventions requires a model of disease transmission that is able to capture the non-linear disease dynamics processes and offers a realistic representation of interventions and their effects.

Although in this analysis we used reduction of *Pf*PR_0-99_ as a health goal, this method can be applied to other continuous disease burden statistics as required (see Additional File 1: Fig. S4.4 for performance predicting malaria incidence reduction). The same rationale applies for investigating other deployment strategies, required doses of interventions, or additional intervention combinations. However, this approach, which uses a smooth GP model, is not tailored for classification and categorical health goals. Nevertheless, it could be adapted to these types of outputs by replacing the GP emulator with a predictive model/alternative algorithm suited for categorical data, such as support vector machines. While sensitivity analysis would still be applicable for identifying the drivers of the categorical outcomes, the optimization questions and analyses would need to be reformulated to be relevant for the chosen categorical outcomes.

While bringing valuable quantitative insights to guide product development, our analysis of novel malaria interventions reproduces previous findings concerning intervention characteristics that are key drivers of impact. Previous studies have shown that intervention coverage is a major determinant of impact in the context of mass drug administration (41), of vaccines (59), as well as of vector control (26). Furthermore, this analysis reaffirms previous work showing the ability of vector control interventions to achieve substantial reductions in malaria burden (60).

This approach constitutes a powerful tool to help address the challenges of current malaria strategies and develop new interventions to progress towards malaria elimination. While currently promising interventions such as insecticide treated nets, seasonal malaria chemoprevention (SMC), and intermittent preventative treatment (IPT) have been very successful at reducing malaria incidence and saving lives, their improved burden reduction and future success is being challenged by limited adherence, limited resources, and time constraints towards increasing coverage and usage in underserved populations, as well as resistance (61). Furthermore, for settings where SMC is not recommended or has not been implemented (for example in East Africa or in perennial settings), there remains a gap in available interventions to protect vulnerable populations who experience the highest burden of malaria. Similarly, for settings with outdoor biting mosquitoes, development and rollout of novel vector control interventions is needed. New therapeutics and immune therapies suitable for seasonal delivery, such as long-acting injectables or monoclonal antibodies, are being developed and may close the development gap (21, 22). However, to efficiently make decisions on their development, guidance on their key performance characteristics and definition of their TPP is needed from early stages.

This quantitative framework can support the development of interventions from the beginning by generating evidence to inform and define evaluation criteria ensuring new products meet relevant health targets, while considering how these products may affect disease burden and epidemiology within a population. As shown here, this relies on iterative dialogue with stakeholders, to first define health targets, simulated scenarios, achievable intervention properties, and operational settings. The modelling part of the framework incorporates all this information as well as relevant disease transmission dynamics, building an *in-silico* system for testing developed interventions. Next, the sensitivity analysis part of the framework informs which intervention characteristics drive impact and are thus crucial in achieving the defined health goal. Providing insights on the development processes to be prioritized. Finally, the optimization analysis part of the framework reveals the potential of the developed intervention and how its efficacy and coverage requirements change according to the defined health targets and deployment setting. The landscapes of intervention profiles help product developers gauge development and investment efforts and select promising products. Furthermore, our approach allows investigating combinations of new and existing interventions, identifying alternatives to alleviate shortcomings such as coverage limitations. To achieve a final TPP, several iterations of this analysis are required, to ensure that the optimal tradeoffs between intervention capabilities and target goals for a given setting are best achieved.

As with all modelling studies, this approach is subject to several limitations. While the emulators capture not just the mean tendency of complex disease models dynamics, but also the inherent output variance caused by the stochasticity in the models (62), the estimations provided in this study are dependent on the performance of the trained emulator. This challenge was addressed with extensive adaptive sampling and testing to ensure a high level of accuracy of the trained emulators (Fig. 3, and Additional File 1: Figs. S4.1-S4.3 and Table S4.1). Despite the intrinsic uncertainty, this framework is intended to provide guiding principles and an efficient means of exploring the high dimensional space of intervention characteristics that otherwise would not be possible. Evidently, this analysis relies on the representativeness of model assumptions of disease and transmission dynamics as well as of expert opinion of likely intervention parameterizations in absence of clinical knowledge. Lastly, this analysis only explored a subset of use-cases, transmission settings, and intervention combinations. Future work should focus on the most likely settings and relevant use-cases as interventions are being developed and corresponding TPP documents are being refined.

Moving beyond the work presented in this paper, this framework would allow combining simulation models with other sources of data describing geographical variation in disease, for example, modelled health systems or modelled prevalence (63) and would allow incorporating interactions of interventions with novel interventions for surveillance. Clinical trials for new interventions could thereby be prioritized to geographical settings, where public health impact is likely to be maximized, and where appropriate, to inform decisions on achieving non-inferiority or superiority endpoints (64). A significant extension would be to incorporate economic considerations that may affect development decisions, including both R&D costs, as well as implementation and systems costs for final deployment.

## Conclusions

In this work, we provide mathematical tools for efficiently and quantitatively defining the minimum profiles of malaria interventions, as well as delivery approaches required to reach a desired health goal. Our framework can be extended and used for any disease where a valid model of disease progression or natural history of disease is available. It can be used to direct the design of novel interventions and to better understand how intervention-specific, epidemiological and systems factors jointly contribute to impact. Most immediately, this approach is highly relevant to define successful interventions against new diseases, and to support efficient, fast development of operational strategies. As uncertainties in disease progression and epidemiology can be incorporated in our approach by accordingly adjusting the putative parameter space of intervention characteristics, it also provides a way to systematically sort through large complex landscapes of unknowns and refine properties of interventions following clinical trials as more knowledge becomes available.

Our framework tackles and moves beyond current challenges in product development. On one hand, it allows rigorous definition of TPPs by efficiently exploring highly complex parameter spaces of disease models, and on the other hand, it allows determinants of desired public health impact to be identified to inform tradeoffs between product characteristics and use-cases.

## Data Availability

All the analysis code used in the paper as well as corresponding documentation, parameterizations and configuration files for the software workflow necessary to generate the simulation data with OpenMalaria and reproduce the analysis are available at https://github.com/SwissTPH/TPP_workflow.

## Declarations

### Ethics approval and consent to participate

No human research participants or animals were involved in this study; therefore, ethics approval and consent to participate were not required.

### Consent for publication

Not applicable.

### Availability of data and materials

All analysis code used for this study, as well as corresponding documentation, parameterizations, and configuration files for the software workflow necessary to generate the simulation data with OpenMalaria and to reproduce this analysis are available in the TPP_workflow repository, https://github.com/SwissTPH/TPP_workflow.

### Authors’ contributions

M.A.P conceived the study. M.G., G.Y. and M.A.P designed the simulation experiments, developed the methodology, and analyzed the results. F.C., E.C., and N.C. provided methodological expertise. E.M.S., N.H., M.M., and S.R contributed expertise to product development, intervention properties, and guided analysis. M.G., G.Y., M.A.P. wrote the manuscript. All authors provided feedback and approved the final manuscript.

## Acknowledgements

We would like to thank Lydia Burgert, Theresa Reiker, Andrew Shattock, Thomas Smith, Sherrie Kelly, and Dylan Muir for insightful discussions and feedback on the developed methodology and the manuscript. We would also like to thank Thomas van Boeckel and Amalio Telenti for providing useful feedback on the manuscript. Calculations were performed at sciCORE (http://scicore.unibas.ch/) scientific computing core facility at University of Basel. We would further like to thank collaborators at the Innovative Vector Control Consortium (IVCC), PATH’s Malaria Vaccine Initiative (MVI), and Jörg Möhrle from Medicines for Malaria Venture (MMV) for their insightful discussions and feedback on the model scenarios. This work has been possible thanks to the Malaria Team at the Bill & Melinda Gates Foundation (BMGF) who facilitated exchanges with the product development partners and supported model scenarios and interpretations. In particular, we would like to thank Scott Miller, Jean-Luc Bodmer, Laura Norris, Bruno Moonen, Dan Strickman, and Philip Welkhoff from the BMGF Team.

## List of abbreviations

ATSB: attractive targeted sugar bait
BMGF: Bill and Melinda Gates Foundation
EIR: entomological inoculation rate
GP: Gaussian process
IPT: intermittent preventative treatment
IVCC: Innovative Vector Control Consortium
PATH-MVI: Program for Appropriate Technology in Health - Malaria Vaccine Initiative
PCR: polymerase chain reaction
*Pf*PR: *Plasmodium falciparum* prevalence of infections
R&D: research and development
RDT: rapid diagnostic test
SMC: seasonal malaria chemoprevention
TPP: Target Product Profiles
WHO: World Health Organization

## Additional file 1

### 1 Stakeholder engagement, role of TPP, model description, and interventions

#### 1.1 Stakeholder engagement

There was active engagement and regular communication exchanges with different expert groups during development of this methodological framework for guiding Target Product Profiles (TPPs) of novel malaria interventions. The aims of these exchanges were (1) to create a communication environment with stakeholders to be able to incorporate their feedback to ensure realistic representation of novel malaria interventions and of the simulated disease transmission settings in this approach, (2) to continuously shape this methodology to enable generating the relevant quantitative evidence to inform TPPs of novel malaria interventions from early stages of product development, and (3) to define and refine the priority research questions to be addressed with our analysis.

By offering a comprehensive snapshot of the development process at any given point in time, a TPP constitutes a vital reference for dialogue between various stakeholders to guide decisions on the development direction (35, 65–68). A well-constructed TPP is thus essential for efficient resource allocation and success during development (35, 68). However, the process of establishing TPPs relies on minimal clinical or quantitative evidence. They are often set by expert opinion and consensus is based on limited quantitative consideration of the complex dynamics of disease or on predictions of the likely intervention impact while achieving the identified unmet health needs (69). Furthermore, few malaria TPPs consider operational aspects such as deployment coverage in addition to product-specific characteristics such as efficacy or half-life. This has implications for the appropriate definition of intervention effectiveness characteristics according to local health systems and health targets (35, 69).

The stakeholders involved in these discussions were the Bill & Melinda Gates Foundation (BMGF), the Innovative Vector Control Consortium (IVCC), and the Program for Appropriate Technology in Health - Malaria Vaccine Initiative (PATH-MVI) and the WHO (7). Exchanges with these stakeholders were coordinated and guided by BMGF through regular meetings as follows. First, there was an initial convening with all stakeholders to frame the key study questions, to establish a network for iterative dialogue and to inform the partners about OpenMalaria, its components, features, and validity of assumptions. At these meetings, there were over 15 participants from BMGF, IVCC, PATH-MVI, and Swiss TPH. The health goal of malaria prevalence reduction in all ages was chosen during this meeting, as well as the five malaria interventions on which to focus our analysis. Second, several one-to-one iterative meetings were held with each stakeholder to define the way the five chosen interventions are implemented in OpenMalaria and to define relevant aspects to consider for their deployment (e.g., targets in the malaria transmission cycle, shape of decay, mass deployment, assessing impact at early and late follow-up), as well as putative ranges of their characteristics. We consulted with BMGF and PATH-MVI to refine the modeled vaccines, monoclonal antibodies, and drugs, and with IVCC to refine attractive targeted sugar baits and eave tubes. The set of simulated transmission settings (seasonality, indoor biting) were also defined during these meetings. Interim meetings were held with several stakeholders where we presented proof-of-concept and intermediate analyses. These meetings ensured building trust and a high level of confidence from our stakeholders in this analysis, while shaping the research questions. For example, an intermediate result was that the determinants of impact of novel interventions were different in low as compared with moderate transmission settings. This motivated exploring a wide range of *Pf*PR settings to ascertain how impact determinants were modulated as transmission increased. Another early result was that the efficacy, coverage, and half-life requirements of monoclonal antibodies and vaccines were significantly high, and that it was infeasible to reach high targets of prevalence reduction in high-transmission settings deploying these interventions alone. The subsequent discussions led to the exploration of deploying combinations of these interventions with an antimalarial drug and to refining target health goals based on the capabilities of the interventions to reduce burden. Results were presented during several stakeholder meetings and included in reports along with outlining prospects for future work.

Regular presentations of this ongoing analysis were provided to BMGF. These presentations covered relevant model assumptions for implementing different interventions, focusing on aspects where stakeholder feedback was needed, as well as intermediary results where model predictions for the interventions were shown. This ensured a dynamic iterative exchange where stakeholders were directly involved in shaping intervention characteristics, allowing them to see how their input was incorporated in this methodology (Table 1) and how these assumptions influenced the impact predictions of this analysis. These exchanges helped us design and refine the means of communication for this methodological approach and model predictions to a wider stakeholder audience. For this purpose, communication elements were used, such as tables summarizing the key quantitative results (e.g., Table 2), clear definitions of modeled intervention properties (see Additional file 1: section 1.2.3, and Table 1), schematic diagrams offering an intuitive representation for the methodological processes (Fig. 1 and 2), and a standardized system of plots showing results for the different interventions explored (Fig. 2-6). There were three types of standardized plots. First, sensitivity analysis plots (Fig. 2B and 4) displaying, for each intervention, the relative importance of the intervention characteristics in determining the impact on *Pf*PR_0-99_ reduction. Relative importance was calculated following global sensitivity analysis using Sobol’ indices (see Additional file 1: section 4.1 for a detailed description and calculations). These plots facilitated visual communication of key drivers of intervention impact and importantly how these drivers changed across various transmission settings, across immediate versus late follow-up of intervention impact, as well as across interventions. Second, landscapes of minimum intervention properties plot displaying, for different malaria transmission levels, an overview of the minimum intervention requirements in terms of coverage, efficacy, and half-life for a wide range of target *Pf*PR_0-99_ reductions (Fig. 2C - left panel, Fig. 5A, C, and E and 6A and C) (see Additional file 1: section 5.1 for a detailed description of the optimization process). These plots allowed for communication of overall intervention capabilities in reducing malaria prevalence, as well as their minimal requirements across different malaria transmission settings and for a wide range of *Pf*PR_0-99_ reduction targets. These requirements defined the profiles of intervention characteristics. Third, minimum profiles of properties plot displaying for different malaria transmission levels, the estimated minimum intervention requirements in terms of coverage, efficacy, and half-life for a chosen *Pf*PR_0-99_ reduction target (Fig. 2C – right, 5B, D, and F, and 6B and D) (see Additional file 1: section 5.1 for a detailed description of the optimization process). These plots facilitated the visual communication of the minimal intervention profiles and their variance across a range of different transmission settings for a chosen *Pf*PR_0-99_ reduction target. Furthermore, they allowed direct comparison of the required intervention profiles between immediate and long follow-up, across different deployment regimes (once versus twice per year), as well as across different intervention strategies (medical interventions deployed separately or in combination with an antimalarial drug).

#### 1.2 Description of the OpenMalaria model of malaria transmission dynamics

##### 1.2.1 Individual-based model of malaria transmission

OpenMalaria is an individual-based model of malaria transmission that considers the natural history of malaria in humans linked with a deterministic, entomological model of the mosquito oviposition cycle and malaria transmission in mosquitoes (70, 71) (Table S1.1). The modelled transmission cycle (Fig. 1) considers the chain of processes following infection of a human host, simulating malaria infection in individuals and modelling infection characteristics such as parasite density, duration of infection, infectivity to mosquitoes, and health outcomes such as anemia, other morbidities, or mortality. OpenMalaria specifically captures heterogeneity in host exposure, susceptibility, and immune response, taking into consideration the effects of several factors such as acquired immunity, human demography structure, and seasonality (72–75). Furthermore, the model includes a detailed representation of the health system (76), and a wide range of human and vector control interventions while tracking multiple health outcomes over time (Fig. 1 and Table S1.1).

In OpenMalaria, the pattern of yearly malaria infection in the absence of interventions is determined by the input entomological inoculation rate (EIR) (Additional file 1: Fig. S2.1). Each simulated infected human host has an associated parasite density and duration of infection (modelled individually and capturing disease effects such as immunity, infectiousness to mosquitoes, morbidity, or mortality). Setting-specific characteristics such as population demographics, mosquito species entomological characteristics or seasonality are explicitly modelled and a wide range of human and vector interventions can be applied, affecting the transmission cycle at various stages. Various health outcomes are monitored over time, including *Plasmodium falciparum* prevalence of infections (*Pf*PR), uncomplicated clinical disease, severe disease in and out of hospital, and malaria mortality. Disease dynamics model assumptions have been described and validated with field data in previous studies, notably in (39) and very recently revisited in (54). Several major consensus modeling studies have shown the validity of OpenMalaria predictions regarding the public health impact of the RTS,S vaccine (37), mass drug administration (41), or vector control (42). Additional validation studies conducted using published data on various malaria indicators such as EIR, *Pf*PR, incidence, and mortality have shown that OpenMalaria accurately captures the seasonally-dependent relationships between these indicators (43).

OpenMalaria has been widely documented and validated against a multitude of field studies, compared to existing models, and used in extensive studies to provide evidence for the epidemiological effects of various interventions (37, 39, 51, 58, 77, 78). It comprises 14 model variants based on distinct sets of assumptions of its epidemiology and transmission components (40). For the present analysis, the “base” simulation model was used. Mathematical equations of the OpenMalaria model, its assumptions, calibration, and validation have been thoroughly described in numerous publications (see the Background section), and are therefore not specified here, however, an overview of the key modelled disease processes and assumptions along with the corresponding references where model descriptions have been elaborated are provided in Table S1.1. In the sections below, an overview of the calibration and simulation settings used for this study has been provided.

**Table S1.1.**
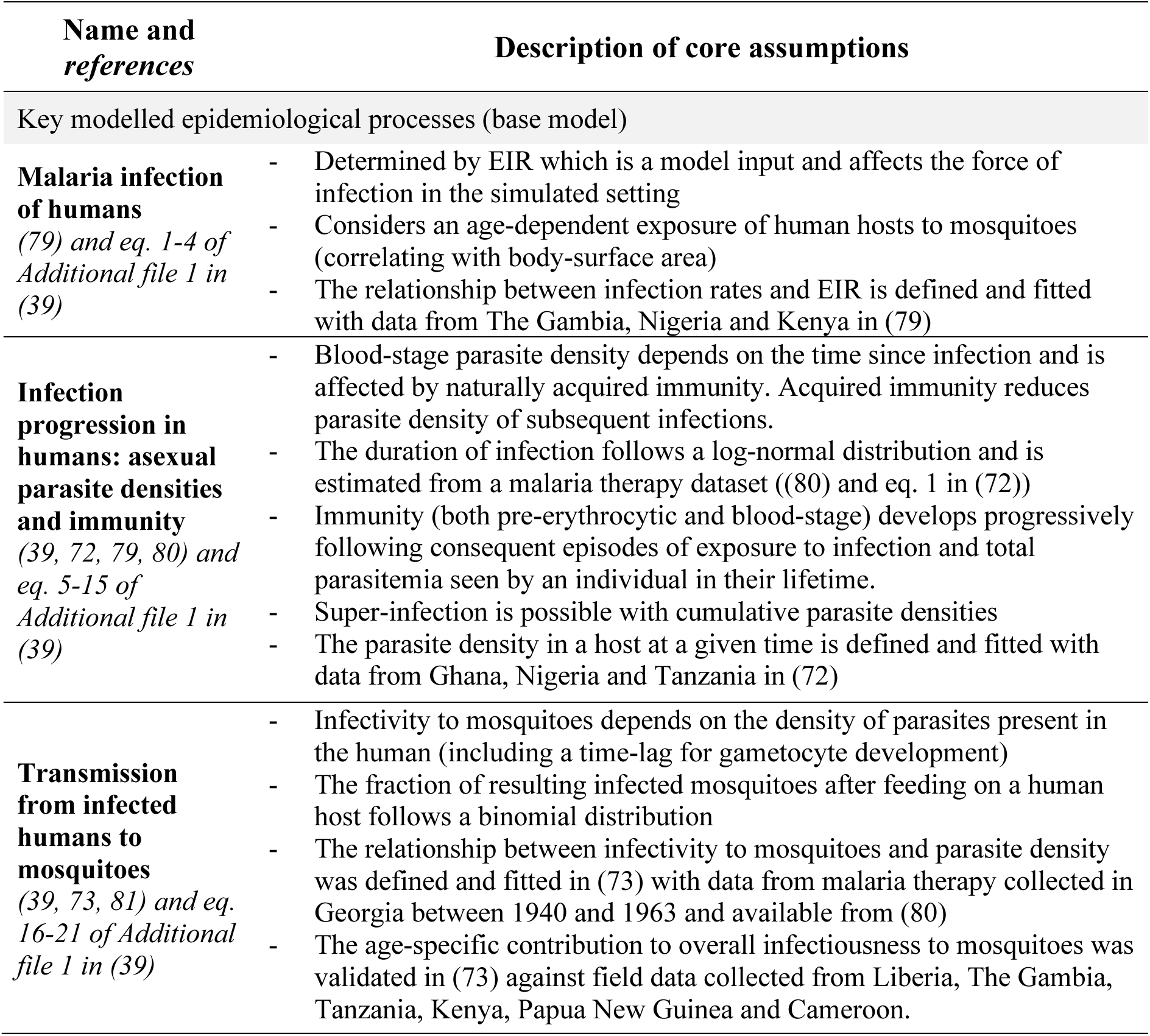
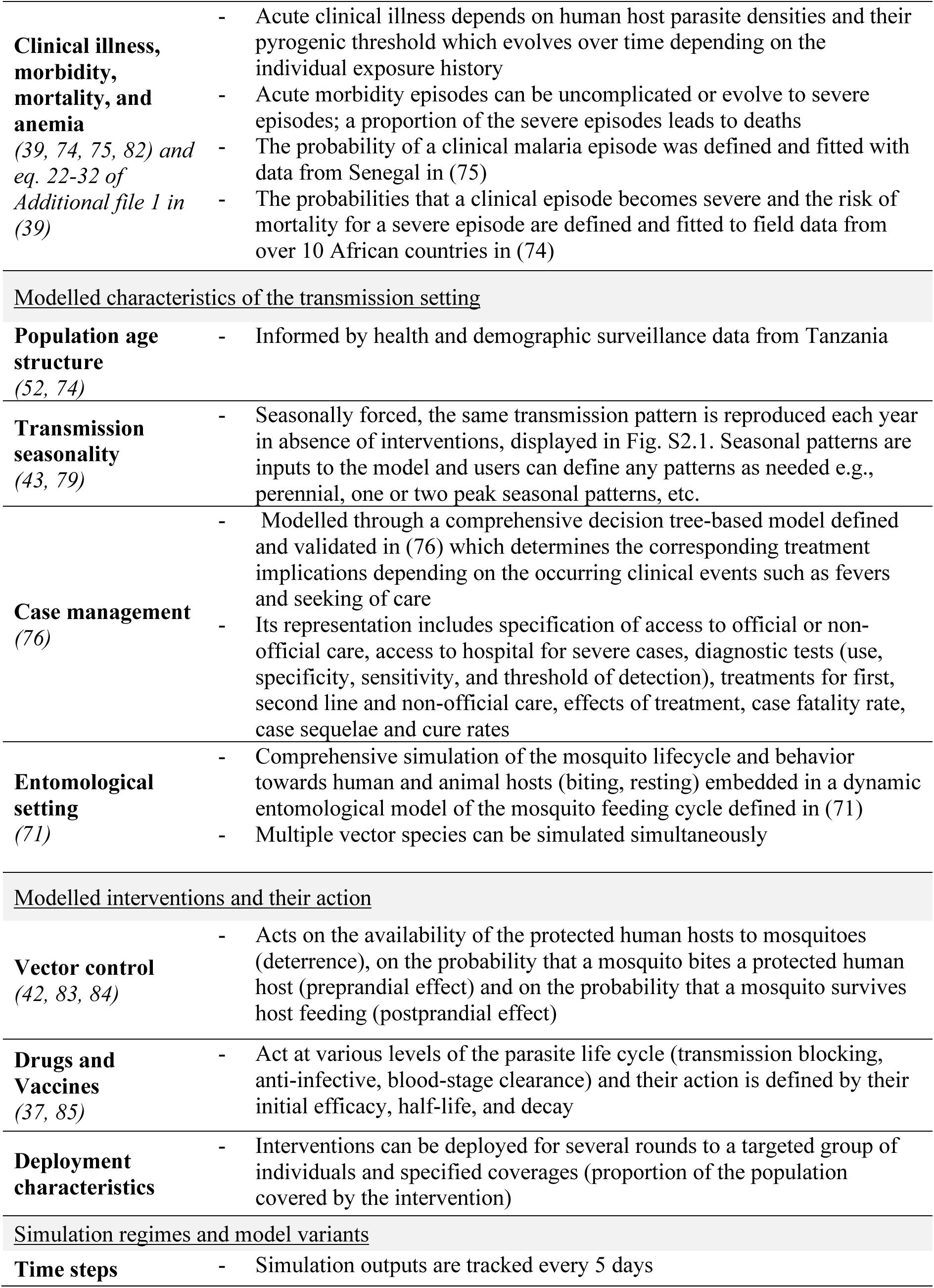
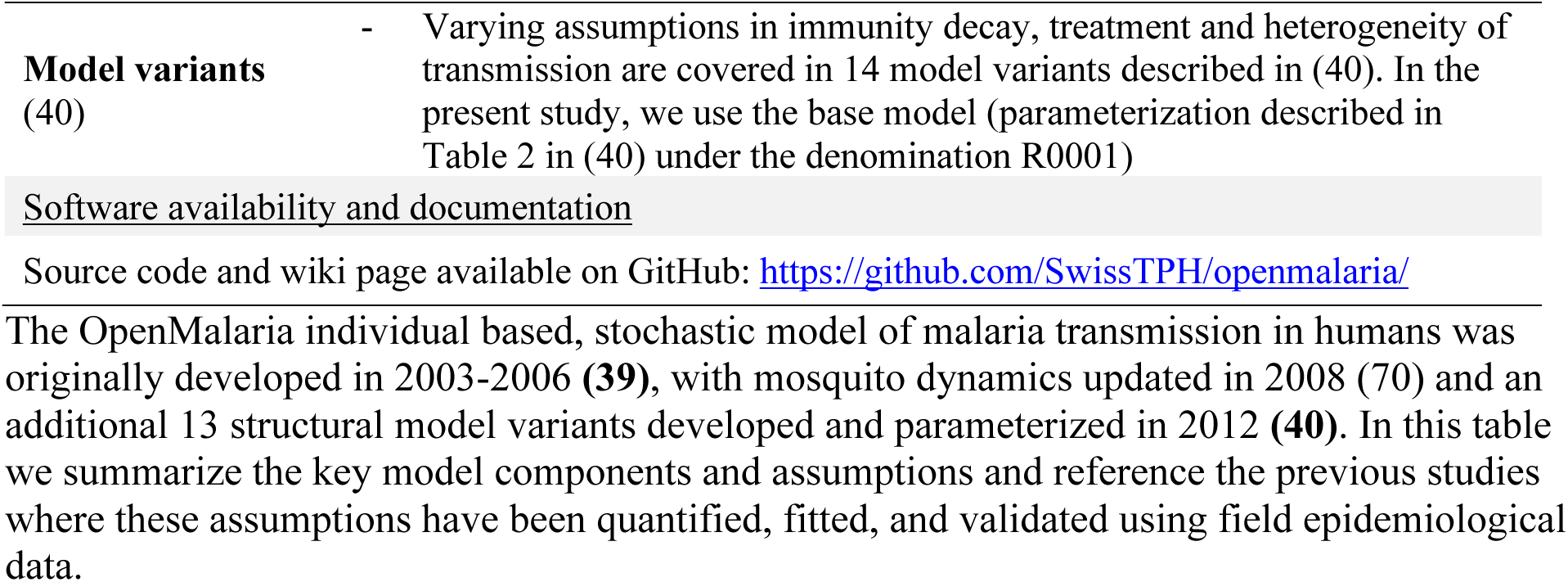
Overview of the OpenMalaria model components.

| Name and references | Description of core assumptions |
| --- | --- |
| Key modelled epidemiological processes (base model) |  |
| **Malaria infection of humans**<br>*(79) and eq. 1-4 of Additional file 1 in (39)* | - Determined by EIR which is a model input and affects the force of infection in the simulated setting<br>- Considers an age-dependent exposure of human hosts to mosquitoes (correlating with body-surface area)<br>- The relationship between infection rates and EIR is defined and fitted with data from The Gambia, Nigeria and Kenya in (79) |
| **Infection progression in humans: asexual parasite densities and immunity**<br>*(39, 72, 79, 80) and eq. 5-15 of Additional file 1 in (39)* | - Blood-stage parasite density depends on the time since infection and is affected by naturally acquired immunity. Acquired immunity reduces parasite density of subsequent infections.<br>- The duration of infection follows a log-normal distribution and is estimated from a malaria therapy dataset ((80) and eq. 1 in (72))<br>- Immunity (both pre-erythrocytic and blood-stage) develops progressively following consequent episodes of exposure to infection and total parasitemia seen by an individual in their lifetime.<br>- Super-infection is possible with cumulative parasite densities<br>- The parasite density in a host at a given time is defined and fitted with data from Ghana, Nigeria and Tanzania in (72) |
| **Transmission from infected humans to mosquitoes**<br>*(39, 73, 81) and eq. 16-21 of Additional file 1 in (39)* | - Infectivity to mosquitoes depends on the density of parasites present in the human (including a time-lag for gametocyte development)<br>- The fraction of resulting infected mosquitoes after feeding on a human host follows a binomial distribution<br>- The relationship between infectivity to mosquitoes and parasite density was defined and fitted in (73) with data from malaria therapy collected in Georgia between 1940 and 1963 and available from (80)<br>- The age-specific contribution to overall infectiousness to mosquitoes was validated in (73) against field data collected from Liberia, The Gambia, Tanzania, Kenya, Papua New Guinea and Cameroon. |
| **Clinical illness, morbidity, mortality, and anemia**<br>(39, 74, 75, 82) and eq. 22-32 of Additional file 1 in (39) | - Acute clinical illness depends on human host parasite densities and their pyrogenic threshold which evolves over time depending on the individual exposure history<br>- Acute morbidity episodes can be uncomplicated or evolve to severe episodes; a proportion of the severe episodes leads to deaths<br>- The probability of a clinical malaria episode was defined and fitted with data from Senegal in (75)<br>- The probabilities that a clinical episode becomes severe and the risk of mortality for a severe episode are defined and fitted to field data from over 10 African countries in (74) |
| <u>Modelled characteristics of the transmission setting</u> |  |
| **Population age structure**<br>(52, 74) | - Informed by health and demographic surveillance data from Tanzania |
| **Transmission seasonality**<br>(43, 79) | - Seasonally forced, the same transmission pattern is reproduced each year in absence of interventions, displayed in Fig. S2.1. Seasonal patterns are inputs to the model and users can define any patterns as needed e.g., perennial, one or two peak seasonal patterns, etc. |
| **Case management**<br>(76) | - Modelled through a comprehensive decision tree-based model defined and validated in (76) which determines the corresponding treatment implications depending on the occurring clinical events such as fevers and seeking of care<br>- Its representation includes specification of access to official or non-official care, access to hospital for severe cases, diagnostic tests (use, specificity, sensitivity, and threshold of detection), treatments for first, second line and non-official care, effects of treatment, case fatality rate, case sequelae and cure rates |
| **Entomological setting**<br>(71) | - Comprehensive simulation of the mosquito lifecycle and behavior towards human and animal hosts (biting, resting) embedded in a dynamic entomological model of the mosquito feeding cycle defined in (71)<br>- Multiple vector species can be simulated simultaneously |
| <u>Modelled interventions and their action</u> |  |
| **Vector control**<br>(42, 83, 84) | - Acts on the availability of the protected human hosts to mosquitoes (deterrence), on the probability that a mosquito bites a protected human host (preprandial effect) and on the probability that a mosquito survives host feeding (postprandial effect) |
| **Drugs and Vaccines**<br>(37, 85) | - Act at various levels of the parasite life cycle (transmission blocking, anti-infective, blood-stage clearance) and their action is defined by their initial efficacy, half-life, and decay |
| **Deployment characteristics** | - Interventions can be deployed for several rounds to a targeted group of individuals and specified coverages (proportion of the population covered by the intervention) |
| <u>Simulation regimes and model variants</u> |  |
| **Time steps** | - Simulation outputs are tracked every 5 days |
| **Model variants**<br>(40) | - Varying assumptions in immunity decay, treatment and heterogeneity of transmission are covered in 14 model variants described in (40). In the present study, we use the base model (parameterization described in Table 2 in (40) under the denomination R0001) |

The OpenMalaria individual based, stochastic model of malaria transmission in humans was originally developed in 2003-2006 **(39)**, with mosquito dynamics updated in 2008 (70) and an additional 13 structural model variants developed and parameterized in 2012 **(40)**. In this table we summarize the key model components and assumptions and reference the previous studies where these assumptions have been quantified, fitted, and validated using field epidemiological data.

**Table S2.2.** Estimated core parameters of OpenMalaria describing the natural history of the disease in human hosts.

| Parameter | Estimated value from (40)<br>(model R0001) |
| --- | --- |
| Lower limit of success probability of inoculations at high EIR | 0.049 |
| Critical value of EIR | 0.032 inoculations/person-night |
| Lower limit of success probability of inoculations in immune individuals | 0.14 |
| Critical value of cumulative number of entomological inoculations | 1'514.4 inoculations |
| Steepness of relationship between success of inoculation and the critical value of cumulative number of entomological inoculations | 2.04 |
| Variation between hosts on parasite densities (variance of log-normal distribution) | 10.174 |
| Critical value of cumulative number of parasite days | 3.52 parasite-days/$\mu\text{l} \times 10^{-7}$ |
| Critical value of cumulative number of infections | 97.3 infections |
| Maternal protection at birth | 0.90 |
| Decay of maternal protection | 2.53 per year |
| Fixed variance component for densities | 0.66 $[\ln(\text{parasites}/\mu\text{l})]^2$ |
| Critical value of cumulative number of infections for variance in parasite densities | 0.92 infections |
| Critical value in determining an increase of the pyrogenic threshold | 6'502.3 parasites/$\mu\text{L}$ |
| Factor determining an increase in the pyrogenic threshold | 143'000 parasites$^2\mu\text{l}^{-2}\text{day}^{-1}$ |
| Density bias (non Garki) | 0.177 |
| Mass action parameter | 1 |
| Case fatality for severe episodes in the community compared to hospital | 2.09 |
| Co-morbidity intercept relevant to indirect mortality | 0.019 |
| Non-malaria intercept for infant mortality rate | 49.5 deaths/1000 live births |
| Density bias (Garki) | 4.796 |
| Parasitemia threshold for severe episodes type B<sub>1</sub> | 784'000 parasites/μL |
| Immune penalty | 1 |
| Immune effector decay | 1 |
| Prevalence of co-morbidity/susceptibility at birth relevant to severe episodes | 0.092 |
| Decay rate of the pyrogenic threshold | 2.5 per year |
| Critical value of parasite density in determining an increase in the pyrogenic threshold | 0.6 parasites/μl |
| Asexual immunity decay | 0 |
| Pyrogenic threshold at birth | 296.302 parasites/μl |
| Idete multiplier | 2.798 |
| Critical age of co-morbidity | 0.117 |

**Table S1.3.** OpenMalaria model parameters describing the dynamics of the mosquito feeding cycle.

| Parameter | Value |
| --- | --- |
| Duration of the period when a mosquito is resting | 3 days |
| Extrinsic incubation period (time required for sporozoite development) | 11 days |
| Proportion of mosquitoes which are host seeking on same day as oviposition | 0.313 |
| Maximum proportion of the day that a mosquito spends host-seeking | 0.33 |
| Probability that a mosquito survives the feeding cycle | 0.623 |
| Probability that a mosquito successfully bites the chosen human host | 0.95 |
| Probability that a mosquito escapes the human host and finds a resting place after biting | 0.95 |
| Probability that a mosquito successfully rests after finding a resting site | 0.99 |
| Probability that a mosquito successfully lays eggs given that it has rested | 0.88 |
| Human blood index: the proportion of resting mosquitoes which fed on human blood during their last feed | 0.5 |

These parameters have been previously estimated in (70) and were kept unchanged during all the simulations in the present study.

##### 1.2.2 Calibration of the disease model and description of simulation experiments

The present analysis is based on a previously calibrated version of the model that reflects demographic, epidemiology, entomology, health system, and seasonality of a health facility catchment area in Tanzania (56, 74, 76). Core calibration parameters were previously estimated using a genetic algorithm approach, with optimization of a weighted sum over 10 objective functions (40). These objective functions represent key epidemiological relationships captured from available study survey and study site data as follows (along with figures displaying data and model fits from the reference studies): age patterns of incidence after interventions (Fig. 5 from (79)), age patterns of prevalence (Fig. 4 in (72)), age patterns of parasite density (Fig. 6 in (72)), age patterns of the multiplicity of infections (Fig. 5 in (72)), age patterns of clinical malaria incidence (Fig. 1 in (75)), age patterns of the parasite density threshold for clinical attacks (Fig. 4 in (75)), hospitalization rate in relation to prevalence in children (Fig. 2 in (74)), age patterns of hospitalization in relation to severe malaria (Fig. 4 in (74)), malaria specific mortality in children less than 5-years-old (Fig. 7 and 8 in (74)), and indirect malaria infant mortality rate (Fig. 9 in (74)). Full calibration procedure details have been described in (40) and (54), and the estimated parameters have been previously summarized in Table 1 in (39), in Table 3 in (40), and in Additional file 1: Table S1 in (54). A summary of the model parameters has been provided in Additional file 1: Tables S1.2 and S1.3.

As summarized in the Methods section, the simulated human population size in this analysis was 10,000 individuals, with its age structure informed by data collected from a health and demographic surveillance site in Ifakara, Tanzania, available through the INDEPTH network (52). For all simulations, it is assumed there were no imported infections during the entire study period.

Health system characteristics (Additional file 1: Table S1.1) were defined through parameterization of a case management model based on data provided by the Tanzanian National Malaria Control Program (76). To define the simulated case management level, the probability of effective cure within two weeks from the onset of fever (E_14_) was varied within the interval [0-0.8] corresponding to a probability of seeking care (access to treatment) within 5 days from the onset of fever (E_5_) within the interval [0.04-0.5] (77). During model simulations, the case management level was held constant over time.

Mosquito entomological parameters and seasonal exposure patterns were estimated from field studies conducted in the Namawala and Michenga villages located nearby Ifakara in Tanzania (86, 87). Two archetypal seasonal settings were simulated: a seasonal exposure setting with one transmission peak in September estimated from the mentioned field studies (Fig. S2.1), and a perennial setting with uniform, constant exposure throughout the year. Two mosquito species were present in the simulated settings: endophagic (indoor-biting, human blood index of 0.99) and exophagic (outdoor-biting, human blood index of 0.5), respectively. The ratios between the population sizes of indoor and outdoor mosquito species were classified into three levels corresponding to high (indoor proportion of 0.8 of total mosquito population), mid (indoor proportion of 0.5) and low biting (indoor proportion of 0.2). The extent of malaria transmission in each simulation was defined by the annual entomological inoculation rate (EIR). For each simulation, EIR was sampled from the interval [1-25] leading to a simulated range of *Plasmodium falciparum* parasite rate or prevalence (*Pf*PR) distributions across the various transmission settings (Additional file 1: Fig. S2.1 and S2.3, and Table S2.1).

##### 1.2.3 Definition of intervention profiles

As summarized in the Methods section, a standardized representation for each malaria intervention was built. Accordingly, a malaria intervention was characterized through the targets of the transmission life cycle it affects, along with the efficacy, half-life, and decay of its effect (Fig. 1, Additional file 1: Fig. S2.2, and Table 1). The efficacy of a therapeutic or immunologic intervention was quantified by its ability to clear parasites or to prevent infection, while for mosquito-targeted interventions (vector control) it corresponded to the ability of the intervention to kill or to prevent mosquitoes from biting humans. For each intervention, its efficacy decayed over time according to a specific decay type (defined in Fig. S2.2). Intervention coverage was quantified by the percentage of the population affected by the respective intervention. Where interventions were applied to individual humans they were equally applied across ages, and not targeted to certain populations. Geographical setting characteristics such as entomological inoculation rates (EIR), seasonality, case-management coverage, as well as transmission and vector characteristics were also included in the simulation specifications (Fig. 1, Table 1).

The following intervention targets were defined in the transmission cycle (Fig. 1): “anti-infective” as acting at the liver stage and preventing occurrence of a new infection, “blood stage clearance” as clearing blood-stage parasites by administration of a drug, “transmission blocking” as preventing parasite development into gametocytes, “mosquito life-cycle killing effect” as killing mosquitoes during different stages of their life cycle, for example, before a blood meal (pre-prandial killing) and/or after a blood meal (post-prandial killing). Furthermore, mosquitoes are affected by vector control interventions according to their indoor and outdoor biting patterns.

The length of the intervention effect was described by half-life for exponential, sigmoidal, or biphasic decay profiles, or by duration for step-like decay profiles. Generally, half-life refers to the half-life of intervention efficacy decay, representing the time in which the initial intervention efficacy has been reduced by 50% (Additional file 1: Fig. S2.2, Table 1). The full duration of this effect is equivalent to the entire decay time. For simplicity, since only one intervention had a step-like decay, we used the words half-life and duration interchangeably.

Each intervention or combination of interventions was applied as mass intervention targeting to all ages equally, along with continuous case management. As part of development of this approach, targeting of particular populations or age groups was not examined. Mass intervention packages were deployed for a long model warm-up period (150 years) and were then implemented in June and/or December for three years (Fig. 3, Additional file 1: Fig. S3.1). Coverage at deployment time refers to the percentage of the population covered by the intervention’s initial efficacy, irrespective of how many doses/applications are required to reach that coverage, assuming that the necessary doses have previously occurred. The base scenario XML files used for these simulations can be found at https://github.com/SwissTPH/TPP_workflow/tree/master/Intervention_scenarios.

##### 1.2.4 Translation of input EIR to *Pf*PR_2-10_ and *Pf*PR_0-99_

For each simulation, OpenMalaria requires the definition of the intensity and seasonality of malaria exposure specified through the input EIR level and its yearly profile in the absence of interventions (Additional file 1: Fig. S2.1). EIR is an appropriate measure for reflecting transmission intensity (88), however it is difficult to measure in the field and its interpretation in the context of intervention impact is difficult to capture when looking at the effects of drugs and vaccines (89, 90). For this reason, although EIR is the force of infection input for all OpenMalaria simulations, simulation outcomes and downstream analyses at the corresponding median *Pf*PR_2-10_ were reported before interventions were deployed. True infection prevalence was reported and not patent PCR or RDT-detected. To do so, the continuous EIR space was discretized into unit-wide intervals and the median *Pf*PR was calculated across the obtained *Pf*PR for all simulations at each discrete interval (Additional file 1: Fig. S2.3).

##### 1.2.5 Definition of intervention impact and health goals

A comprehensive set of simulated scenarios was built by uniformly sampling the parameter space of setting and intervention characteristics. To estimate the impact of the deployed interventions, in each simulation, the reduction in *Pf*PR_0-99_ attributable to the deployed intervention was calculated. *Pf*PR_0-99_ reduction was calculated by comparing the initial average prevalence in the year before any interventions were deployed to the average annual prevalence obtained in the first (short follow-up) and third year (long follow-up) after interventions deployment (Fig. 3 and Additional file 1: Fig. S3.1). Consequently, the defined health goals corresponded to a given minimum threshold of *Pf*PR_0-99_ reduction that the deployed interventions should achieve.

Additional file 1: Figures S3.2-S3.4 present the distributions of obtained *Pf*PR_0-99_ reduction for the OpenMalaria simulation experiments covering all the interventions and deployments investigated in the present study. In seasonal, low-transmission settings (EIR < 2) a high proportion of simulations reached elimination before any intervention was deployed and were removed from the analysis (Additional file 1: Fig. S3.5). Since this happened for over 75% of simulations at EIR < 2, we did not investigate optimal intervention profiles for transmission settings with EIR < 2. Arguably, for settings close to elimination, a different health goal, such as the probability of elimination, would be more appropriate which is outside the scope of this study, which focuses on reducing *Pf*PR_0-99_.

### 2 Disease scenarios

**Fig. S2.1.**
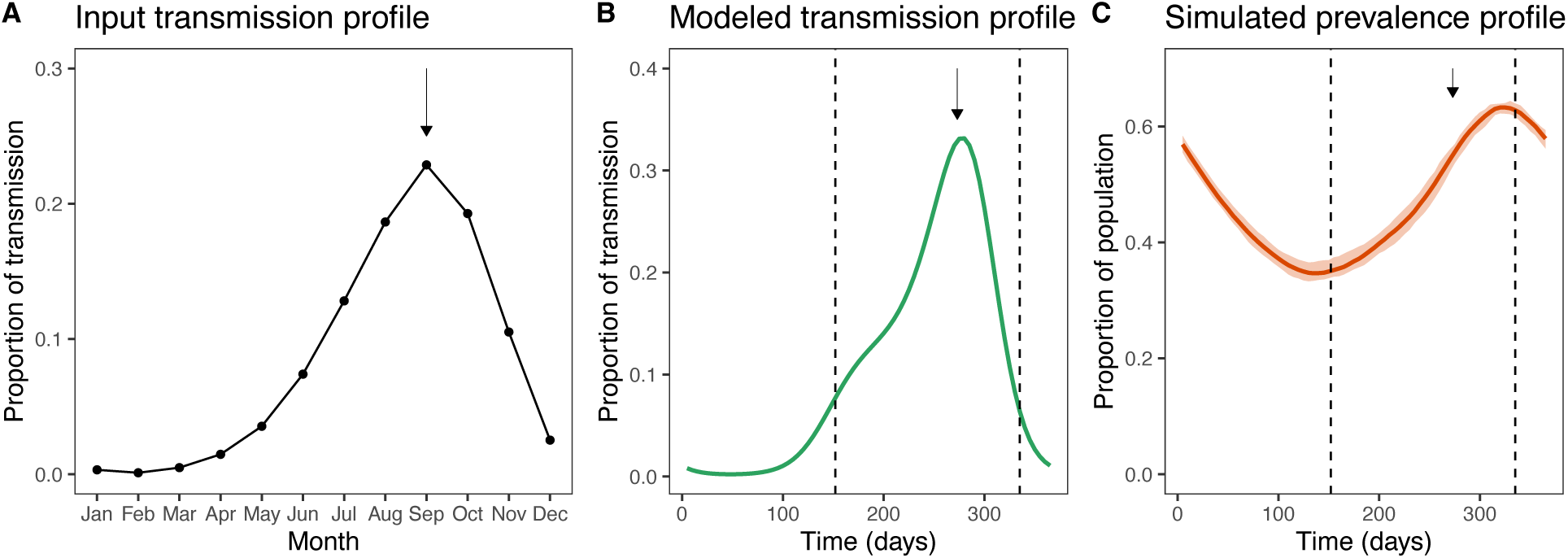
Illustration of the yearly malaria transmission and prevalence patterns in simulated seasonal settings. (A) Observed, normalized, monthly seasonal pattern of malaria EIR in Namawala, Tanzania extracted from (74). (B) Corresponding input, 5-day seasonal EIR pattern used in OpenMalaria simulations, obtained by scaling and extrapolating the monthly seasonality profile from (74) to 5- day time steps. For this example, the simulated input EIR was 7.78 infectious bites per person per year. (C) Resulting simulated yearly *Pf*PR_0-99_ profile. In all figures, the arrows indicate the month of September, the peak of transmission and show the delay between the peak of transmission and the resulting peak in malaria prevalence. The dotted vertical lines on figures (B) and (C) indicate the deployment times of first and second rounds of malaria interventions when applicable.

**Fig. S2.2.**
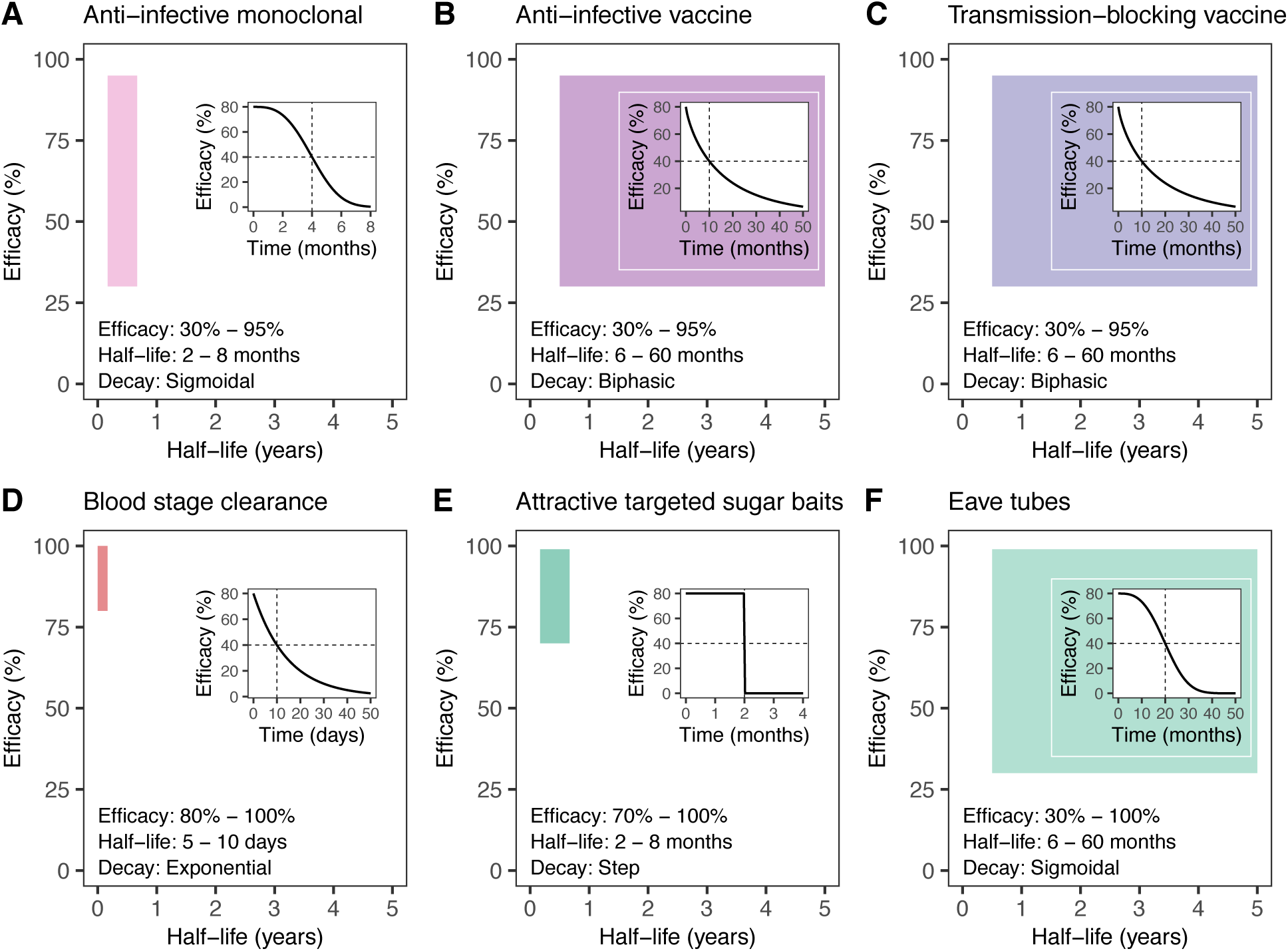
Representation of decay and the range of efficacy and half-life against different parasite or vector targets for intervention-agnostic malaria interventions. The simulated malaria interventions (A–F) were modeled in terms of their targets in the malaria transmission cycle. The effect of each intervention is represented through the half-life of its decay (x-axis) as well as the initial efficacy (y-axis). The color blocks represent the range of parameter space of efficacy and half-life of decay considered in the current analysis for each intervention. The half-life and the color block do not represent the entire duration of effect, as that depends on the decay shape chosen for each intervention. The decay shape for each intervention is displayed in the right side insert of each plot where the dotted lines specify the half-life and corresponding half of the intervention efficacy. The definitions of all the parameter ranges for all interventions are provided in each figure on the lower left side and detailed in Table S2.1.

**Fig. S2.3.**
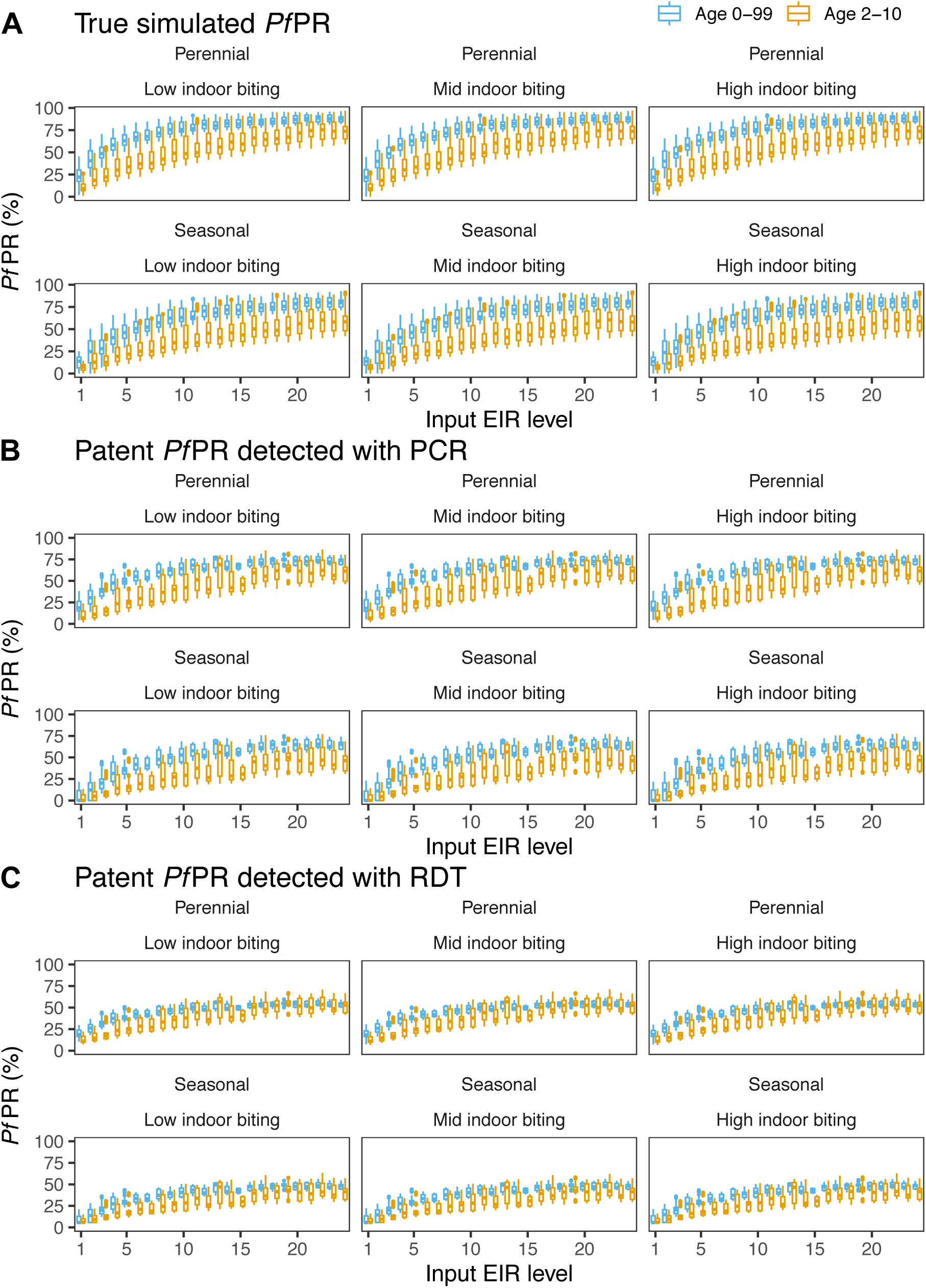
Simulated distributions of true and patent (detected with PCR or RDT) *Pf*PR_0-99_ and *Pf*PR_2-10_ for various input EIR levels in absence of interventions. The input entomological inoculation rate (EIR) defines the simulated malaria transmission level. In every simulation experiment, EIR was uniformly sampled from the interval [1, 25]. In figures (A)–(C), each panel corresponds to a simulated setting and presents the distributions of true (A), patent with PCR (B) and patent with RDT (C) *Plasmodium falciparum* prevalence (*Pf*PR, shown with boxplots, blue for 0-99-year-old and orange for 2-10-year-old) at varying EIR levels (x-axis). The 6 represented settings are defined by the seasonality pattern (perennial shown in the first row, or seasonal shown in the second row of each figure) and mosquito indoor biting behavior (low-shown in the first column, mid-shown in the second column or high-indoor biting shown in the third column of each figure). Each EIR level on the x-axis is defined as a set of continuous input EIR values which range between the current level and the current level-1, e.g., an input EIR level of 1 contains EIR values in the interval (0, 1]. For each EIR level and setting, the case management levels, i.e., the probability of seeking care (access to treatment) within 5 days from the onset of fever (E_5_), was varied within the interval [0.04-0.5]. PCR: polymerase chain reaction. RDT: rapid diagnostic test.

**Table S2.1.**
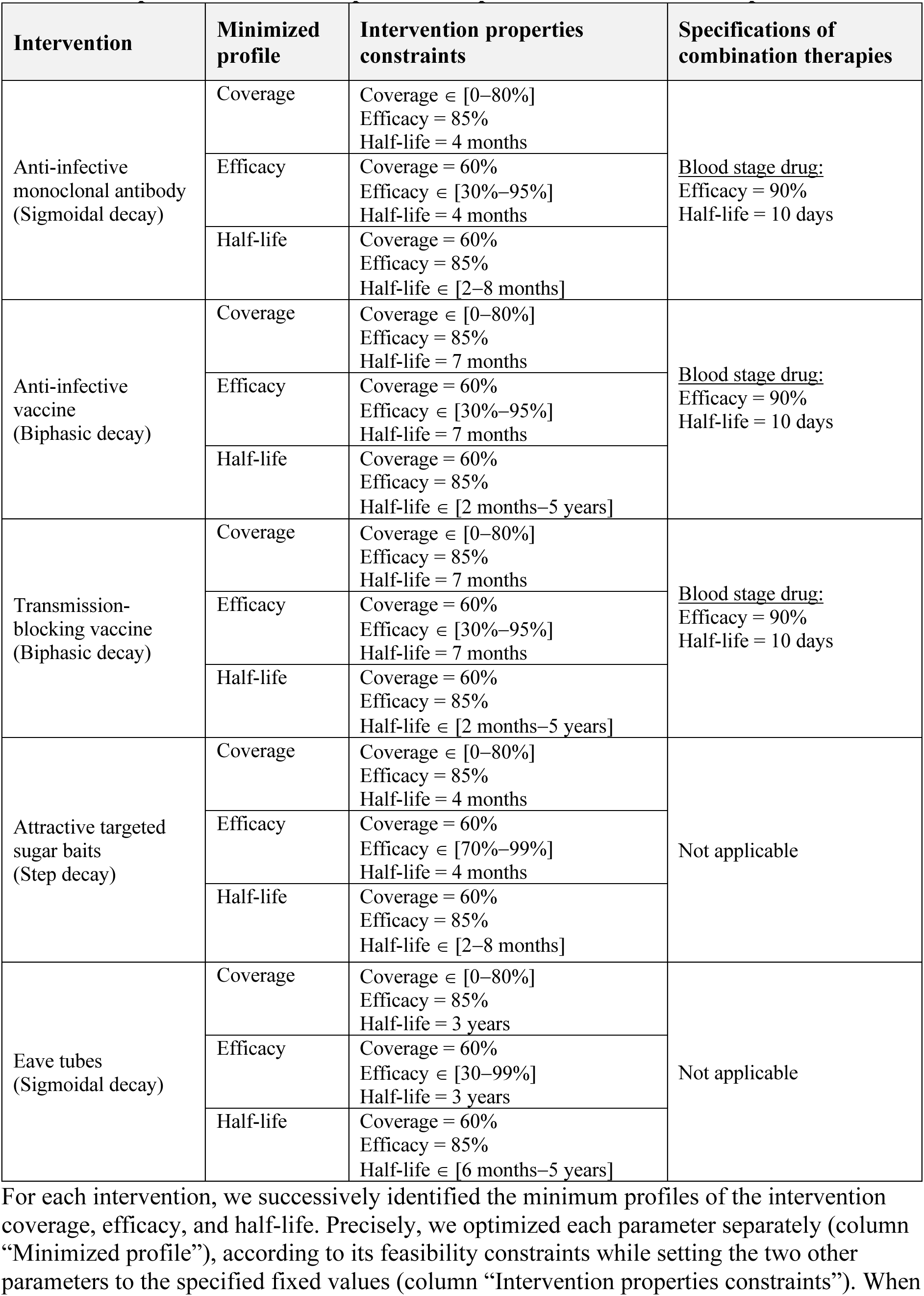
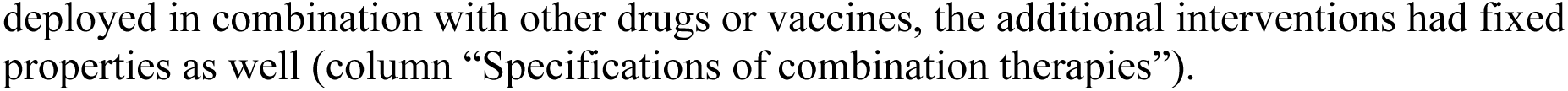
Specifications of the optimization procedure for TPP development.

#### 2.1 Building a disease model emulator with Gaussian processes

As stated in the Methods, since it was computationally intensive and challenging to run an exhaustive number of simulations in order to explore the entire parameter space for diverse combinations of interventions, settings, and deployments using OpenMalaria, machine learning techniques and kernel methods were applied. Precisely, starting from a training dataset of simulations generated with OpenMalaria, Gaussian process (GP) models (50) were used to infer the relationship between simulation variables (e.g., intervention coverage, half-life, efficacy, etc.) and corresponding intervention impact (*Pf*PR_0-99_ reduction). This approach allowed a fast, simplified predictive model that could provide estimates of the disease model output for any new inputs to be built without running new OpenMalaria model simulations.

Gaussian process models are non-parametric models which define a prior probability distribution over a collection of functions using a kernel, smoothing function. Precisely, given the relationship

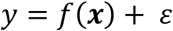

where *y* is the *Pf*PR_0-99_ reduction here, and ***x*** represents the set of intervention parameters *x_1_*, …, *x_n_*, the main assumption of a GP is that

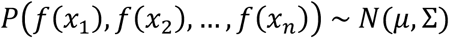

where

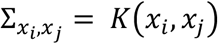

is the covariance matrix of the Gaussian distribution, μ is its mean, and *K* is a kernel function (50). Once data are observed, the posterior probability distribution of the functions consistent with the observed data can be derived, which is then used to infer outcomes at unobserved locations in the parameter space (50). The intuition behind a GP model is based on the “smoothness” relationship between its components. Accordingly, points which are close in the input parameter space will lead to close points in the output space.

#### 2.2 Training data

For each intervention and setting, a training dataset was built using discrete Latin hypercube uniform sampling (91) across the input parameter space (defined in Table 1). This sampling scheme ensured uniform coverage of the parameter space and a representative set of points spanning the variability of the predicted output across the entire space. Ten stochastic realizations (replicates) of each sampled data point were considered. OpenMalaria was run on the sampled data and *Pf*PR_0-99_ was calculated for short and long follow-up. The size of the training set was varied between 10 and 1,000 points (100-10,000 including replicates) for several simulation experiments (Additional file 1: Fig. S4.1) and the performance of the trained GP was assessed via the Pearson correlation coefficient *r^2^*. The minimum training set size which led to r^2^ > 0.95 was selected for the remaining simulation experiments.

#### 2.3 Gaussian process emulators

For each transmission setting and intervention, a GP model with a Gaussian kernel was trained for a 5-fold cross-validation scheme using the training dataset with OpenMalaria simulations. For training the GP, the R package HetGP version 1.1.1 was used (92, 93). HetGP is a powerful implementation of GP models, featuring heteroskedastic GP modeling embedded in a fast and efficient maximum-likelihood-based inference scheme.

GP performance was assessed by calculating the correlation between true and predicted outputs on out-of-sample test sets, as well as the mean squared error (Additional file 1: Fig. S4.2 and S4.3 and Table S4.1). Precisely, the training set was split into 5 subsets and, iteratively, 4 of these subsets were used for training the GP, while the remaining set was used as an out-of-sample test set during the cross-validation procedure. After assessing the prediction error obtained during the cross-validation procedure, the GP was trained using the entire training set.

Furthermore, since the trained GP model provides the mean and variance for each predicted output, this probabilistic representation was used to assess the uncertainty of the trained model across the entire parameter space and to refine the GP model through adaptive sampling (94–96). Accordingly, new training points from high-uncertainty regions of the parameter space were iteratively sampled and the model was updated with the new training samples. Precisely, 100,000 points were sampled using Latin hypercube sampling and the variance of the predicted output was evaluated with the previously trained GP emulator. Samples from “rare” regions of the parameter space containing fewer outputs were prioritized. To do so, the output *Pf*PR_0-99_ range was classified into 4 bins and the numbers of predicted outcomes corresponding to the 100,000 sampled points that fell in each bin (output density) calculated. Of the sampled points, 30 points (300 including replicates) were chosen proportional to the output density in each bin and with the highest predicted variance. This procedure was repeated five times to ensure that the correlation between true and predicted values on an out-of-sample test set had reached a plateau. Finally, a separate out-of-sample test set was built to assess the overall performance of the GP (Additional file 1: Fig. S4.2, S4.3, and Table S4.1).

### 3 Results: Disease model simulation

**Fig. S3.1.**
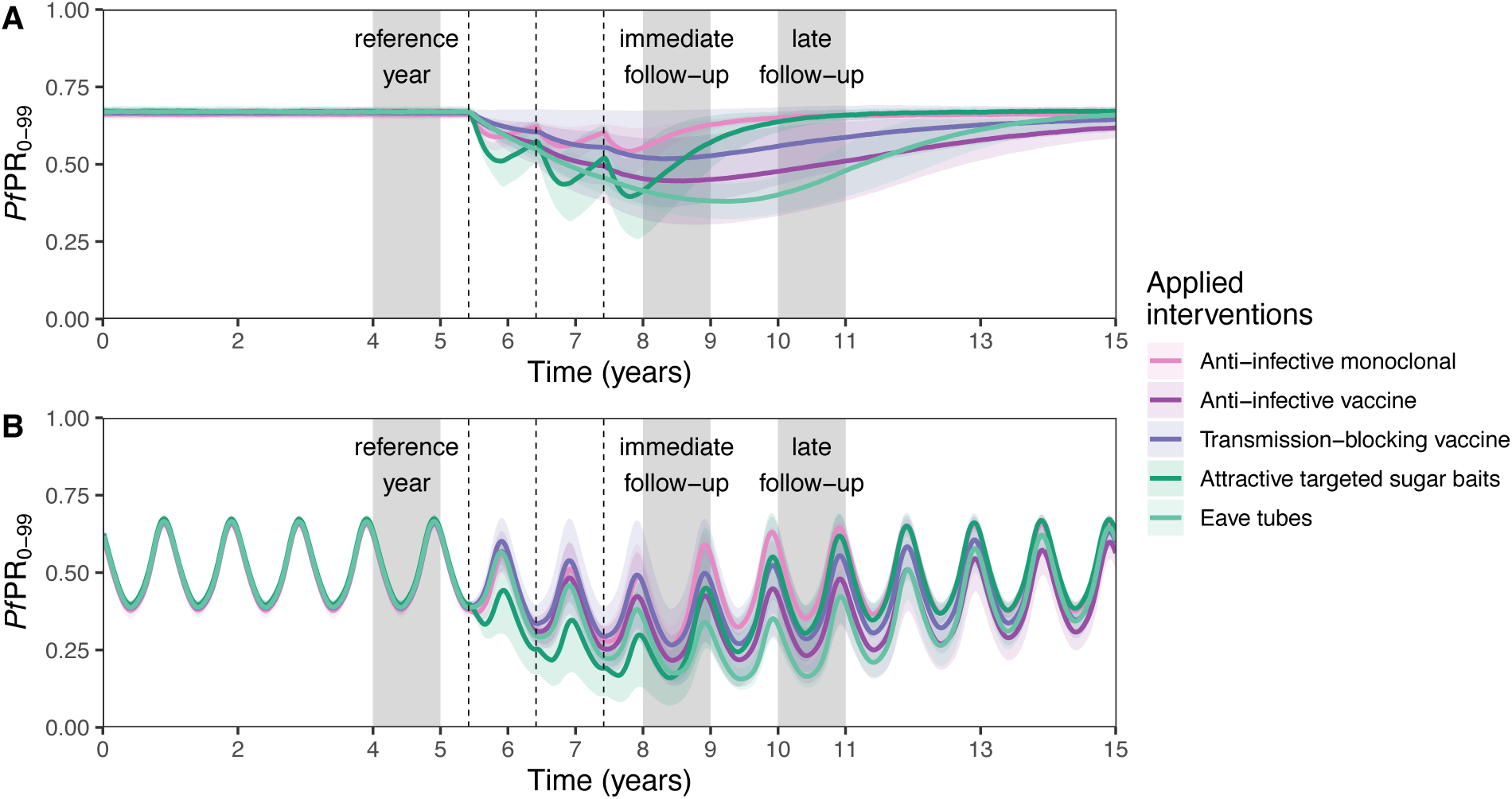
Examples of OpenMalaria simulation outputs. Time series of simulated malaria *Pf*PR_0-99_ in a perennial (A) and seasonal (B) setting. Both figures display the prevalence of malaria cases, *Pf*PR_0-99_, (y-axis) across time (x-axis). Interventions targeting different stages in the malaria transmission cycle (different colors) are applied once per year at the beginning of June (vertical dotted lines, in this example for three years of deployment). The effect of each intervention is assessed by evaluating the *Pf*PR_0-99_ reduction in all ages relative to the year prior deployment (first grey block). Two outcomes are assessed, following an immediate and late follow-up (second and third grey blocks), depending on whether the average prevalence is calculated across the next year after deployment, or across the third year after deployment, respectively.

**Fig. S3.2.**
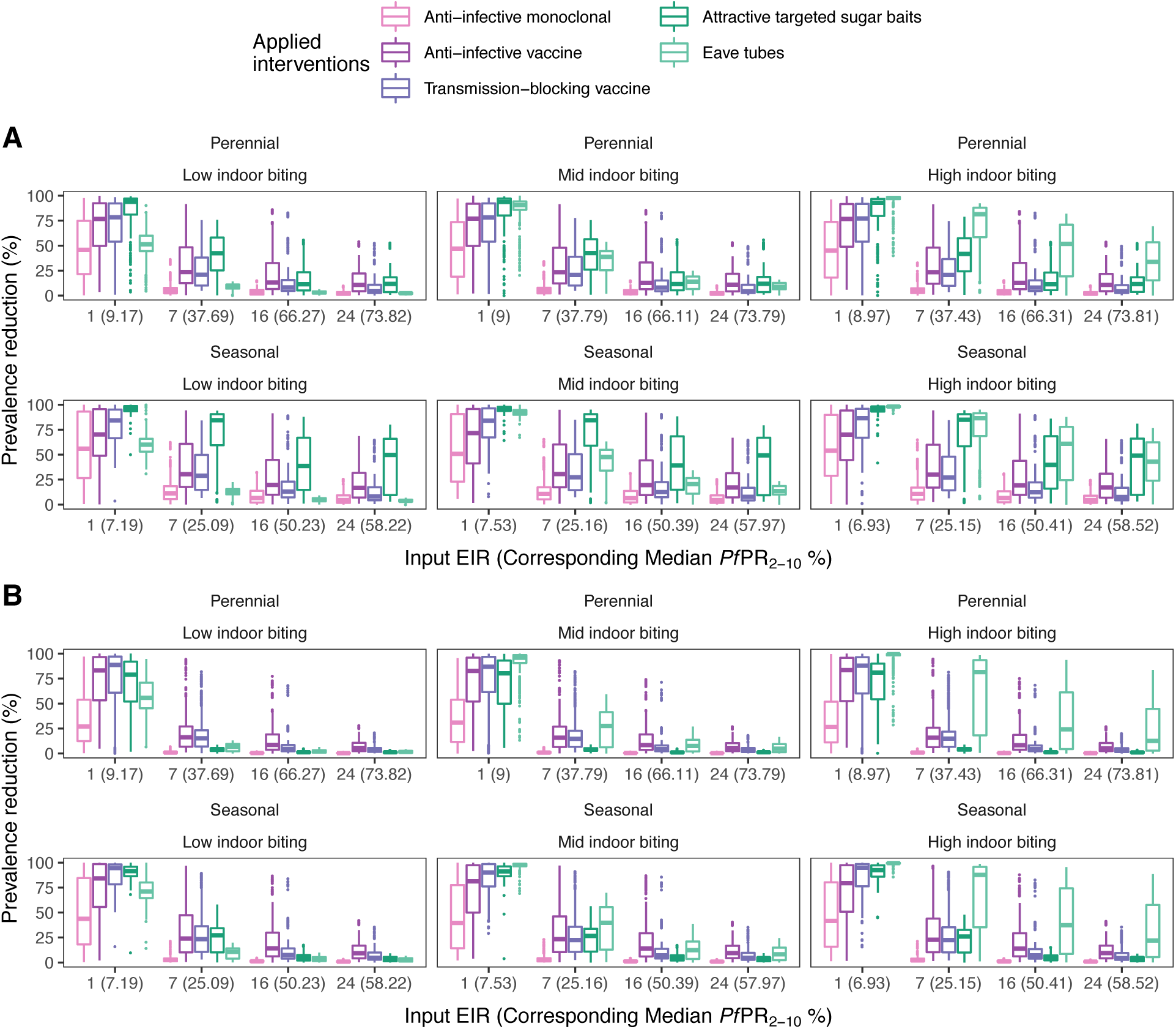
Distributions of prevalence reduction following yearly deployment of single interventions. Prevalence reduction was calculated by comparing the initial prevalence in the year before any interventions were deployed to the yearly prevalence obtained in the following year (short follow-up, panel A) and in the third year (long follow-up, panel B) after deployment of interventions. Each individual figure corresponds to a simulated setting and presents the distributions of *Pf*PR_0-99_ reduction (shown with boxplots) at varying EIR as well as corresponding simulated *Pf*PR_2-10_ levels (x-axis). Each boxplot displays the interquartile range (box), the median value (horizontal line), the largest and smallest values within 1.5 times the interquartile range (whiskers), and the remaining outside values (points). The 6 represented settings in each panel are defined by the seasonality pattern (perennial or seasonal) and mosquito indoor biting behavior (low, mid, or high indoor biting). Each EIR level on the x-axis is defined as a set of continuous input EIR values which range between the current level and the current level-1, e.g., an input EIR level of 1 contains all EIR values in the interval (0, 1]. The definitions and ranges of all the EIR levels is included in Table S2.1.

**Fig. S3.3.**
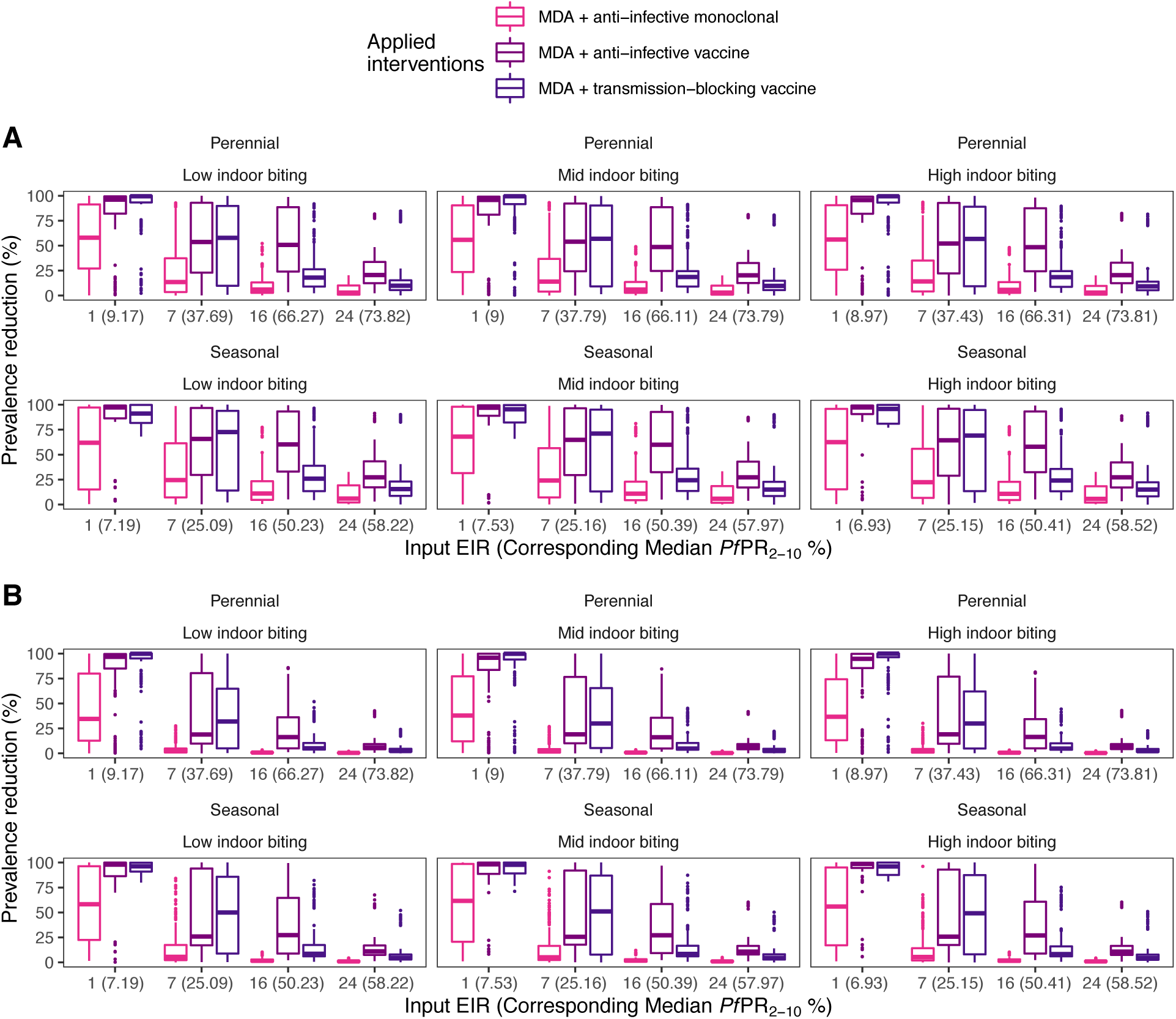
Distributions of prevalence reduction following yearly deployment of combinations of interventions. Prevalence reduction was calculated by comparing the initial prevalence in the year before any interventions were deployed to the yearly prevalence obtained in the following year (short follow-up, panel A) and in the third year (long follow-up, panel B) after deployment of interventions. Each individual figure corresponds to a simulated setting and presents the distributions of *Pf*PR_0-99_ reduction (shown with boxplots) at varying EIR as well as the corresponding simulated *Pf*PR_2-10_ levels (x-axis). Each boxplot displays the interquartile range (box), the median value (horizontal line), the largest and smallest values within 1.5 times the interquartile range (whiskers), and the remaining outside values (points). The 6 represented settings in each panel are defined by the seasonality pattern (perennial or seasonal) and mosquito indoor biting behavior (low, mid, or high indoor biting). Each EIR level on the x-axis is defined as a set of continuous input EIR values which range between the current level and the current level-1, e.g., an input EIR level of 1 contains EIR values in the interval (0, 1]. The definitions and ranges of all the EIR levels are included in Table S2.1. MDA: mass drug administration.

**Fig. S3.4.**
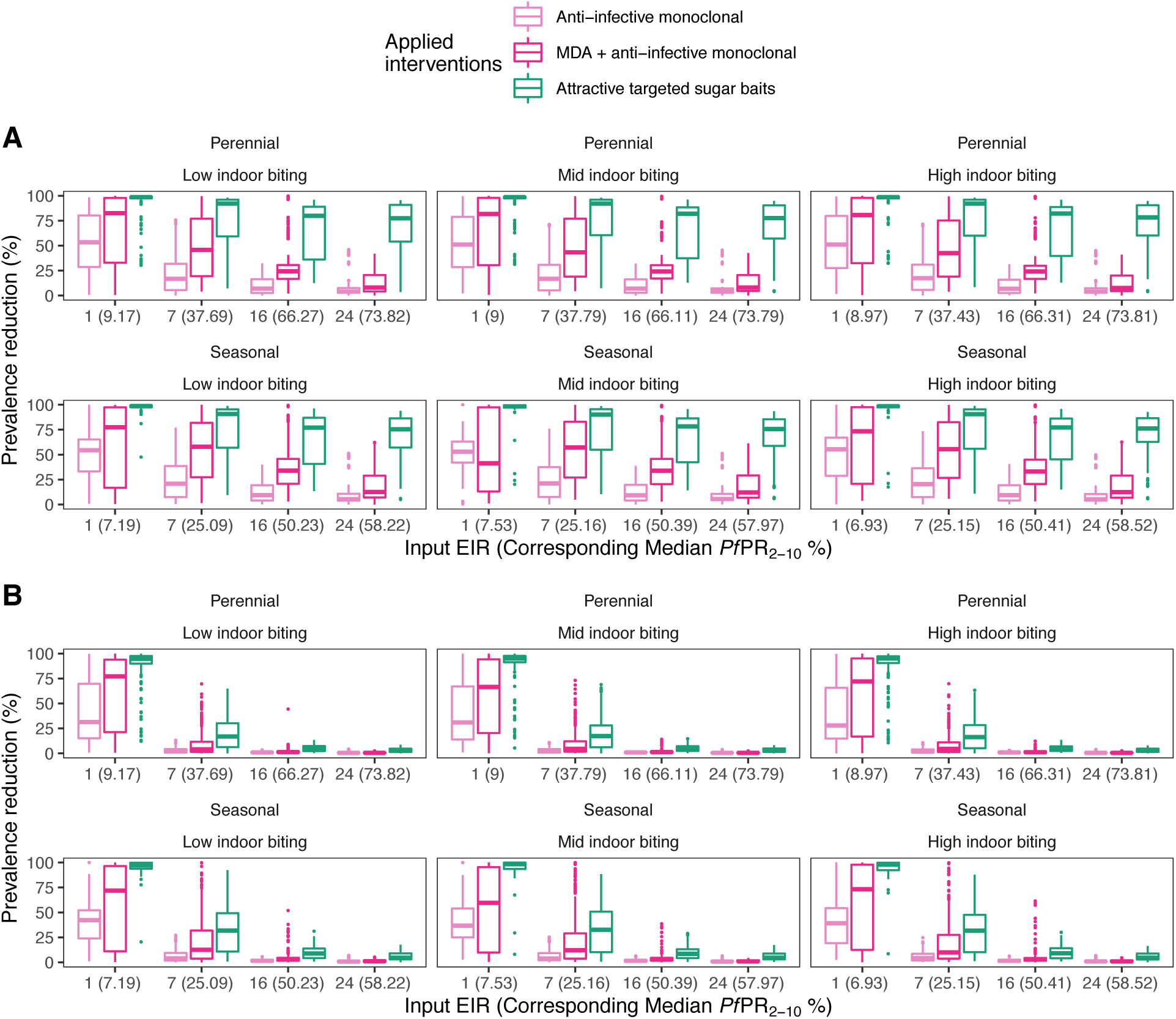
Distributions of prevalence reduction following deployment of single and combinations of interventions twice per year. Prevalence reduction was calculated by comparing the initial prevalence in the year before any interventions were deployed to the yearly prevalence obtained in the following year (short follow-up, panel A) and in the third year (long follow-up, panel B) after deployment of interventions. Each individual figure corresponds to a simulated setting and presents the distributions of *Pf*PR_0-99_ reduction (shown with boxplots) at varying EIR as well as corresponding simulated *Pf*PR_2-10_ levels (x-axis). Each boxplot displays the interquartile range (box), the median value (horizontal line), the largest and smallest values within 1.5 times the interquartile range (whiskers), and the remaining outside values (points). The 6 represented settings in each panel are defined by the seasonality pattern (perennial or seasonal) and mosquito indoor biting behavior (low, mid, or high indoor biting). Each EIR level on the x-axis is defined as a set of continuous input EIR values which range between the current level and the current level-1, e.g., an input EIR level of 1 contains EIR values in the interval (0, 1]. The definitions and ranges of all the EIR levels for all simulated settings is included in Table S2.1. MDA: mass drug administration.

**Fig. S3.5.**
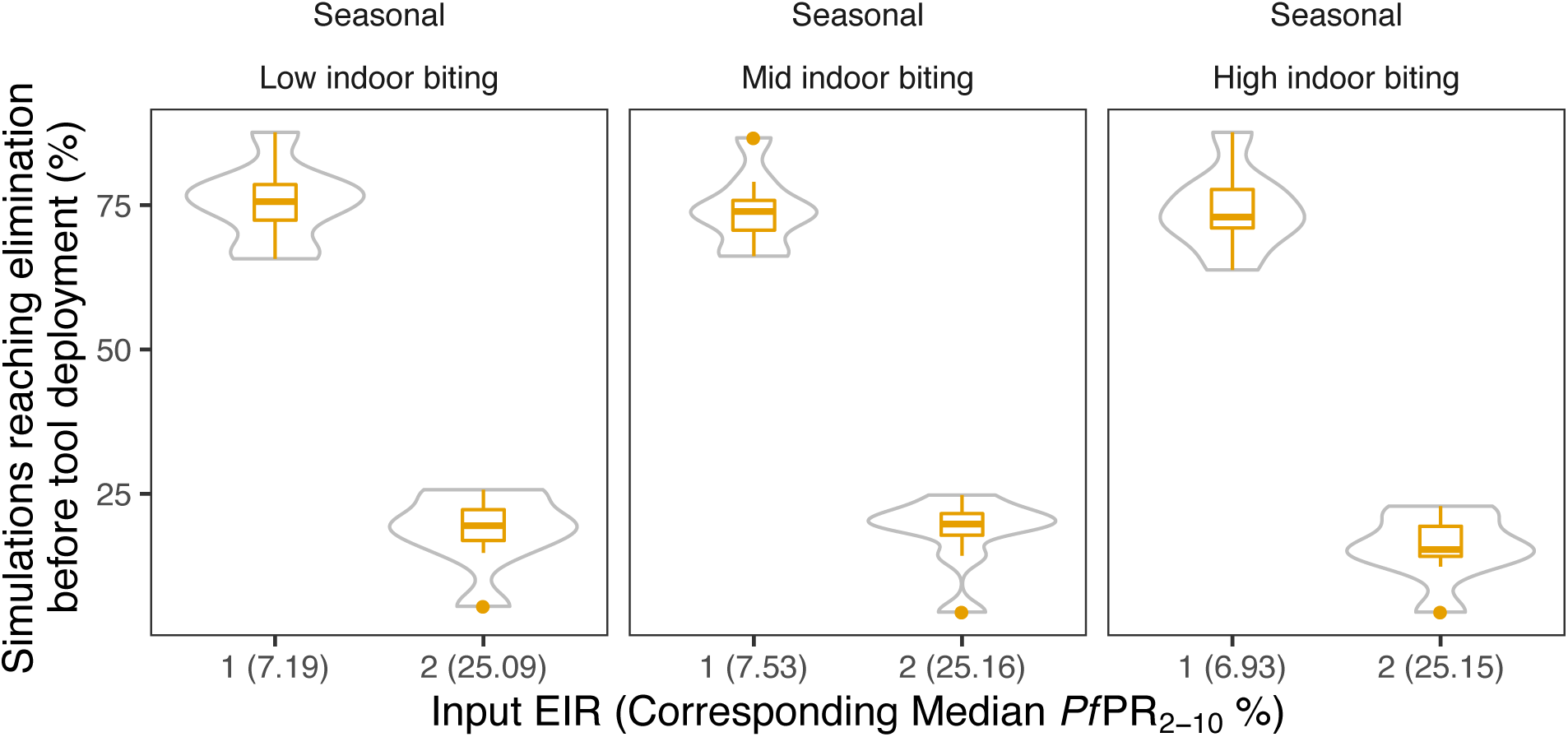
Simulations reaching malaria elimination before intervention deployment. The violin plots and boxplots in each panel present the distributions of the percentage of simulations reaching malaria elimination (*Pf*PR_0-99_ = 0) before intervention deployment (this can arrive due to case management and only occurs in seasonal settings), across all simulated interventions and intervention combinations.

### 4 Sensitivity analysis and emulator performance results

#### 4.1 Identifying impact determinants through sensitivity analysis

To estimate the contribution of each model input and its interactions with the other inputs to the variance of the model outcome, a global sensitivity analysis based on variance decomposition (55) was conducted. This analysis shows which input parameters have higher impact on the model outcome. It relies on the decomposition of the output variance in a sum of individual input parameter conditional variances:

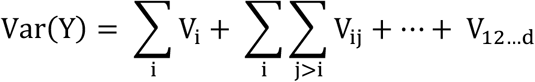

where *Y* is the model outcome (in this case, *Pf*PR_0-99_ reduction), *d* is the number of model inputs, and the conditional variances defined as:

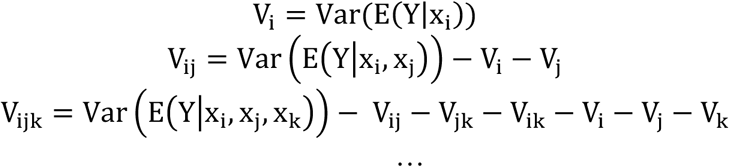

with *x_1_*, …, *x_n_* representing the model input parameters.

Based on the above decomposition of output variance, the first order sensitivity index is defined as:

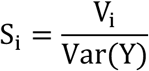

and corresponds to the proportion of output variance assigned to the main effect of *X_i_*, i.e., regardless of its interactions with other model inputs (55, 97).

To account for the contribution of each model input, as well as the variance of its interactions with other inputs to the variability of the model output, the total effect sensitivity index is used:

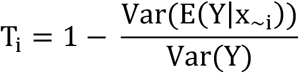

where ∼*i* stands for all indices except *i* (55, 97).

In the above decomposition of model output variance, by replacing the expressions of the sensitivity indexes, the following properties can be deduced:

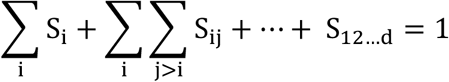

and

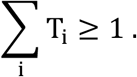

To compute the sensitivity indexes, the function *soboljansen* from the R package *sensitivity* (98) was used. The function estimates the sensitivity indices through MCMC sampling, using a Monte Carlo approximation for computing conditional expectations. Within the sampling scheme, 100,000 points to estimate the sensitivity indices were sampled.

Calculating the sensitivity indices defined above, the variance of the GP emulator output was thus decomposed into proportions attributable to intervention characteristics, i.e., intervention efficacy, half-life, and deployment coverage, as well as access to care. Using the main effects, the relative importance *r_i_* of each characteristic as a proxy for impact determinants was defined as follows:

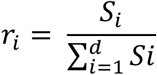

where *d* is the number of intervention characteristics and 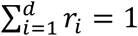.

#### 4.2 Results: Sensitivity analysis and emulator performance

**Fig. S4.1.**
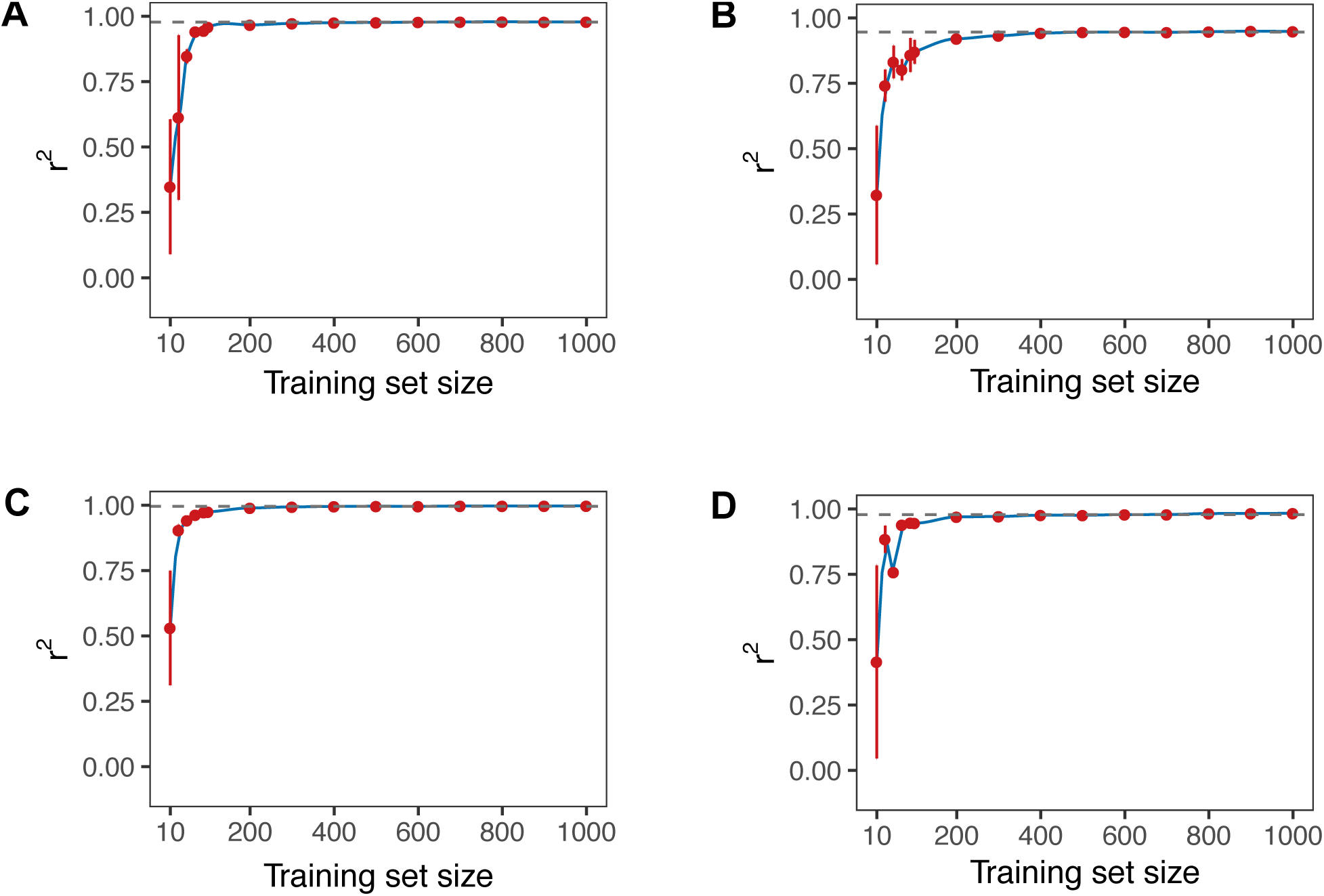
Assessment of the performance of the trained GP depending on the training set size. Each figure presents the Pearson correlation coefficient r^2^ between true and predicted values on a broad range of out-of-sample test sets of varying length, when simulating deployment of an anti-infective monoclonal antibody deployed once per year (A) or twice per year (B) as well as in combination with a blood-stage drug once (C) or twice per year (D).

**Fig. S4.2.**
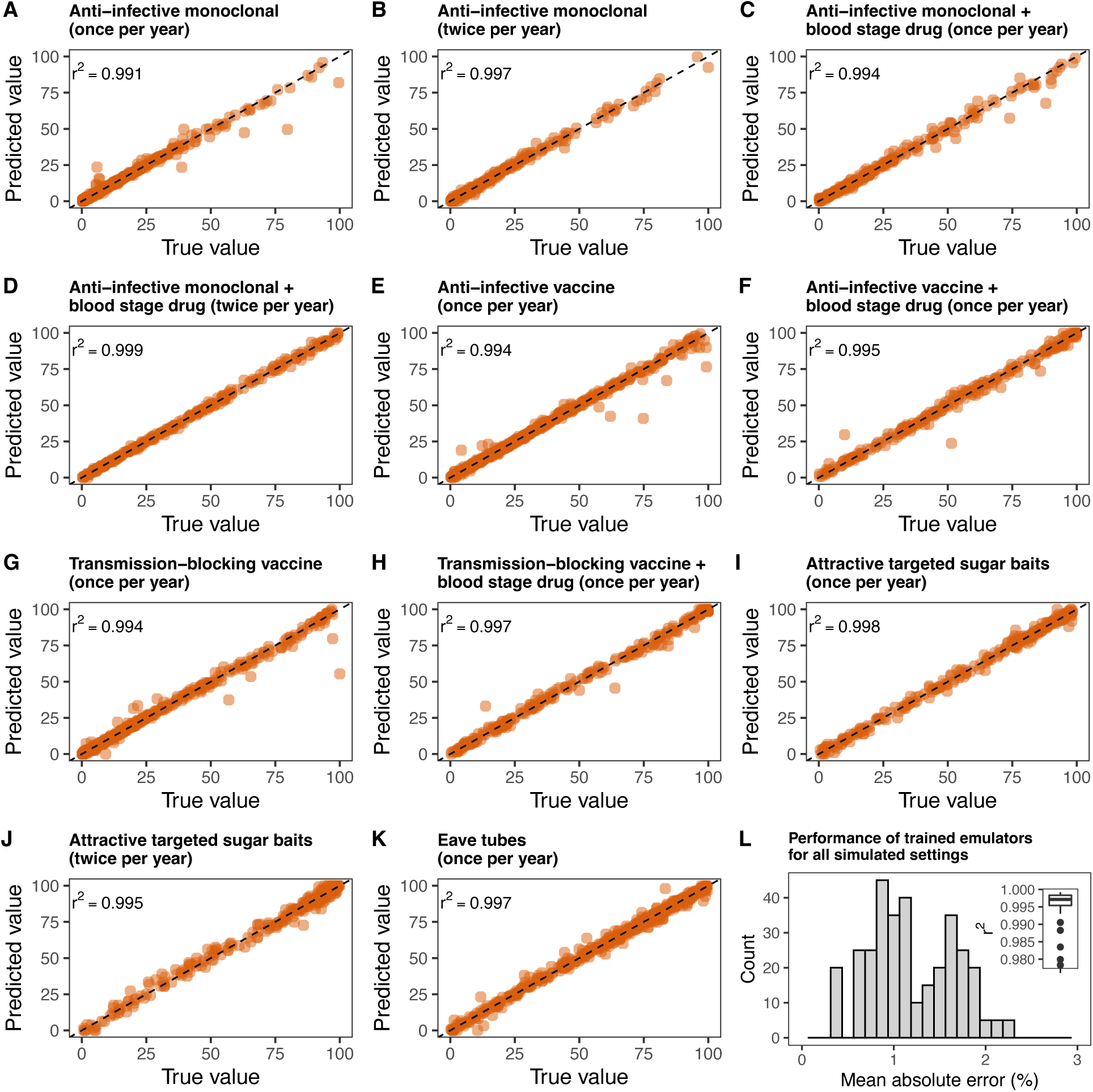
Performance of the trained GP emulators predicting immediate intervention impact. For a wide range of deployed interventions and transmission settings (see Methods), GP emulators were trained to predict the immediate impact of each intervention, i.e., the resulting average *Pf*PR_0-99_ reduction in the year following deployment of the intervention. The performance of the trained emulators was assessed by inspecting the Pearson correlation coefficient (r^2^) and the mean absolute error between true and predicted values on an out-of-sample test set. Figures (A) – (K) display the true and predicted values of each trained emulator across all deployed interventions in a seasonal transmission setting with high indoor biting. Figure (L) summarizes r^2^ and the mean absolute error of all the trained emulators for all simulated transmission settings and interventions (the simulated settings were defined by seasonality and mosquito biting patterns, see Table S2.1 for detailed values per setting).

**Fig. S4.3.**
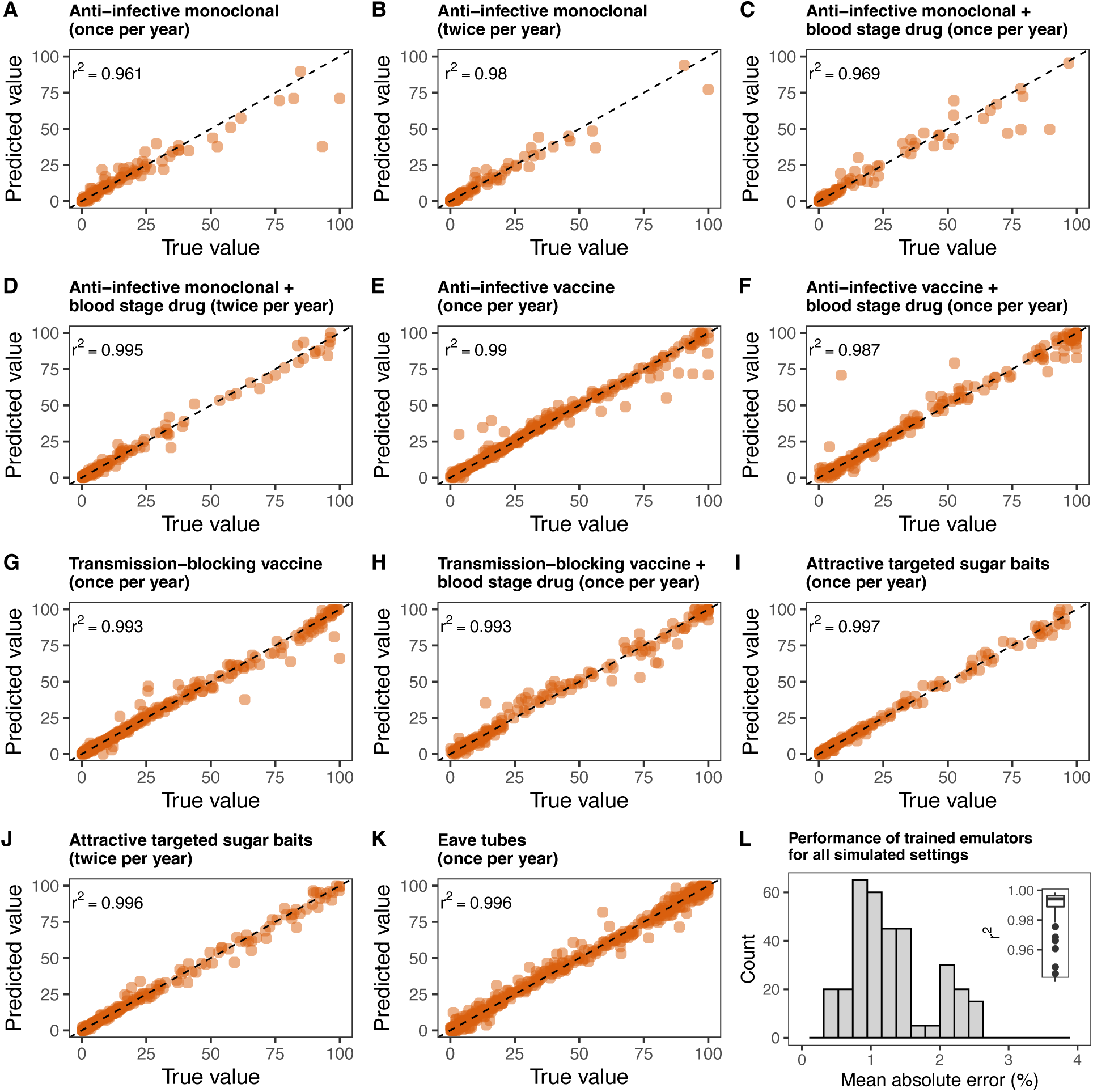
Performance of the trained GP emulators predicting long-term intervention impact. For a wide range of deployed interventions and transmission settings (see Methods section), GP emulators were trained to predict the immediate impact of each intervention, i.e., the resulting average *Pf*PR_0-99_ reduction in the third year following deployment of the intervention. The performance of the trained emulators was assessed by inspecting the Pearson correlation coefficient (r^2^) and the mean absolute error between true and predicted values on an out-of-sample test set. Figures (A) – (K) display the true and predicted values of each trained emulator across all deployed interventions in a seasonal transmission setting with high indoor biting. Figure (L) summarizes r^2^ and the mean absolute error of all the trained emulators for all simulated transmission settings and interventions (the simulated settings were defined by seasonality and mosquito biting patterns, see Table S2.1 for detailed values per setting).

**Fig. S4.4.**
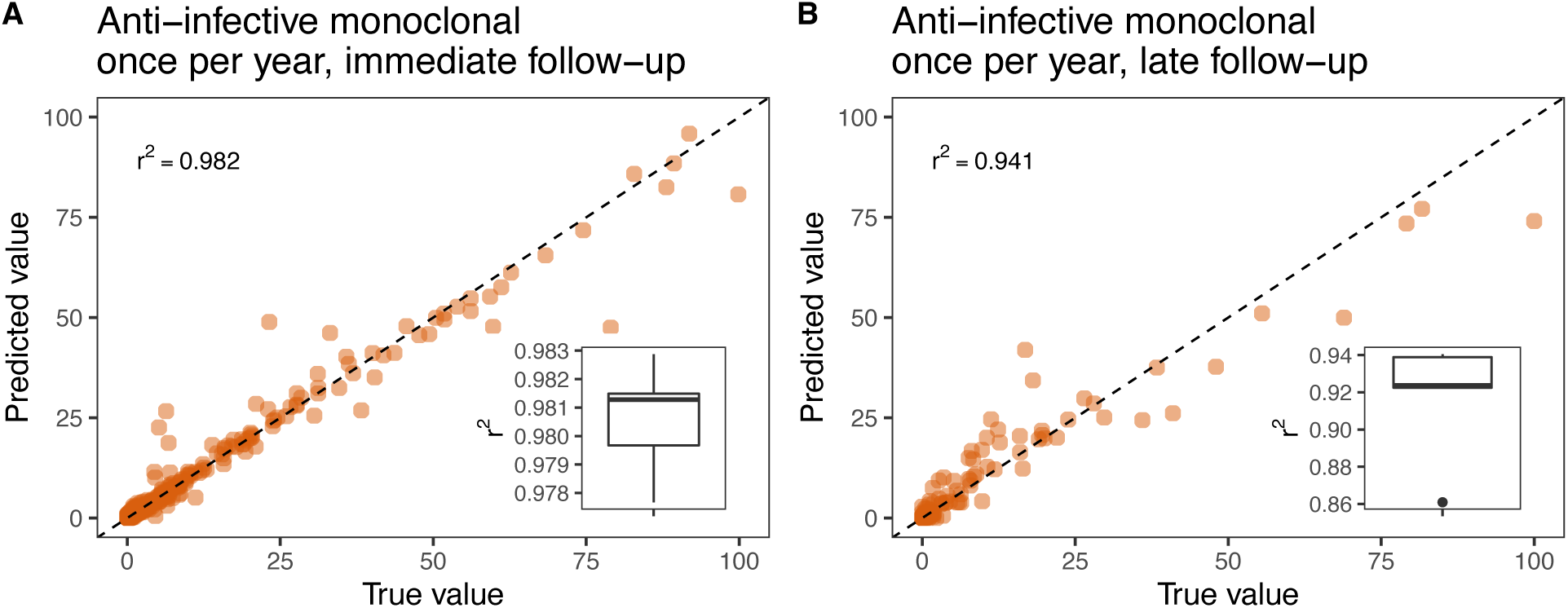
Performance of trained GP emulators predicting incidence reduction. *Plasmodium falciparum* malaria incidence reduction in all ages in the first (A) and third (B) year following deployment of a monoclonal antibody intervention. The performance of the trained emulators was assessed by inspecting the Pearson correlation coefficient (r^2^) in a cross-validation scheme (lower right boxplot displays the distribution of r^2^ obtained on the left-out test sets during cross-validation) as well as on an out-of-sample test set (upper left corner).

**Fig. S4.5.**
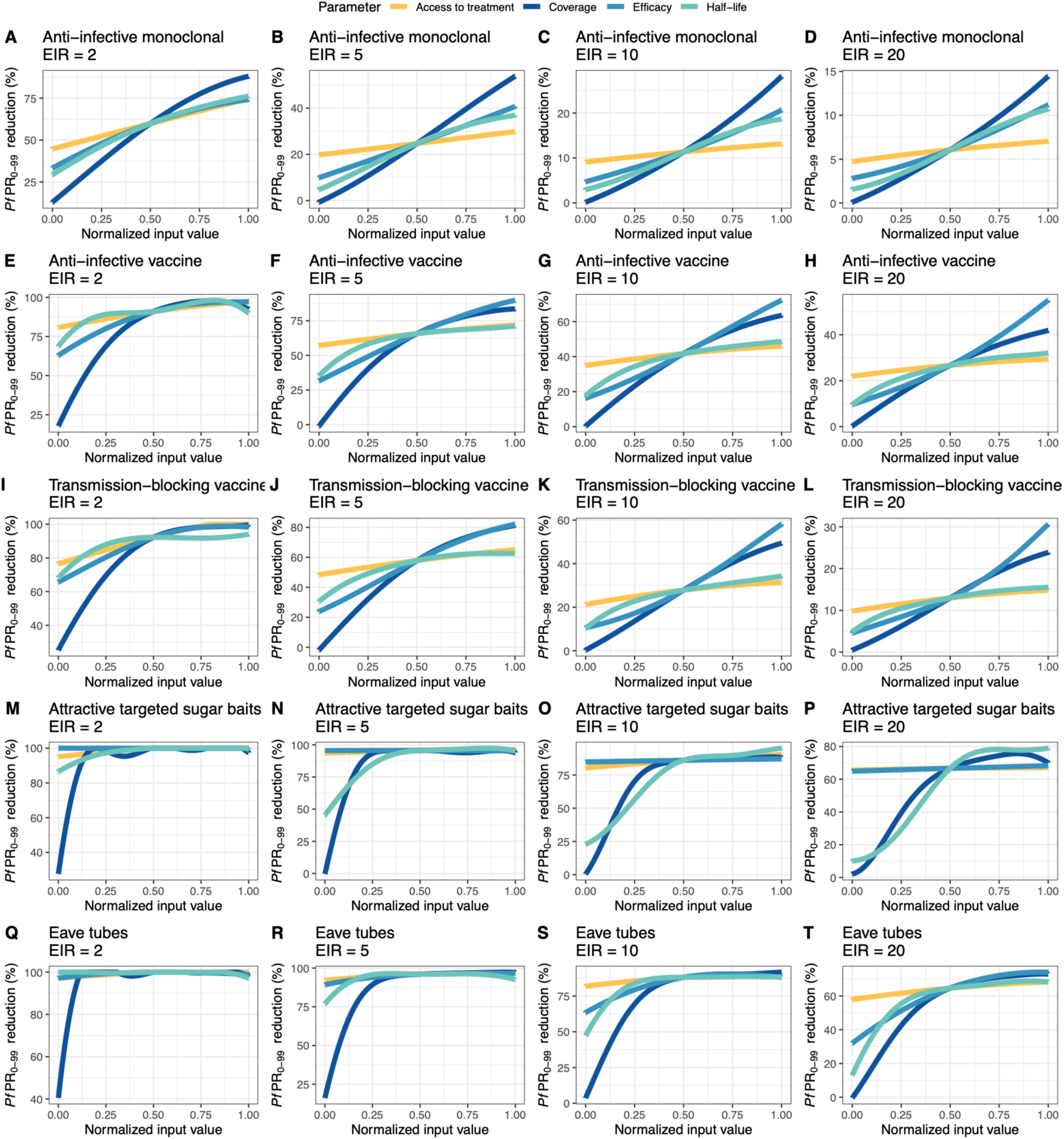
Relationships between input intervention parameters and the predicted immediate *Pf*PR_0-99_ reduction with the trained GP emulator. Each parameter (intervention characteristic) was varied in turn across its defined range (parameter ranges are defined in Table 1) while the remaining parameters were set to their average values. The figures display the immediate *Pf*PR_0-99_ reduction predicted with the GP emulator. A min-max normalization was used to display the varying input values of each of the parameters.

**Fig. S4.6.**
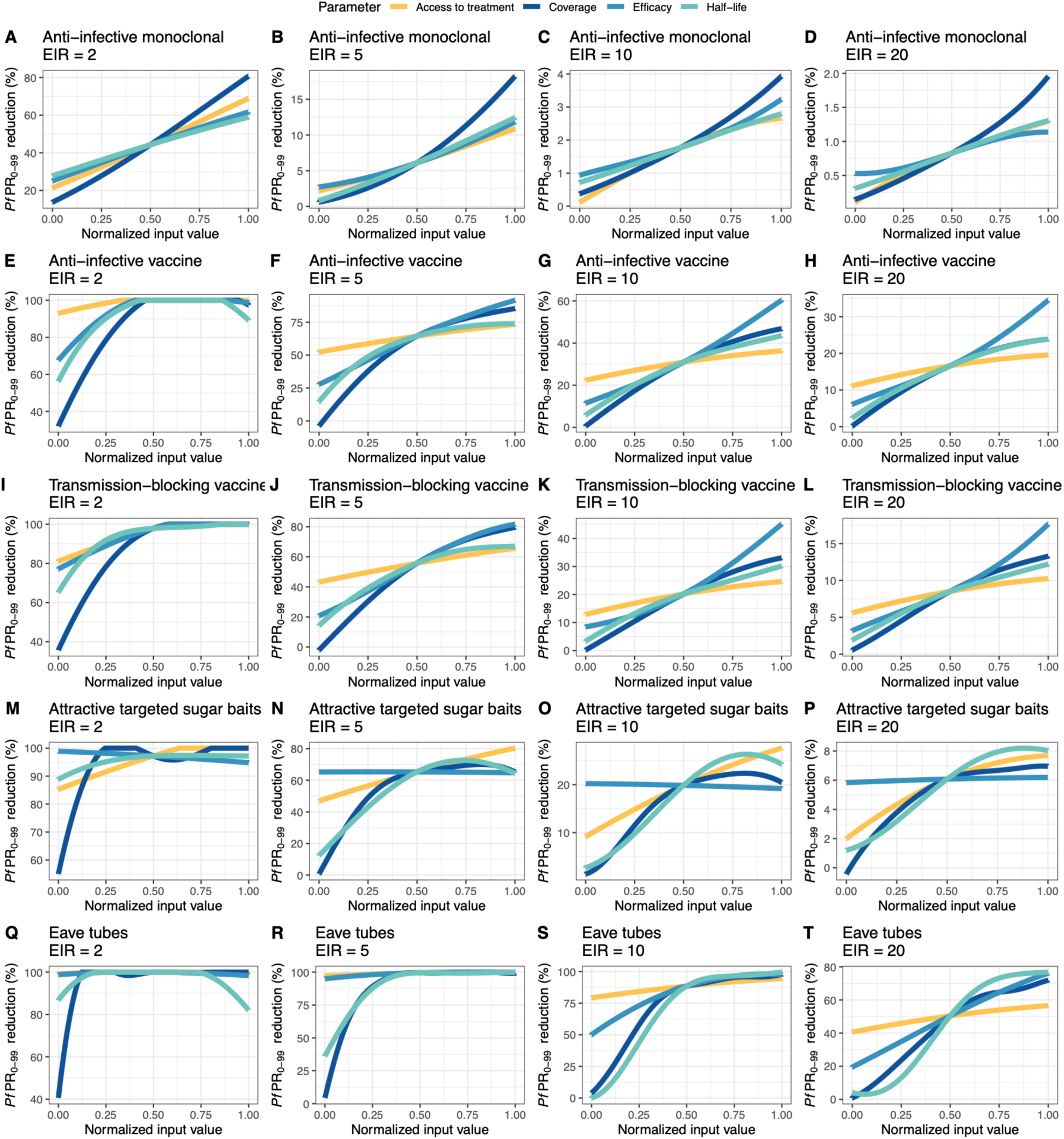
Relationships between input intervention parameters and the predicted long-term *Pf*PR_0-99_ reduction with the trained GP emulator. Each parameter (intervention characteristic) was varied in turn across its defined range (parameter ranges are defined in Table 1) while the remaining parameters were set to their average values. The figures display the long-term *Pf*PR_0-99_ reduction predicted with the GP emulator. A min-max normalization was used to display the varying input values of each of the parameters.

**Fig. S4.7.**
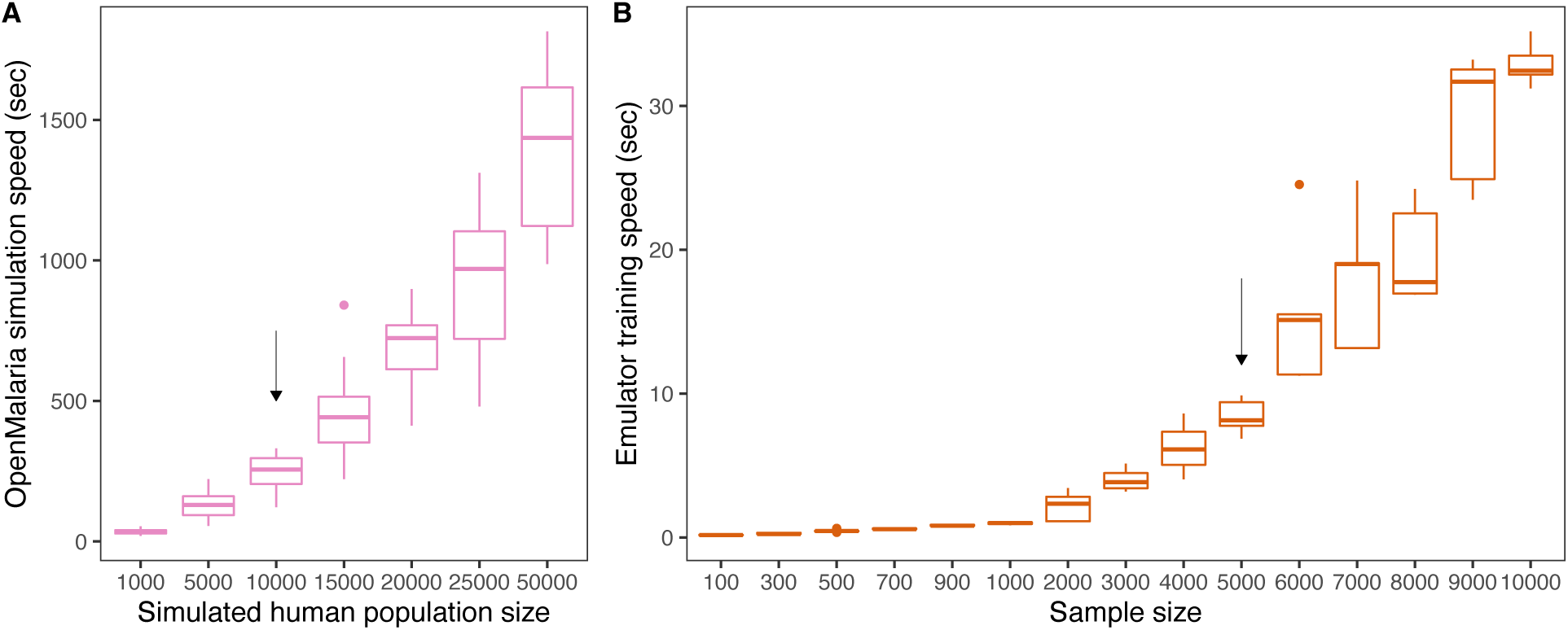
Execution time of OpenMalaria simulations and GP emulator training. (A) CPU execution time of a single OpenMalaria simulation for varying population size. The arrow indicates the population size of 10,000 human hosts used in the present study. (B) CPU time required for training the GP emulator using training sets of OpenMalaria simulations of varying size. The arrow indicates the typical sample size used in the present analysis. In both figures, execution times were estimated 5 times in each case and the resulting distribution was displayed (boxplots).

**Table S4.1.** Performance of the trained GP emulators predicting immediate and long-term intervention impact.

| **Intervention(s)**<br>**(deployment)** | **Training**<br>**set size** | **Test**<br>**set size** | **Cross-validation $r^2$ and**<br>**(mean error)** | **Test set $r^2$ and (mean**<br>**error)** |
| --- | --- | --- | --- | --- |
| Anti-infective<br>monoclonal antibody<br>(once/year) | 10000 | 5000 | **Immediate:** 0.99 (1.02%)<br>**Long:** 0.96 (1.15%) | **Immediate:** 0.99 (0.63%)<br>**Long:** 0.97 (0.68%) |
| Anti-infective<br>monoclonal antibody<br>(twice/year) | 5000 | 2500 | **Immediate:** 0.99 (1.11%)<br>**Long:** 0.97 (1.32%) | **Immediate:** 0.99 (0.91%)<br>**Long:** 0.99 (0.83%) |
| Anti-infective<br>monoclonal antibody +<br>Blood stage drug<br>(once/year) | 10000 | 5000 | **Immediate:** 0.99 (1.34%)<br>**Long:** 0.96 (1.74%) | **Immediate:** 0.99 (1.18%)<br>**Long:** 0.98 (1.05%) |
| Anti-infective<br>monoclonal antibody +<br>Blood stage drug<br>(twice/year) | 5000 | 2500 | **Immediate:** 0.99 (1.26%)<br>**Long:** 0.97 (1.98%) | **Immediate:** 0.99 (0.98%)<br>**Long:** 0.99 (1.12%) |
| Anti-infective vaccine<br>(once/year) | 10000 | 5000 | **Immediate:** 0.99 (1.08%)<br>**Long:** 0.99 (1.3%) | **Immediate:** 0.99 (0.99%)<br>**Long:** 0.99 (1.16%) |
| Anti-infective vaccine<br>+ Blood stage drug<br>(once/year) | 5000 | 2500 | **Immediate:** 0.99 (1.18%)<br>**Long:** 0.99 (1.63%) | **Immediate:** 0.99 (1.57%)<br>**Long:** 0.99 (2.25%) |
| Transmission-blocking<br>vaccine (once/year) | 10000 | 5000 | **Immediate:** 0.99 (1.13)<br>**Long:** 0.99 (1.25%) | **Immediate:** 0.99 (0.89%)<br>**Long:** 0.99 (1.07%) |
| Transmission-blocking<br>vaccine + Blood stage<br>drug (once/year) | 5000 | 2500 | **Immediate:** 0.99 (1.25%)<br>**Long:** 0.99 (1.53%) | **Immediate:** 0.99 (1.68%)<br>**Long:** 0.99 (2.23 %) |
| Attractive targeted<br>sugar baits (once/year) | 5000 | 2500 | **Immediate:** 0.99 (1.26%)<br>**Long:** 0.98 (1.71%) | **Immediate:** 0.99 (1.98%)<br>**Long:** 0.99 (1.19%) |
| Attractive targeted<br>sugar baits (twice/year) | 5000 | 2500 | **Immediate:** 0.99 (1.09%)<br>**Long:** 0.99 (1.98%) | **Immediate:** 0.99 (1.03%)<br>**Long:** 0.99 (1.29%) |
| Eave tubes (once/year) | 10000 | 5000 | **Immediate:** 0.99 (1.11%)<br>**Long:** 0.99 (1.3%) | **Immediate:** 0.99 (0.89%)<br>**Long:** 0.99 (1.26%) |

### 5 Finding minimal intervention properties and results for key determinants of impact

#### 5.1 Finding minimal intervention properties

As summarized in the above and in the Methods, the trained GP models for each transmission setting and intervention were used within a general-purpose optimization scheme to identify minimum intervention properties that reach a defined *Pf*PR_0-99_ reduction goal given operational and intervention constraints.

Let

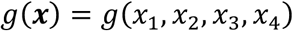

denote the GP model predicting the mean prevalence reduction obtained after deploying an intervention with given characteristics in a transmission setting, with

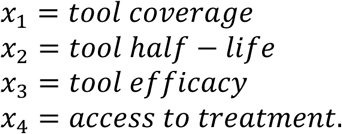

For various levels of *Pf*PR_0-99_ denoted with *p_k_*, each intervention characteristic was optimized separately, keeping the remaining characteristics as well as the level of case management fixed to pre-set levels. Precisely, the optimization procedure searches for

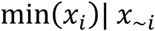

such as

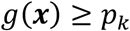

with the constraints:

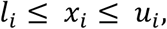

where *l_i_* and *u_i_* are the lower and upper bounds of *x_i_*, respectively and the notation *∼i* is used to represent all the intervention characteristics except *i*. A detailed description of the parameter specifications during optimization for each intervention is provided in Additional file 1: Table S2.1.

To solve the above optimization problem, a general nonlinear augmented Lagrange multiplier method (99, 100) implemented in the R package *Rsolnp* (101) was used. To ensure optimality of the obtained solutions and to avoid local minima, 10 random restarts were chosen among 1,000 uniformly sampled input parameter sets and the optimization procedure was run separately for each restart (implemented in function *gosolnp* in the same R package). To capture the variance of the optimal intervention profile, since the output of a GP model is a distribution, the above optimization problem was solved for several cases, and the distribution of the obtained minima are reported when:

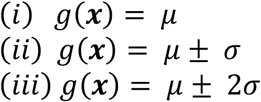

where μ is the predicted mean of the GP model and σ is the standard deviation. Where the nonlinear optimization algorithm did not find any solutions, an additional fine grid search of 10,000 uniformly sampled data points was performed.

Under the simulated levels of case management, before intervention deployment, in seasonal settings, at low-transmission (simulated EIR < 2, corresponding simulated true *Pf*PR_2-10_ < 11.7%), over 75% of simulations reached malaria elimination (*Pf*PR_0-99_ = 0) (Additional file 1: Fig. S3.5). For this reason, the space of obtained prevalence reductions following intervention deployment was rather sparse and the obtained optima were not reliable and often did not converge. Therefore, it was chosen to report minimum intervention profiles for settings with true *Pf*PR_2-10_ >= 11.7% (with RDTs this yields a patent *Pf*PR_2-10_ >= 5.8%).

#### 5.2 Results: Key determinants of impact

**Fig. S5.1.**
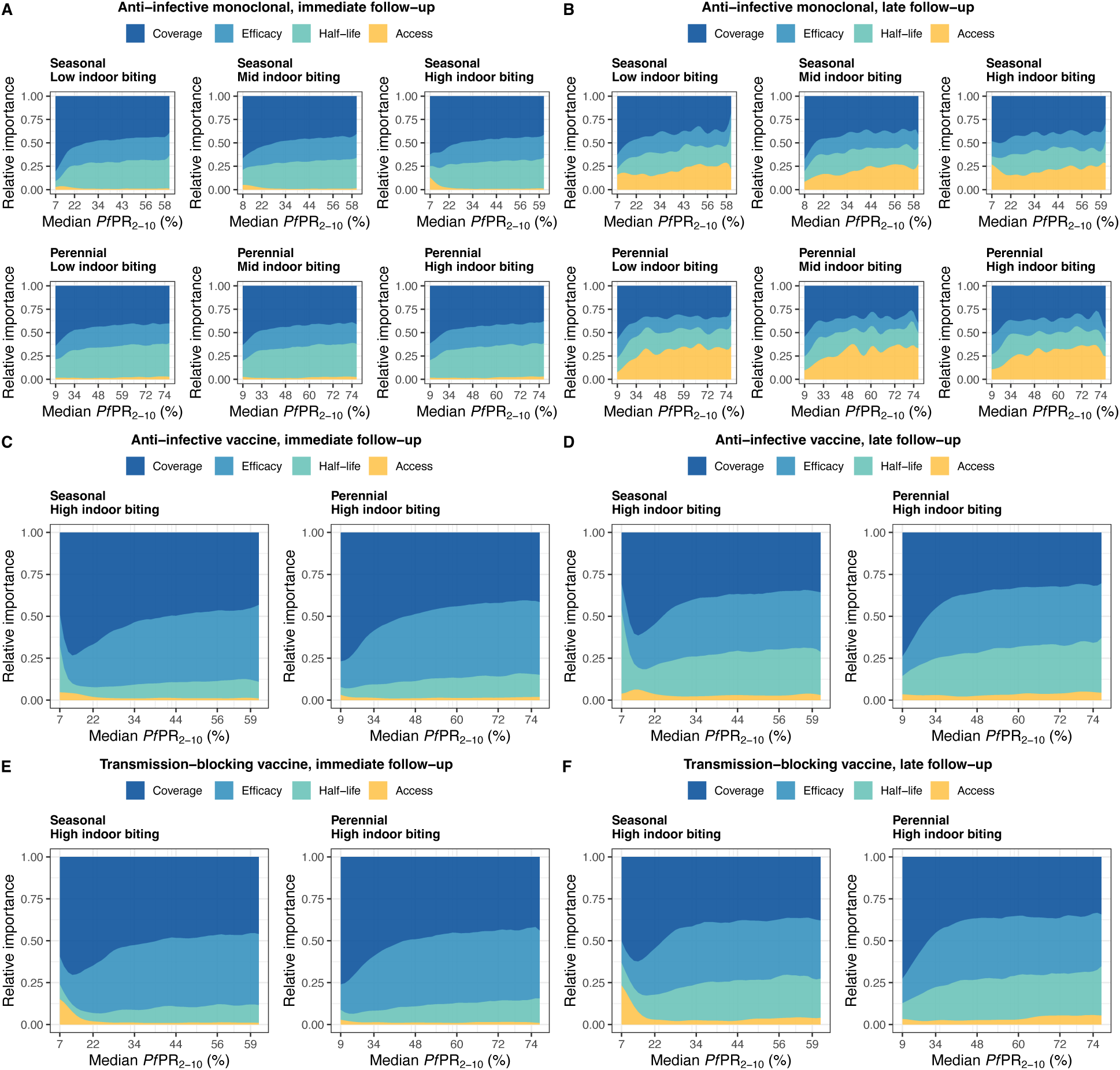
Key drivers of impact for immunological malaria interventions across different transmission settings. Results of sensitivity analysis identifying the determinants of intervention impact on *Pf*PR_0-99_ reduction for anti-infective monoclonal antibodies (A, B), anti-infective vaccines (C, D) and transmission-blocking vaccines (E, F). The distinct colors represent proportions of the GP emulator output variance (relative importance) attributable to intervention efficacy, half-life, deployment coverage, as well as health system access. Determinants of impact are shown for both immediate and late follow-up when interventions are applied once per year for three years in different transmission settings (see full intervention specifications in the Additional file 1: section 1.2.3). The transmission settings are defined by two seasonal settings (seasonal and perennial) and three types of mosquito biting patterns (low, medium. and high indoor biting). The mosquito biting patterns had little to no effect on the results of the sensitivity analysis for these immunological interventions (see results for all settings for monoclonal antibodies in figures A and B). Therefore, only the results for seasonal and perennial settings with high indoor mosquito biting are displayed for the vaccine interventions.

**Fig. S5.2.**
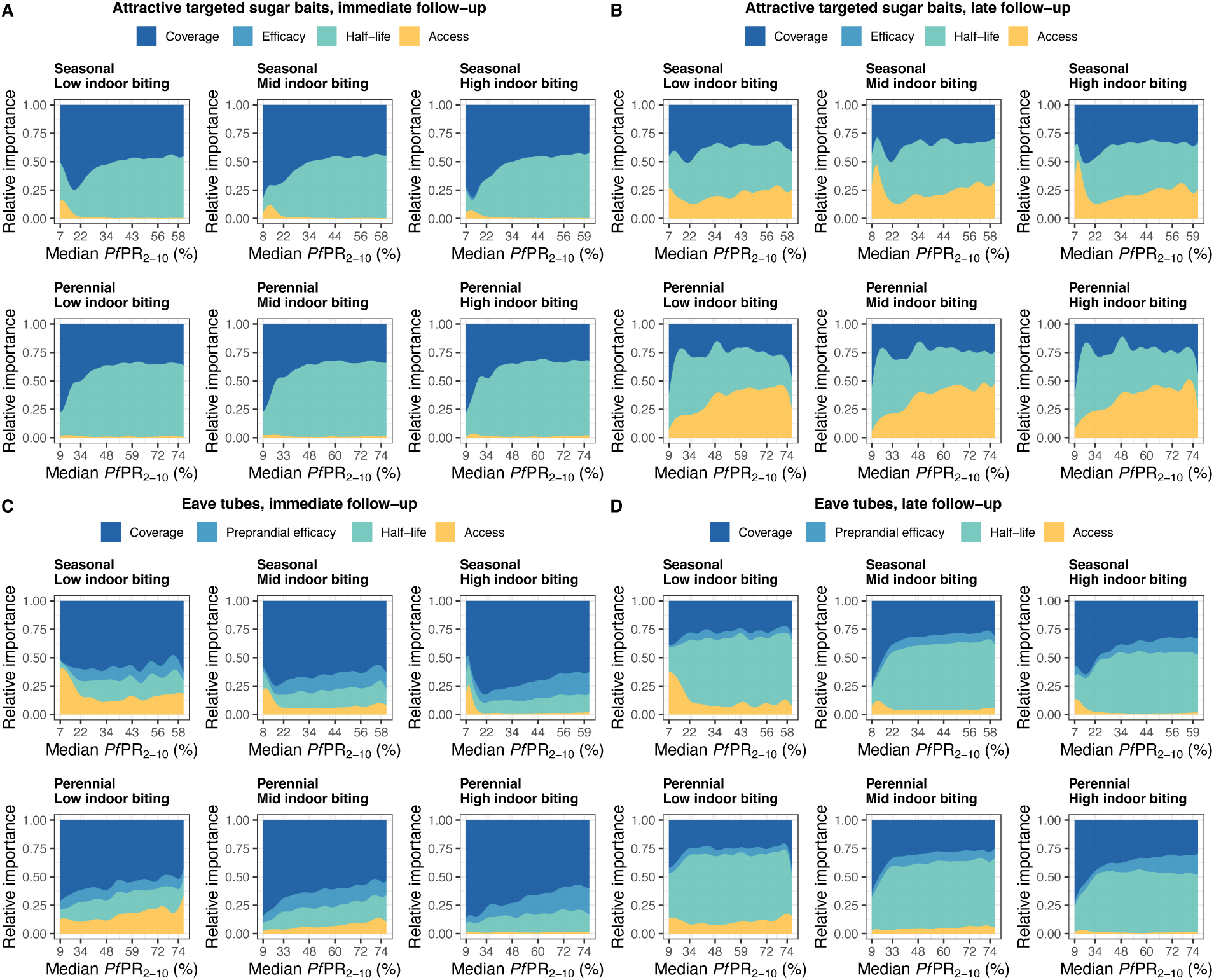
Key drivers of impact for vector control malaria interventions across different transmission settings. Results of sensitivity analysis identifying the determinants of intervention impact on *Pf*PR_0-99_ reduction for attractive targeted sugar baits (A, B) and eave tubes (C, D). The distinct colors represent proportions of the GP emulator output variance (relative importance) attributable to intervention efficacy, half-life, deployment coverage, as well as health system access. Determinants of impact are shown for both immediate and late follow-up when interventions are applied once per year for three years in different transmission settings (see full intervention specifications in the Additional file 1: section 1.2.3). The transmission settings are defined by two seasonal settings (seasonal and perennial) and three types of mosquito biting patterns (low, medium, and high indoor biting). Like for the immunological interventions in the previous figure, we see limited difference between key drivers for attractive targeted sugar baits in different biting settings as mosquitoes sugar feed before indoor or outdoor biting. In contrast, we observe that intervention properties of eave tubes rather than health system access to treatment are larger drivers of impact in indoor biting settings, as mosquitoes in those settings will be more likely to contact the eave tube.

### 6 Results: Feasible landscapes of optimal, constrained intervention profiles

**Fig. S6.1.**
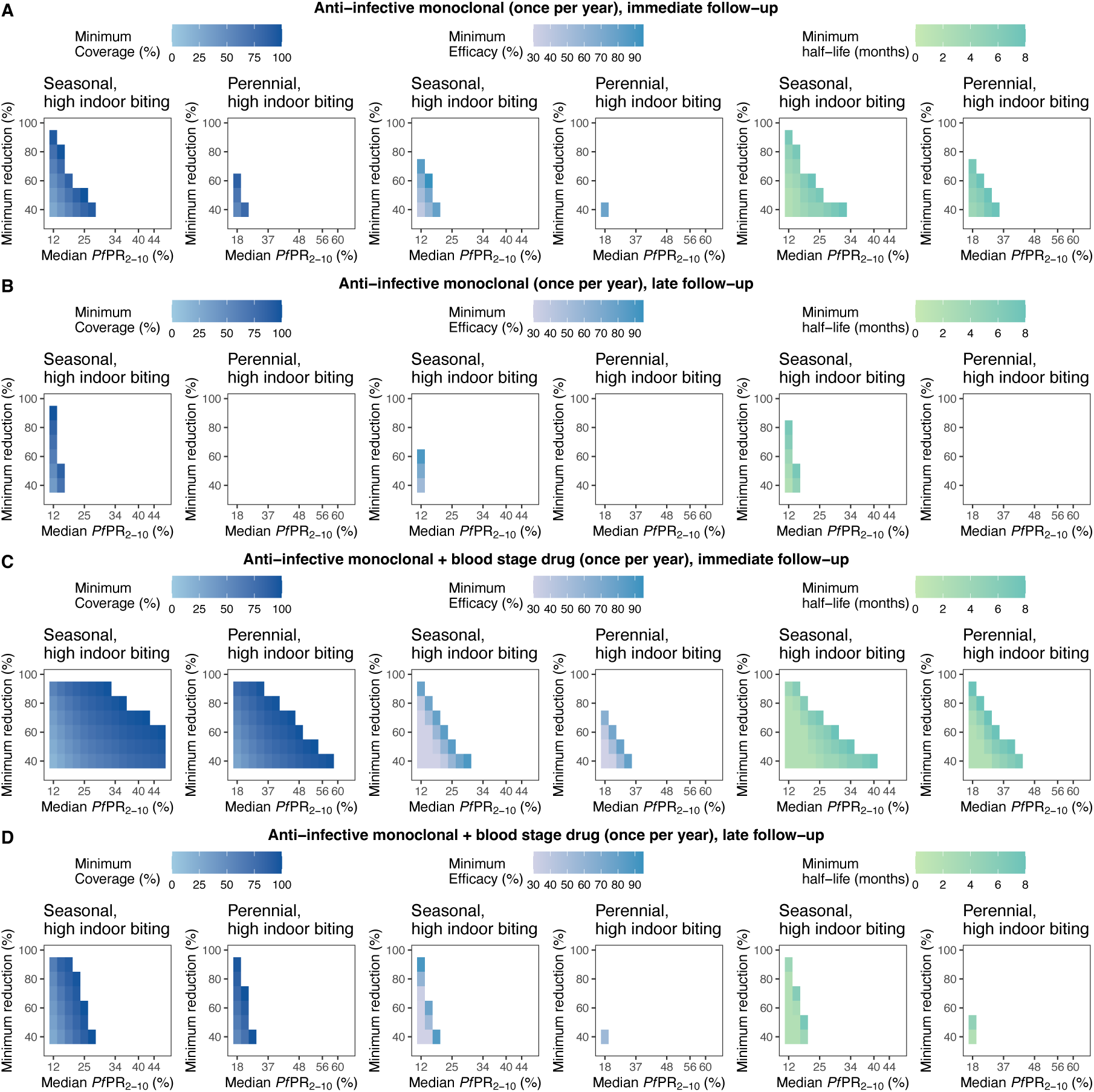
Feasible landscapes of optimal, constrained intervention profiles (TPPs) for an anti-infective monoclonal antibody deployed once per year. The heatmaps represent landscapes of optimal, constrained intervention characteristic profiles (minimum coverage, efficacy, and half-life) required to achieve various health goals (quantified by minimal reduction in PfPR_0-99_, y-axis) across different simulated true PfPR_2-10_ settings (rounded values, x-axis) with seasonal transmission and high indoor mosquito biting. Each intervention characteristic was minimized in turn, while keeping the other characteristics fixed (fixed parameter values for each optimization are specified in Table S2.2). Results are shown for an anti-infective monoclonal antibody delivered alone and assessing immediate (A) and late (B) follow up, as well as when delivered in combination with a blood stage drug assessing immediate (C) and late (D) follow-up. The simulated case management level (E_5_) for all the displayed optimization analyses was assumed 25%.

**Fig. S6.2.**
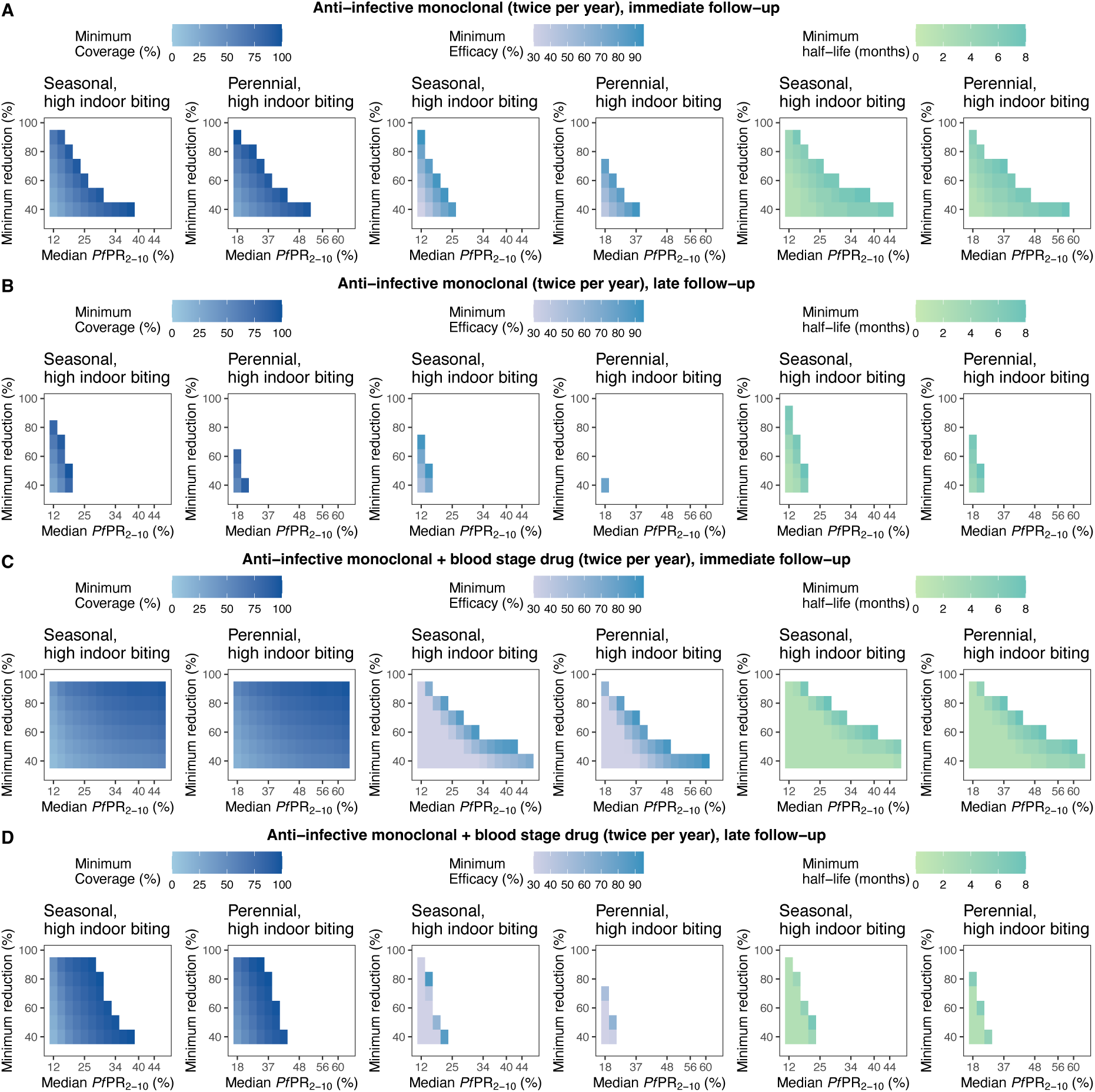
Feasible landscapes of optimal, constrained intervention profiles (TPPs) for an anti-infective monoclonal antibody deployed twice per year. The heatmaps represent landscapes of optimal, constrained intervention characteristic profiles (minimum coverage, efficacy, and half-life) required to achieve various health goals (quantified by minimal reduction in *Pf*PR_0-99_, y-axis) across different simulated true *Pf*PR_2-10_ settings (rounded values, x-axis) with seasonal transmission and high indoor mosquito biting. Each intervention characteristic was minimized in turn, while keeping the other characteristics fixed (fixed parameter values for each optimization are specified in Table S2.2). Results are shown for an anti-infective monoclonal antibody delivered alone and assessing immediate (A) and late (B) follow up, as well as when delivered in combination with a blood stage drug assessing immediate (C) and late (D) follow-up. The simulated case management level (E_5_) for all the displayed optimization analyses was assumed 25%.

**Fig. S6.3.**
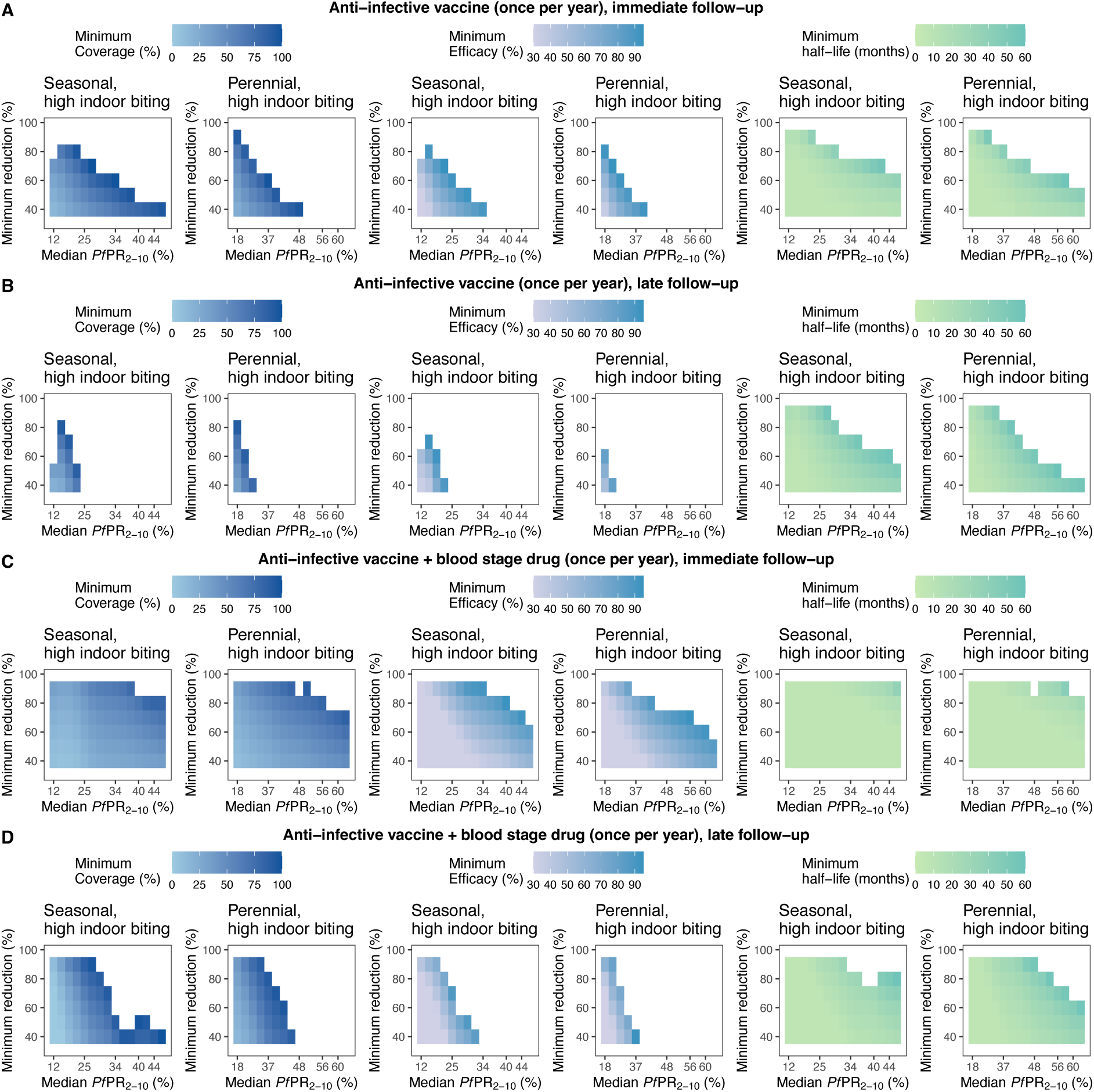
Feasible landscapes of optimal, constrained intervention profiles (TPPs) for an anti-infective vaccine deployed once per year. The heatmaps represent landscapes of optimal, constrained intervention characteristic profiles (minimum coverage, efficacy, and half-life) required to achieve various health goals (quantified by minimal reduction in *Pf*PR_0-99_, y-axis) across different simulated true *Pf*PR_2-10_ settings (rounded values, x-axis) with seasonal transmission and high indoor mosquito biting. Each intervention characteristic was minimized in turn, while keeping the other characteristics fixed (fixed parameter values for each optimization are specified in Table S2.2). Results are shown for an anti-infective vaccine delivered alone and assessing immediate (A) and late (B) follow up, as well as when delivered in combination with a blood stage drug assessing immediate (C) and late (D) follow-up. The simulated case management level (E_5_) for all the displayed optimization analyses was assumed 25%.

**Fig. S6.4.**
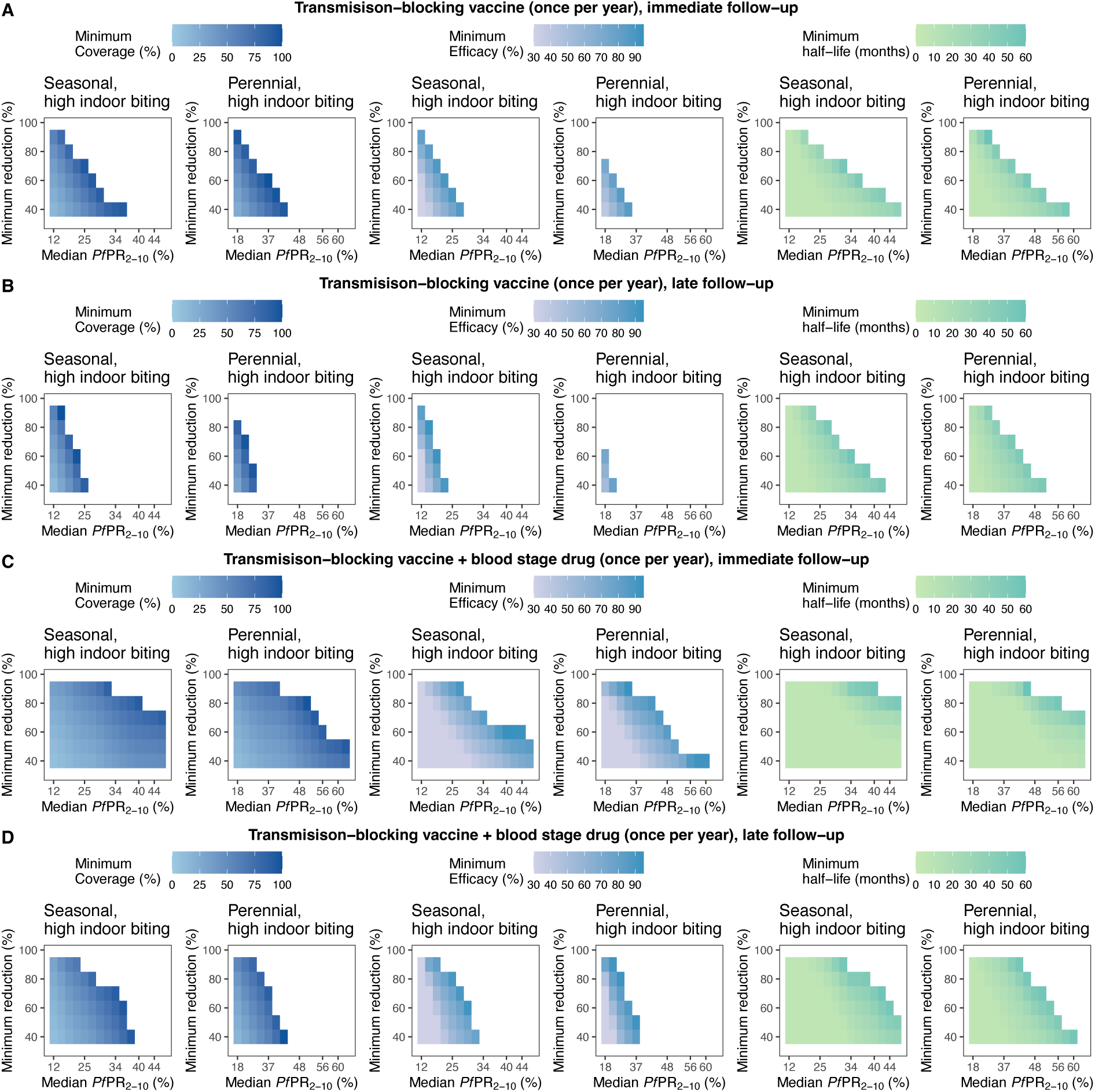
Feasible landscapes of optimal, constrained intervention profiles (TPPs) for a transmission-blocking vaccine deployed once per year. The heatmaps represent landscapes of optimal, constrained intervention characteristic profiles (minimum coverage, efficacy, and half-life) required to achieve various health goals (quantified by minimal reduction in *Pf*PR_0-99_, y-axis) across different simulated true *Pf*PR_2-10_ settings (rounded values, x-axis) with seasonal transmission and high indoor mosquito biting. Each intervention characteristic was minimized in turn, while keeping the other characteristics fixed (fixed parameter values for each optimization are specified in Table S2.2). Results are shown for a transmission-blocking vaccine delivered alone and assessing immediate (A) and late (B) follow up, as well as when delivered in combination with a blood stage drug assessing immediate (C) and late (D) follow-up. The simulated case management level (E_5_) for all the displayed optimization analyses was assumed 25%.

**Fig. S6.5.**
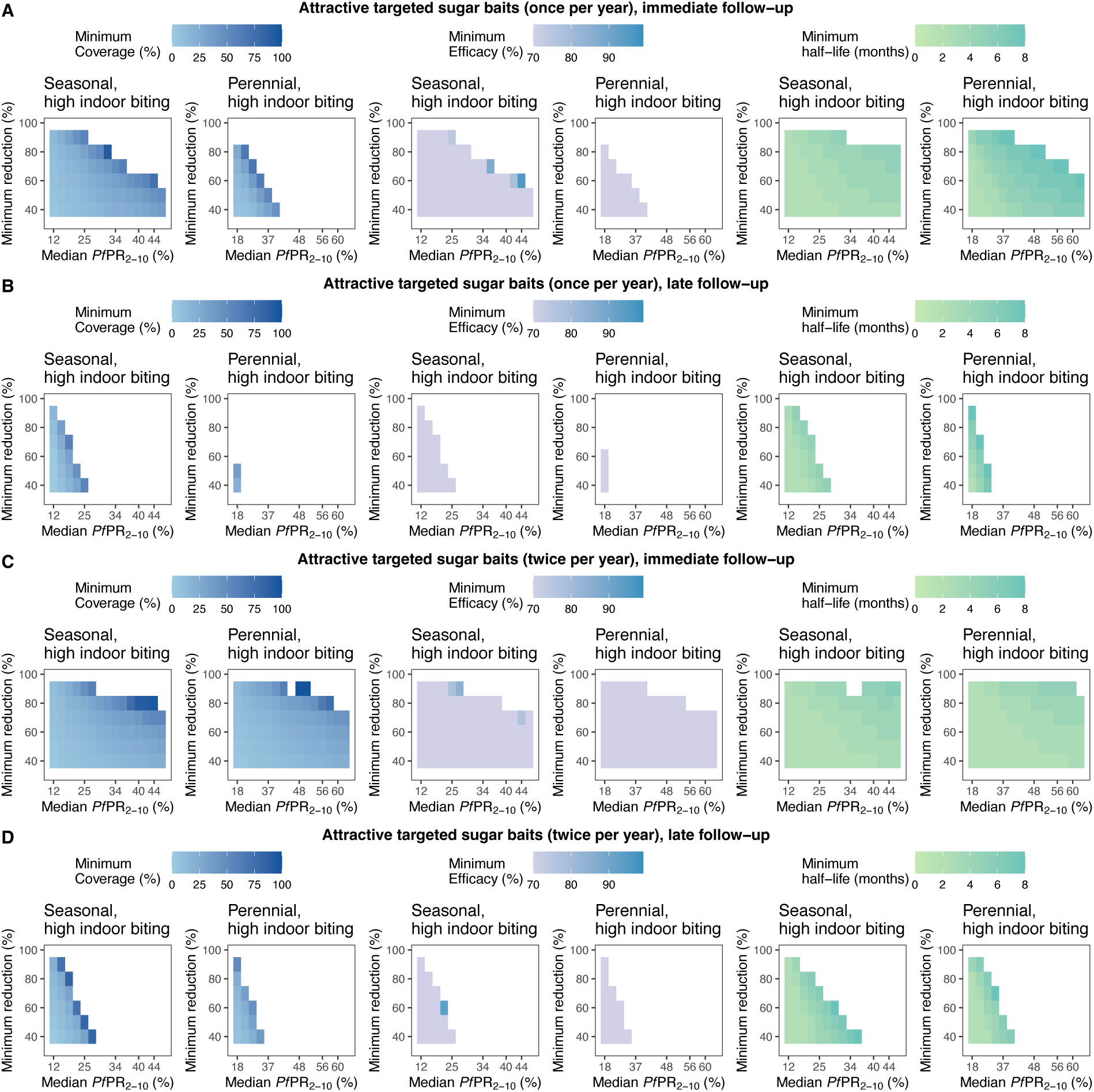
Feasible landscapes of optimal, constrained intervention profiles (TPPs) for attractive targeted sugar baits deployed once or twice per year. The heatmaps represent landscapes of optimal, constrained intervention characteristic profiles (minimum coverage, efficacy, and half-life) required to achieve various health goals (quantified by minimal reduction in *Pf*PR_0-99_, y-axis) across different simulated true *Pf*PR_2-10_ settings (rounded values, x-axis) with seasonal transmission and high indoor mosquito biting. Each intervention characteristic was minimized in turn, while keeping the other characteristics fixed (fixed parameter values for each optimization are specified in Table S2.2). Results are shown for attractive targeted sugar baits delivered alone once per year and assessing immediate (A) and late (B) follow up, as well as when delivered twice per year assessing immediate (C) and late (D) follow-up. The simulated case management level (E_5_) for all the displayed optimization analyses was assumed 25%.

**Fig. S6.6.**
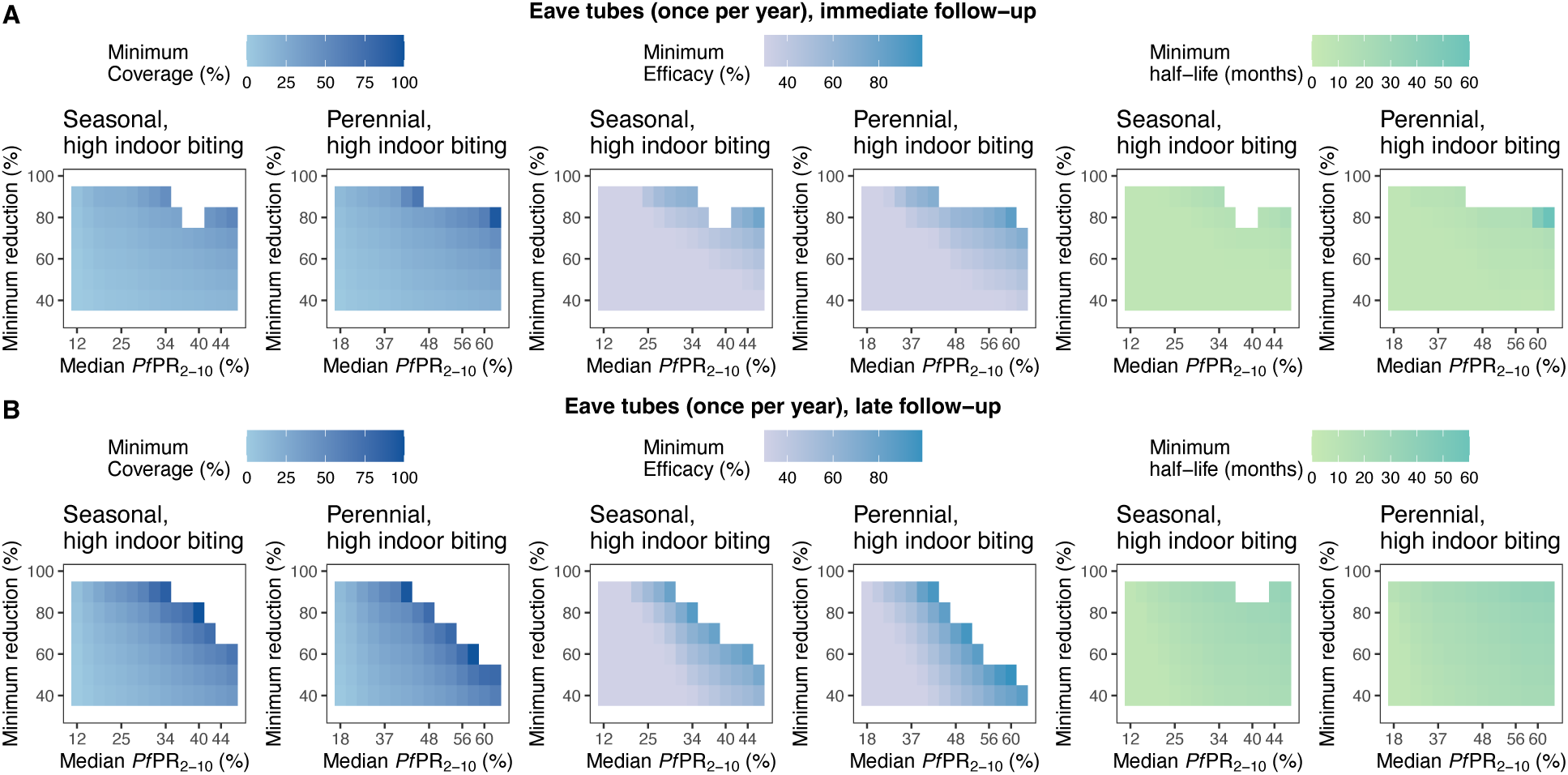
Feasible landscapes of optimal, constrained intervention profiles (TPPs) for eave tubes deployed once per year. Heatmaps represent landscapes of optimal, constrained intervention characteristic profiles (minimum coverage, efficacy, and half-life) required to achieve various health goals (quantified by minimal reduction in *Pf*PR_0-99_, y-axis) across different simulated true *Pf*PR_2-10_ settings (rounded values, x-axis) with seasonal or perennial transmission and high indoor mosquito biting (results for other biting patterns not shown as they are similar). Each intervention characteristic was minimized in turn, while keeping the other characteristics fixed (fixed parameter values for each optimization are specified in Table S2.2). Results are shown for eave tubes delivered alone and assessing immediate (A) and late (B) follow up. The simulated case management level (E_5_) for all the displayed optimization analyses was assumed 25%.

### 7 Results: Optimal intervention profiles

**Fig. S7.1.**
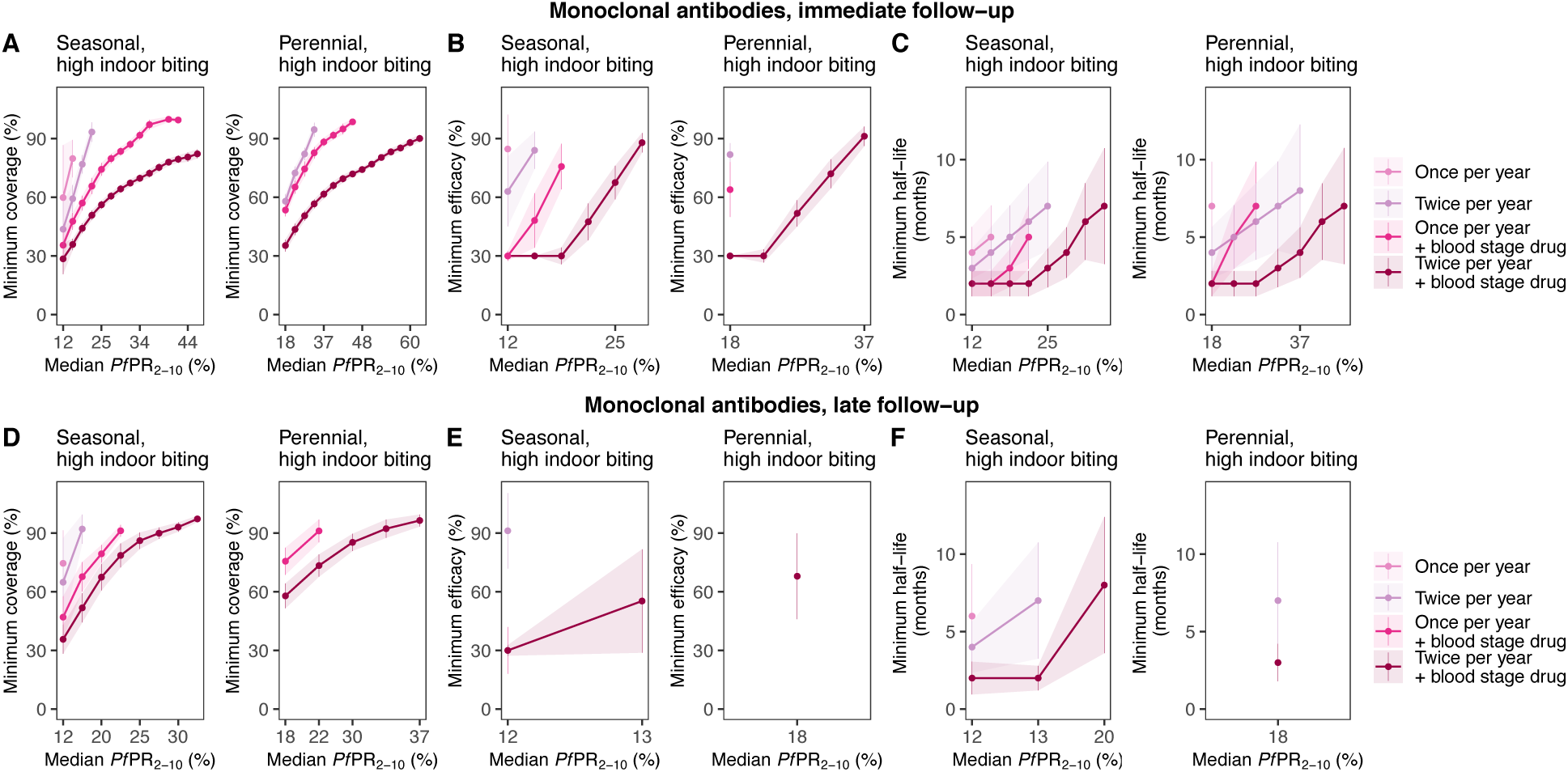
Optimal intervention profiles (TPPs) for anti-infective monoclonal antibodies under various deployment regimes to achieve a PfPR0-99 reduction of at least 70%. Each figure displays minimum, constrained intervention characteristic profiles (minimum coverage, efficacy, and half-life, y-axis) required to achieve a minimal reduction in *Pf*PR_0-99_ of 70% across different simulated true *Pf*PR_2-10_ settings (rounded values, x-axis) with seasonal transmission and high indoor mosquito biting. Each intervention characteristic was minimized in turn, while keeping the other characteristics fixed (fixed parameter values for each optimization are specified in Table S2.2). Results are shown when assessing *Pf*PR_0-99_ reduction at immediate (A-C) and late (D-F) follow up. The simulated case management level (E_5_) for all the displayed optimization analyses was assumed 25%.

**Fig. S7.2.**
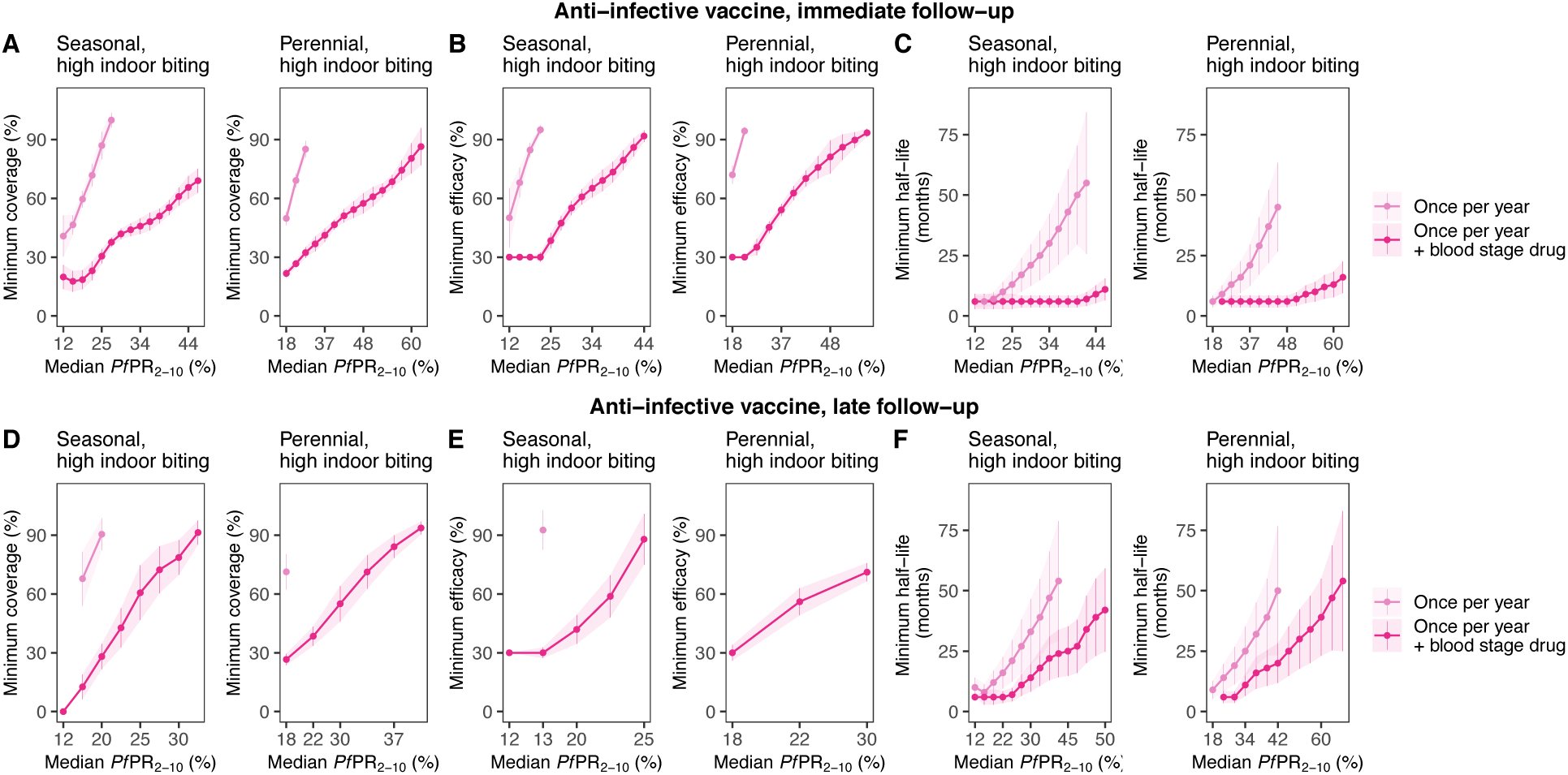
Optimal intervention profiles (TPPs) for anti-infective vaccines under various deployment regimes to achieve a PfPR0-99 reduction of at least 70%. Each figure displays minimum, constrained intervention characteristic profiles (minimum coverage, efficacy, and half-life, y-axis) required to achieve a minimal reduction in *Pf*PR_0-99_ of 70% across different simulated true *Pf*PR_2-10_ settings (rounded values, x-axis) with seasonal transmission and high indoor mosquito biting. Each intervention characteristic was minimized in turn, while keeping the other characteristics fixed (fixed parameter values for each optimization are specified in Table S2.2). Results are shown when assessing *Pf*PR_0-99_ reduction at immediate (A-C) and late (D-F) follow up. The simulated case management level (E_5_) for all the displayed optimization analyses was assumed 25%.

**Fig. S7.3.**
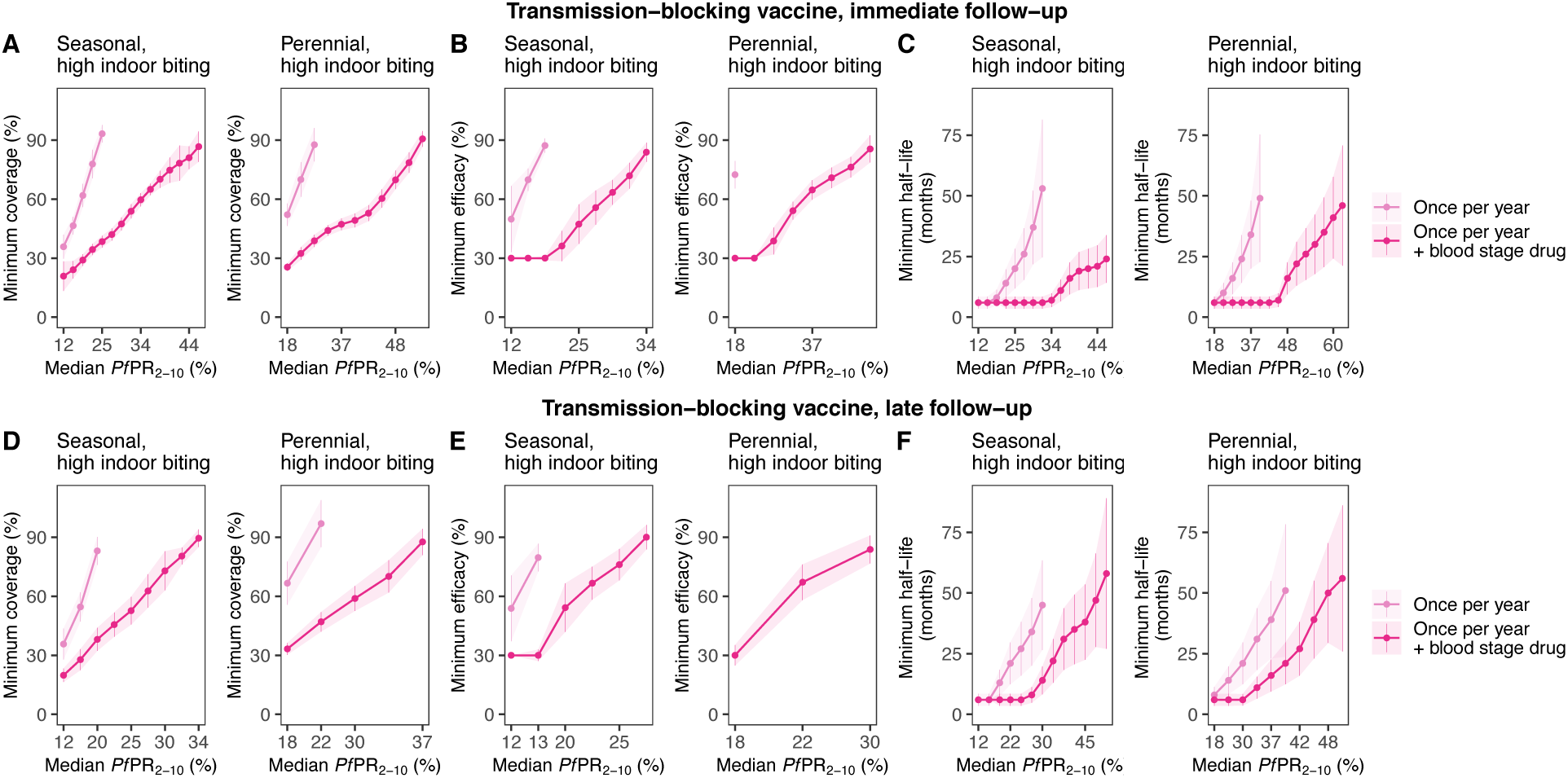
Optimal intervention profiles (TPPs) for transmission-blocking vaccines under various deployment regimes to achieve a *Pf*PR_0-99_ reduction of at least 70%. Each figure displays minimum, constrained intervention characteristic profiles (minimum coverage, efficacy, and half-life, y-axis) required to achieve a minimal reduction in *Pf*PR_0-99_ of 70% across different simulated true *Pf*PR_2-10_ settings (rounded values, x-axis) with seasonal transmission and high indoor mosquito biting. Each intervention characteristic was minimized in turn, while keeping the other characteristics fixed (fixed parameter values for each optimization are specified in Table S2.2). Results are shown when assessing *Pf*PR_0-99_ reduction at immediate (A-C) and late (D-F) follow up. The simulated case management level (E_5_) for all the displayed optimization analyses was assumed 25%.

**Fig. S7.4.**
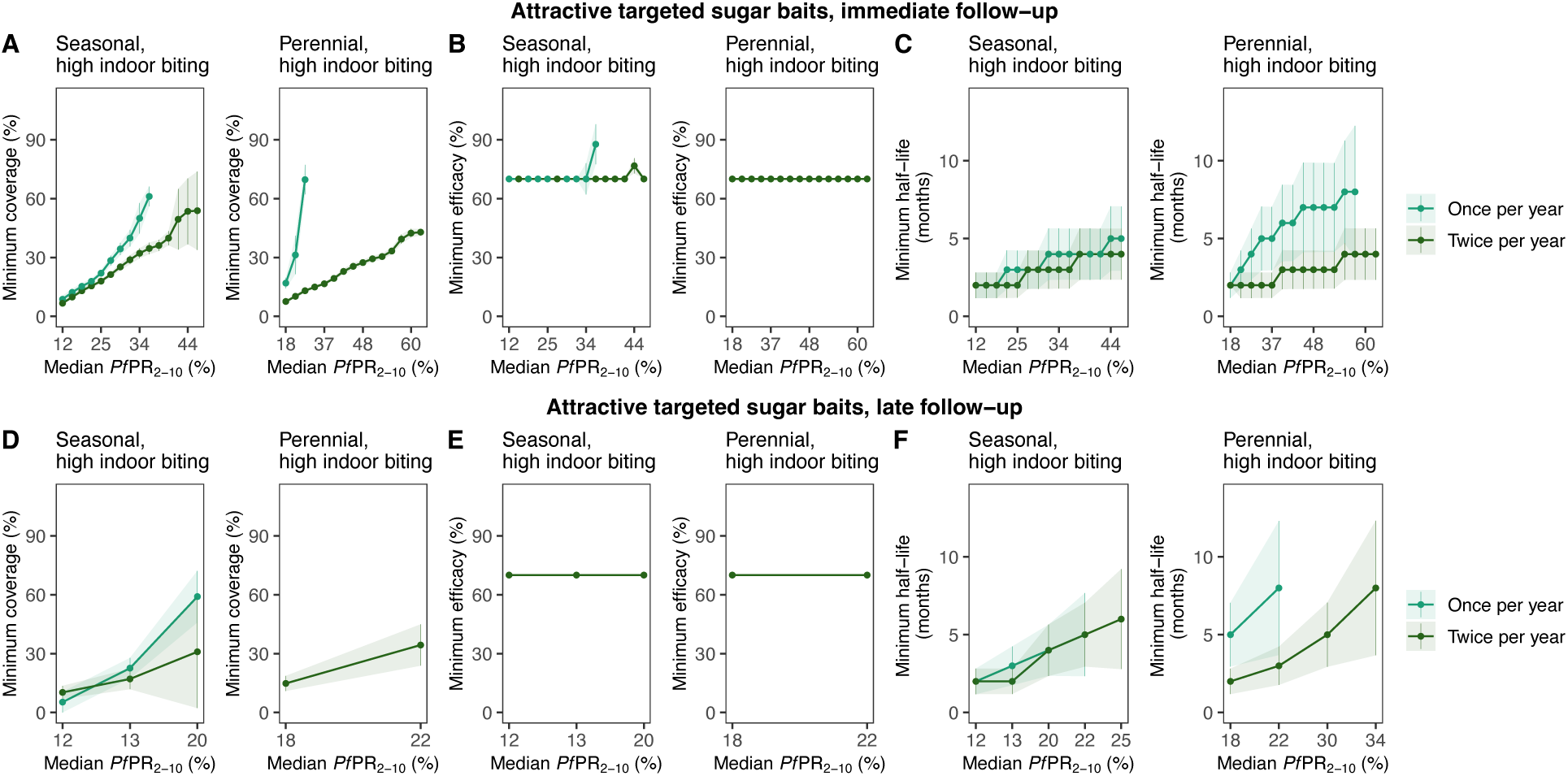
Optimal intervention profiles (TPPs) for attractive targeted sugar baits under various deployment regimes to achieve a PfPR0-99 reduction of at least 70%. Each figure displays minimum, constrained intervention characteristic profiles (minimum coverage, efficacy, and half-life, y-axis) required to achieve a minimal reduction in *Pf*PR_0-99_ of 70% across different simulated true *Pf*PR_2-10_ settings (rounded values, x-axis) with seasonal transmission and high indoor mosquito biting. Each intervention characteristic was minimized in turn, while keeping the other characteristics fixed (fixed parameter values for each optimization are specified in Table S2.2). Results are shown when assessing *Pf*PR_0-99_ reduction at immediate (A-C) and late (D-F) follow up. The simulated case management level (E_5_) for all the displayed optimization analyses was assumed 25%.

**Fig. S7.5.**
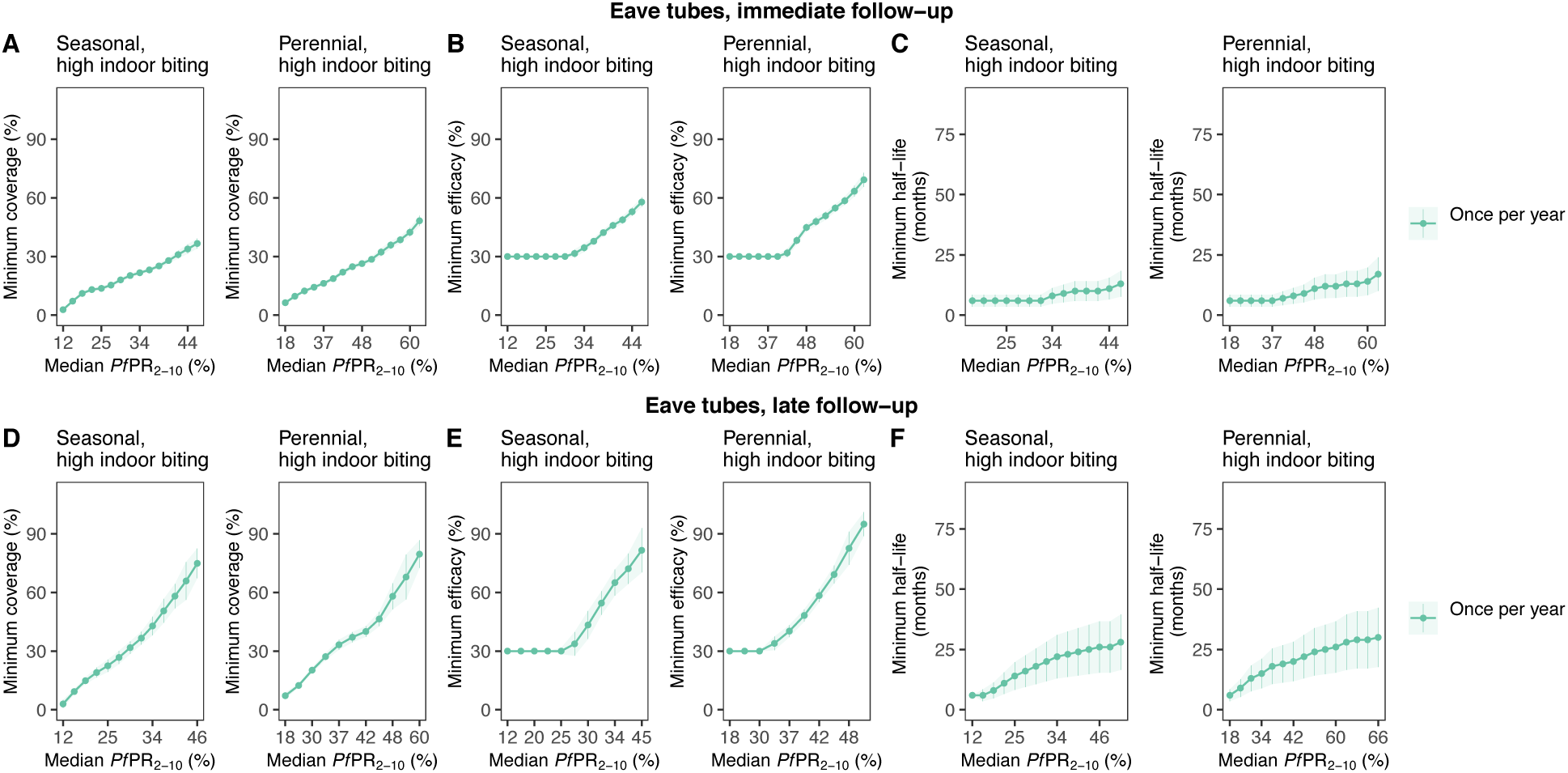
Optimal intervention profiles (TPPs) for eave tubes to achieve a PfPR0-99 reduction of at least 70%. Each figure displays minimum, constrained intervention characteristic profiles (minimum coverage, efficacy, and half-life, y-axis) required to achieve a minimal reduction in PfPR0-99 of 70% across different simulated true PfPR2-10 settings (rounded values, x-axis) with seasonal transmission and high indoor mosquito biting. Each intervention characteristic was minimized in turn, while keeping the other characteristics fixed (fixed parameter values for each optimization are specified in Table S2.2). Results are shown when assessing PfPR0-99 reduction at immediate (A-C) and late (D-F) follow up. The simulated case management level (E5) for all the displayed optimization analyses was assumed 25%.

